# Brick kiln pollution and its impact on health: A systematic review and meta-analysis

**DOI:** 10.1101/2023.11.16.23298642

**Authors:** Laura Nicolaou, Fiona Sylvies, Isabel Veloso, Katherine Lord, Ram K Chandyo, Arun K Sharma, Laxman P Shrestha, David L Parker, Steven M Thygerson, Peter F DeCarlo, Gurumurthy Ramachandran, William Checkley

## Abstract

**Background:** Brick kiln emissions adversely affect air quality and the health of workers and individuals living near the kilns; however, evidence of the impacts of brick kiln pollution remains limited.

**Methods:** We conducted a systematic review of brick kiln pollution (emissions, source contributions and personal exposures) and its effects on health. We extracted articles from electronic databases and through manual citation searching. We estimated pooled, sample-size-weighted means and standard deviations for personal exposures by job type; computed mean emission factors and pollutant concentrations by brick kiln design; and meta-analyzed differences in means or proportions for health outcomes between brick kiln workers (BKWs) and controls or for participants living near or far away from kilns.

**Results:** Our search yielded 1015 articles; 208 (20%) were assessed for eligibility and 101 (10%) were included in our review. We identified three additional studies through manual searching. Of 104 studies, 74 (71%) were conducted in South Asia. The most evaluated pollutants were particulate matter (PM; n=48), sulfur dioxide (SO_2_; n=24) and carbon monoxide (CO; n=22), and the most evaluated health outcomes were respiratory health (n=34) and musculoskeletal disorders (n=9). PM and CO emissions were higher among traditional than improved brick kilns. Mean respirable silica exposures were only measured in 4 (4%) studies and were as high as 620 μg/m^3^, exceeding the NIOSH recommended exposure limit by a factor of over 12. BKWs had consistently worse lung function, more respiratory symptoms, more musculoskeletal complaints, and more inflammation when compared to unexposed participants across studies; however, most studies had a small sample size and did not fully describe methods used for sampling or data collection.

**Discussion:** On average, BKWs had worse health outcomes when compared to unexposed controls but study quality supporting the evidence was low. Few studies reported silica concentrations or personal exposures, but the few that did suggest that exposures are high. Further research is needed to better understand the relationship between brick kiln pollution and health among workers, and to evaluate exposure mitigation strategies.

## INTRODUCTION

Approximately 1,500 billion bricks are produced every year, and ∼90% are produced in Asia.^1,2^ South Asia is the second largest brick-producing region after China, with an estimated annual production of 310 billion bricks.^1^ India, Pakistan, Bangladesh and Nepal are the biggest producers in this region, accounting for nearly 25% of global brick production.^1^ The brick kiln industry in LMICs is labor-intensive, and most kilns are energy inefficient and highly polluting.^1–3^ Mechanised and efficient technologies are limited in number, making up <1% of the 150,000 kilns in South Asia.^1–4^ Compounding this problem, many brick kilns operate in the informal sector with little or no regulation by local governments on labor or kiln emissions.^1,2^

Brick manufacturing involves a number of processes, typically starting with digging or mining of topsoil, mixing and molding of wet clay, and sun-drying of the green bricks.^5^ Once dried, green bricks are carried, usually on the head or back, stacked inside the kiln, and fired. Fired, or red, bricks are then manually carried out of the kiln. Bricks are primarily fired using coal or biomass, but other fuels such as rubber tires, motor oils, trash and plastic are also common.^6–9^ Throughout the brick making processes, workers are exposed to various airborne pollutants. Clay and brick dust contains high concentrations of silica, while the smoke emitted during brick firing contains particulate matter (PM) and gaseous pollutants including sulfur dioxide (SO_2_), carbon monoxide (CO), and nitrogen oxides (NO_x_). An estimated 16 million workers in South Asia alone are exposed to these hazardous pollutants.^1^ Brick kilns are also a major contributor to ambient air pollution and are responsible for up to 91% of total PM emissions in some cities.^1^

Reports from the 2016 Global Burden of Disease highlight the burden of chronic respiratory diseases caused by occupational exposures, namely chronic obstructive pulmonary disease (COPD), asthma, and pneumoconioses such as silicosis and asbestosis.^10^ Among high-risk occupations, those that involve increased exposure to both smoke and dust place workers at the highest risk for developing chronic respiratory symptoms and illnesses, as is the case with BKWs.^11^

Despite the contribution of the brick kiln industry to ambient air pollution and community-wide respiratory illness, data on kiln pollution remains limited.^4^ Most reports on the consequences of brick kiln pollution have been based in urban areas in South Asia. This review expands the evaluation of brick kiln emissions and their health impacts globally, with particular focus on LMICs. We synthesize existing evidence on the effects of brick kiln emissions on the environment and health, identify current gaps in the literature, and discuss implemented interventions to lower brick kiln emissions.

## METHODS

### Search strategy and data sources

The search strategy is provided in the supplementary material and described in the PROSPERO protocol (PROSPERO 2020 CRD42020221833). Sources were extracted from electronic databases including EMBASE, MEDLINE, Web of Science, SciELO, WHO Global Index Medicus, Cochrane Database of Systematic Reviews, and World Bank, as well as reference lists from identified studies and reviews. Electronic searching was carried out in December 2020. Manual and electronic searches were conducted between December 2021 and October 2022 and an updated electronic search was conducted in July 2023. Languages were restricted to English, Nepali, Spanish, Italian, and French. No date limits were applied. Sources identified from electronic databases were combined, duplicates removed, and articles screened for relevance based on title and abstract by two reviewers (LN and FS). Disagreements between the two reviewers were adjudicated by a third reviewer (WC).

### Data extraction

The full text was then acquired for all sources identified as potentially relevant. Studies were included if they provided (1) quantitative data on brick kiln pollutants, including concentrations, emission factors and source contributions to ambient air pollution; or (2) health outcomes among brick kiln workers (BKWs) or community members living near brick kilns; or (3) a comparison between brick kiln exposed and unexposed participants. Studies were excluded if they were conference abstracts or proceedings; provided data on pollution that does not directly impact human health (e.g., relevant to soil health or climate change); did not provide concrete data on health outcomes, pollutants at kiln sites or contribution of brick kiln emissions to the surrounding air pollution (e.g., ambient pollutant concentrations measured in the vicinity of kilns; source apportionment where brick kiln emissions are lumped together with other sources); reported modeled, rather than measured, exposures or outcomes (e.g., estimates of brick kiln emissions using existing emissions inventories and atmospheric dispersion modelling or cancer risk assessment using the incremental lifetime cancer risk approach); or were case reports, reviews and studies with duplicate data. In case of disagreement between reviewers, articles were adjudicated by the third reviewer. Articles were then sorted by categories (Pollution, Health, Pollution and Health) and summary tables were created including the following details for pollutant-related studies: region, sampling dates, measurement type, sampling location, sample size, number and duration of measurements, and pollutant(s) reported. Summary tables for health outcomes studies included region, dates, study design, characteristics of the study population (age, gender, sample size, inclusion criteria), and outcome(s) reported.

### Methodological quality assessment

For studies reporting health outcomes, we assessed methodological quality using the Newcastle-Ottawa scale, which provides a standardized approach to grade the quality of nonrandomized studies. The Newcastle-Ottawa scale consists of eight criteria across three domains including selection of the study groups, comparability of the study groups, and ascertainment of the exposure or outcome for case-control and cohort studies, respectively.^12^ Since a validated risk of bias tool for exposure studies does not exist, we developed a 6-item scale to assess the quality of papers reporting pollution data. Two of three reviewers (LN, KL, WC) scored the studies independently and disagreements were resolved by the third.

### Statistical methods

We estimated pooled, sample-size-weighted means and standard deviations for personal exposures by job type. We could not do the same for emission factors and concentrations because the sampling intervals were too variable and as a result variances were not on the same time scales. Instead, we computed means by brick kiln design. We categorized Bull’s trench (BTK), clamp (CK), downdraft (DDK), traditional-campaign (TCK), traditional-fixed (TFK) and traditional-improved (TIK) kilns as traditional, while forced-draft zigzag (FDZ), Hoffmann (HK), Marquez (MK) and double-domed Marquez (MK2), natural draft zigzag (NDZ), tunnel (TK) and vertical shaft (VSBK) kilns were classified as improved designs. Emission factors and concentrations below the limit of detection (LOD) were assigned a value equal to the *LOD/√*2.

Analyses were performed on natural-log transformed data and values were then back-transformed to obtain geometric means and confidence intervals.^13^ We converted results reported in ppm to mg/m^3^ using mg/m^&^ = ppm <u>^’(^</u>, where *MW* is the molecular weight (g/mol) and *Vm* is the molar gas volume (L/mol). Assuming a pressure of 1 atmosphere and a temperature of 25 ℃, *V_m_* = 24.465 L/mol for ideal gases. For VOCs, which were measured using an Aeroqual 500, we applied the device’s conversion factor of 2.5 to determine concentrations in mg/m^3^._14_ For studies that did not report the modified combustion efficiency (MCE), we estimated it as MCE = τιCO_2_/(τιCO_2_+τιCO), where τιCO_2_ and τιCO are the total amounts of CO_2_ and CO emitted, respectively. Non-physical and inconsistent data (e.g. CO_2_ emission factors >3667 g/kg fuel^15^; higher MCE in fugitive than flue gas emissions^16^) were flagged and excluded from the meta-analysis. Emission factors and pollutant concentrations used in our analyses are shown in Tables S4 and S5.

We computed study-specific proportions of respiratory symptoms and diseases or the mean (and standard deviation) of lung function values for each study among BKWs and control groups or for participants living near and far away from the brick kilns. For lung function data, we provided study-specific means and absolute mean differences in lung function. We calculated an overall mean difference for lung function values between BKWs and controls. We used a fixed effects meta-analysis to summarize mean differences across studies. We also summarized lung function data stratified by both BKW exposure and tobacco smoking status and provided mean percent predicted values for both BKWs and controls. We calculated the average proportion of respiratory symptoms and disease for BKWs using the reciprocal of variances as weights. We also calculated risk differences (i.e., the absolute difference in proportions) of respiratory symptoms between BKWs and controls or for participants living near and far away from the brick kilns using a fixed effects meta-analysis.

Analyses were conducted in R version 4.2.1 alias Funny-Looking Kid.^17^

## RESULTS

### Literature search

We identified 1089 references through electronic searching and 6 through manual searching. After removing duplicates, screening of titles and abstracts, and evaluation of full-text articles, we identified 104 studies for review (Figure 1). Fifty-eight (56%) reported data on brick kiln pollution,^6,8,15,16,18–68^ 25 (24%) on health,^5,69–93^ and 21 (20%) on both pollution and health.^7,9,94–111^ We mapped countries where brick kiln research was conducted in Figure 2. Most were in South Asia (Online Supplement).

**Figure 1.**
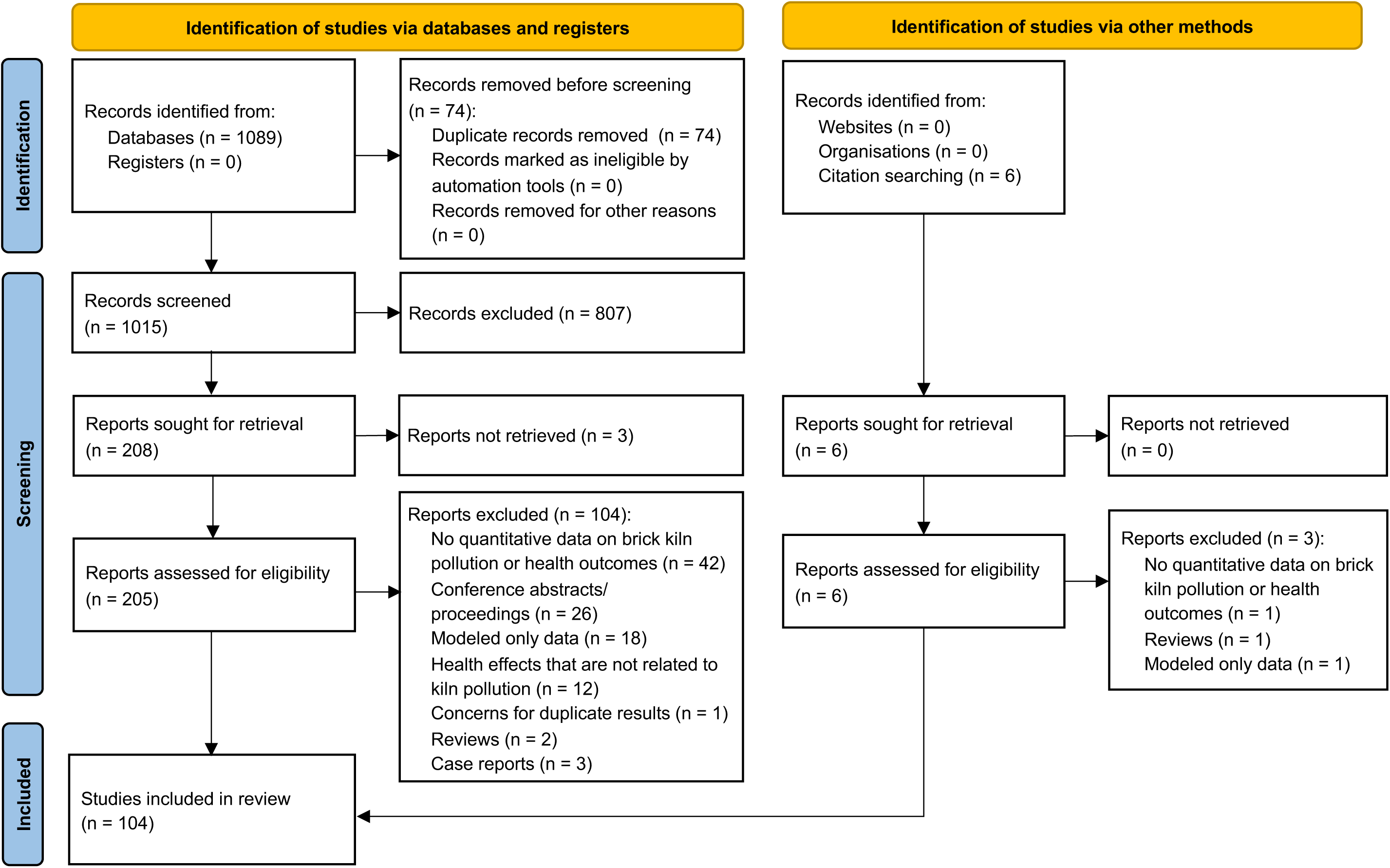
Selection process for identifying original articles for the systematic review of brick kiln pollution and its impact on human health.

**Figure 2.**
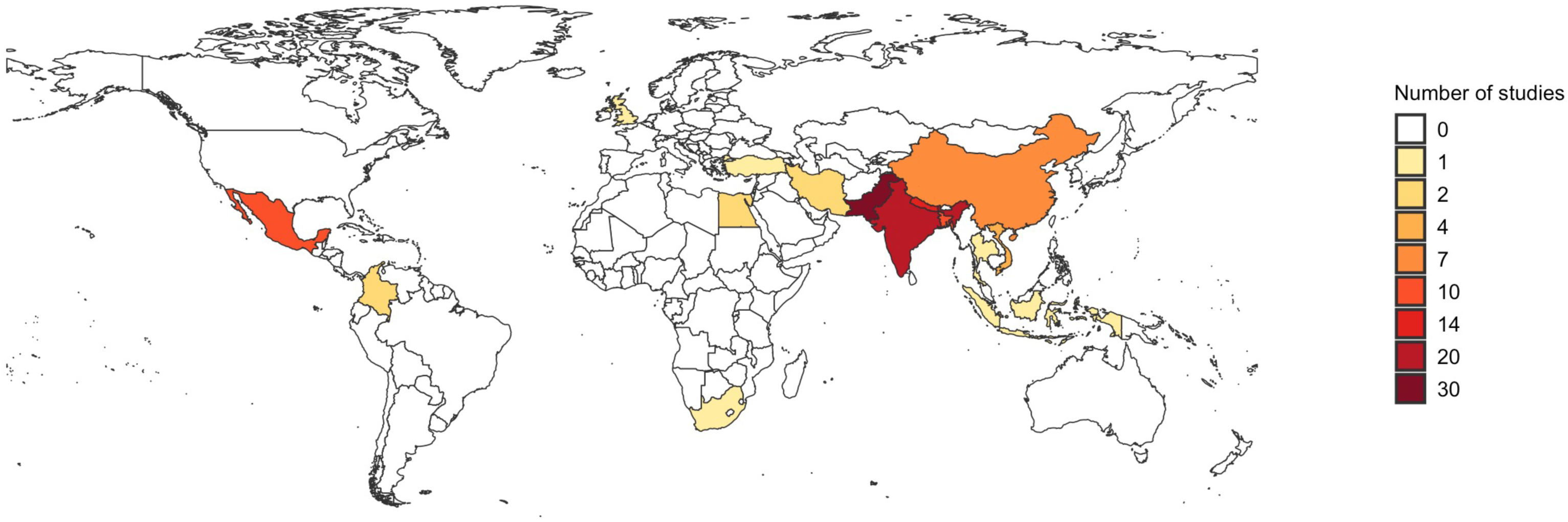
Number of studies per country included in this systematic review.

### Brick kiln pollution

Seventy-nine studies reported data on brick kiln pollutants or toxic exposures (Table S1). Overall study quality was low (Table S2). We report on study characteristics in Table S1 and summarize type of pollutant data, pollutants measured and measurement locations in Table S3. The most measured pollutants were PM, SO_2_ and CO, and most studies performed measurements at kiln sites (n=65, 82%), including flue gas and in-stack sampling, samples collected at various locations within the kiln site, and measurements of personal exposures and exposure biomarkers in BKWs and children living at the site (Table S3).

#### Pollutant emission factors from brick kilns

PM and NOx were the most reported emission factors (Online Supplement). Emission factors for PM, BC, EC, and OC were higher for traditional when compared to improved brick kilns (Figure 3). Emissions of OC were considerably larger than EC across all studies (Table S4).

**Figure 3.**
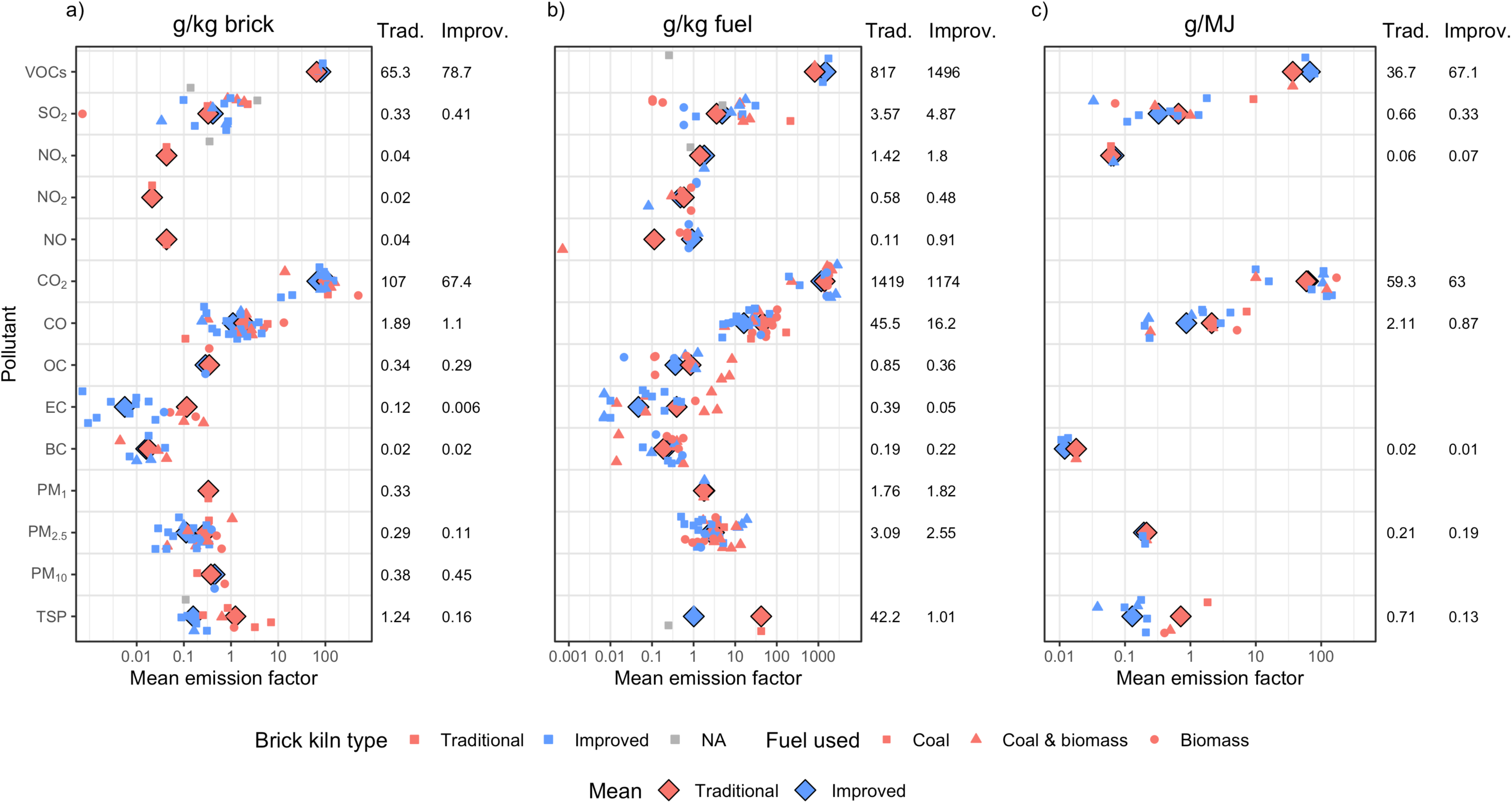
Mean emission factors in (a) g/kg brick, (b) g/kg fuel, and (c) g/MJ. We plot emission factors for multiple pollutants, categorized by brick kiln design and fuel used. Symbols in red and blue represent kiln types categorized as traditional and improved, respectively. Symbols in grey represent kilns whose type was not specified. Brick kilns that used coal only are shown as squares, those that used coal and biomass are shown as triangles, and those that used biomass only are shown as circles. Means for traditional and improved kiln designs are shown in the red and blue diamonds, respectively. Values displayed on the right of each plot represent the mean emission factors ± standard deviations for traditional vs improved kiln designs, weighted by sample size.

Among gaseous pollutants, CO_2_ was the largest emission and was similar in both traditional and improved kilns (Figure 3). Emission factors of NO, NO_2_ and NO_x_ were low across both traditional and improved kilns. Emission factors of SO_2_ displayed a large variability across studies. In general, brick kilns using biomass had lower SO_2_ emission factors than those using coal. As expected, due to the lower combustion efficiency of traditional kilns, CO emission factors were higher among traditional vs improved kilns (mean MCEs were 0.96 ± 0.02 and 0.98 ± 0.01 for traditional and improved kilns, respectively). This trend held true when further stratifying by fuel type (Tables S7 – S9). CO emission factors decreased with increasing MCE (Figure S1).

We identified five studies that reported various VOC emission factors,^31,36,37,56,65^ and found that VOCs were lower in brick kilns than in food or pharmaceutical factories. Some studies reported higher VOC emissions in improved kilns when compared to traditional kilns; however, VOC emissions depend not only on kiln design but also on fuel type.^36^ We summarize findings on VOC emissions from all five studies in the Online Supplement.

Three studies reported PAH emission factors.^16,34,41^ Emission factors of PAHs and other incomplete combustion products were lower in stack gas than fugitive emissions due to the longer reaction time along the chimney.^16^ The most abundant PAHs from a clamp kiln were chrysene, benz(a)anthracene, benzo(e)pyrene, and 1-methylcrysene.^41^ Very low levels of PAHs were observed from a zigzag kiln, reflecting relatively complete combustion of the coal.^34,41^ We summarize findings from each study in the Online Supplement.

#### Pollutant concentrations at brick kiln sites

We report on the types of pollutant concentrations measured at the brick kiln sites in Table S9. Traditional kilns emitted higher concentrations of PM, BC, EC, OC and CO, similar concentrations of VOCs and SO_2_, and lower concentrations of NO_x_ when compared to improved brick kilns (Figure 4). High concentrations of VOCs, CO_2_, CO and SO_2_ were measured in-stack among both traditional and improved kilns (Figure 4a). Mean out-of-stack concentrations of CO, SO_2_ and NO_2_ in traditional kilns were above the WHO recommended limits for 24-hour mean exposures but below the National Institute for Occupational Safety and Health (NIOSH) time-weighted average recommended exposure limit for up to a 10-hour workday (Figure 4b). Mean PM_10_ and PM_2.5_ concentrations from traditional kilns were both higher than the WHO limits, whereas mean PM_2.5_ from improved kilns was below the recommended limit.

**Figure 4.**
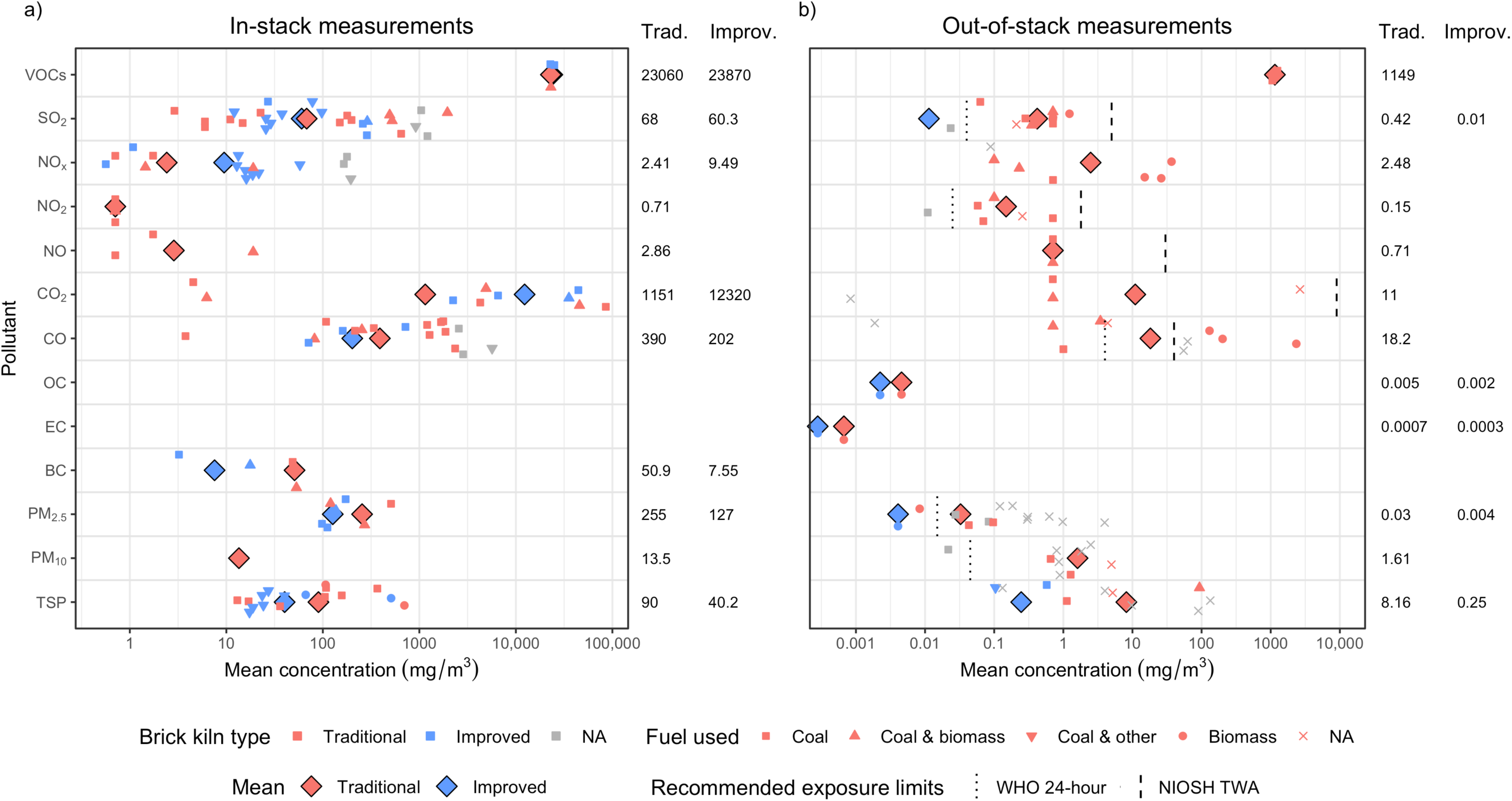
Mean pollutant concentrations from (a) in-stack measurements, and (b) brick kiln site measurements. Symbols in red and blue represent kiln types categorized as traditional and improved, respectively. Symbols in grey represent kilns whose type was not specified. Brick kilns that used coal only are shown as squares, those that used coal and biomass are shown as upward-pointing triangles, those that used coal and solid waste are shown as downward-pointing triangles, and those that used biomass only are shown as circles. Means for traditional and improved kiln designs are shown in the red and blue diamonds, respectively. Values displayed on the right of each plot represent the mean pollutant concentrations ± standard deviations for traditional vs improved kiln designs, weighted by sample size.

We identified six studies reporting various PAH concentrations.^30,39,40,54,62,104^ We summarized findings of the individual studies in the Online Supplement. Briefly, we identified that the type and concentration of PAH depends on type of fuel used and type of sample (soot or soil). The most common PAHs identified were acenaphtylene, phenanthrene, chrysene, and fluorene.

Six studies reported metal concentrations.^30,47,49,52,61,96^ We summarized findings of the individual studies in the Online Supplement. Briefly, heavy metals such as mercury, lead, barium, zinc, chromium and cadmium were commonly identified at the kiln sites. Most studies reported that heavy metal concentrations per gram of sample tested exceeded background levels or national or international regulatory levels.

Surprisingly, only one study reported silica concentrations.^8^ Beard et al.^8^ measured pollutant concentrations, including respirable silica, inside and outside 16 BKW homes across four brick kilns in Bhaktaphur, Nepal. The geometric mean silica (quartz) concentration across all 32 samples collected was 6.22 μg/m^3^, while concentrations of silica in the form of cristobalite and tridymite were all below the limit of detection. These results suggest that silica concentrations are typically low at on-site worker housing and that silica exposures among BKWs are likely to occur predominantly during work hours.

#### Contribution of brick kiln emissions to surrounding ambient air pollution

Fourteen studies reported the contribution of brick kiln emissions to the surrounding ambient air pollution in South Asia (Table S3). We report on data from individual studies in the Online Supplement. Brick kilns appear to contribute to ambient PM_2.5_, PM_10_ and BC anywhere in the range of 20%-41%, up to 28% and 6%-91%, respectively, depending on the location and model used. ^24–29,35,112,113^ Brick kilns are also primary contributors to EC (40%)^44^ and NMVOCs (10%)^55^ in some settings. Source apportionment studies in and adjacent to the Kathmandu valley, Nepal, showed that coal combustion in brick kilns accounted for 9-17% of the organic material in ambient PM_1_. ^112,113^

In India and Nepal, ambient BC, PM_2.5,_ PM_10_ and TSP were reported to be 70-180% higher in the winter season when the kilns are operational compared to the monsoon season when kilns are not operational.^22,103^ Sites near kilns are also associated with higher levels of ambient pollution. In Pakistan, for example, ambient PM_10_, SO_2_ and NO_2_ was 500%, 28% and 270% higher, respectively, in an area where brick kilns were operational compared to an area where kilns were not operational.^95^ Similarly, another study in Pakistan found that PM_2.5_ and PM_10_ concentrations were 4.6 times higher within 3 km of kilns compared to >3 km from the kilns.^7^

#### Personal exposures of brick kiln workers

There were 6 (8%) studies that reported personal exposures to pollutants among BKWs (Table S3). Data collected for BKWs across five different job types, or similar exposure groups (SEGs), in 16 brick kilns in Kathmandu Valley, Nepal, revealed that average exposures to total suspended particles (TSP), respirable suspended particles (RSP) and silica were highest among red brick loaders (Figure 5).^14,94^ Green brick molders had the lowest exposures to RSP and silica, while TSP exposures were lowest among firemen. On average, exposures to respirable silica exceeded the NIOSH recommended exposure limit of 50 μg/m^3^ in all job types, with mean exposures 1.4 to 6.6 times higher than the limit.

**Figure 5.**
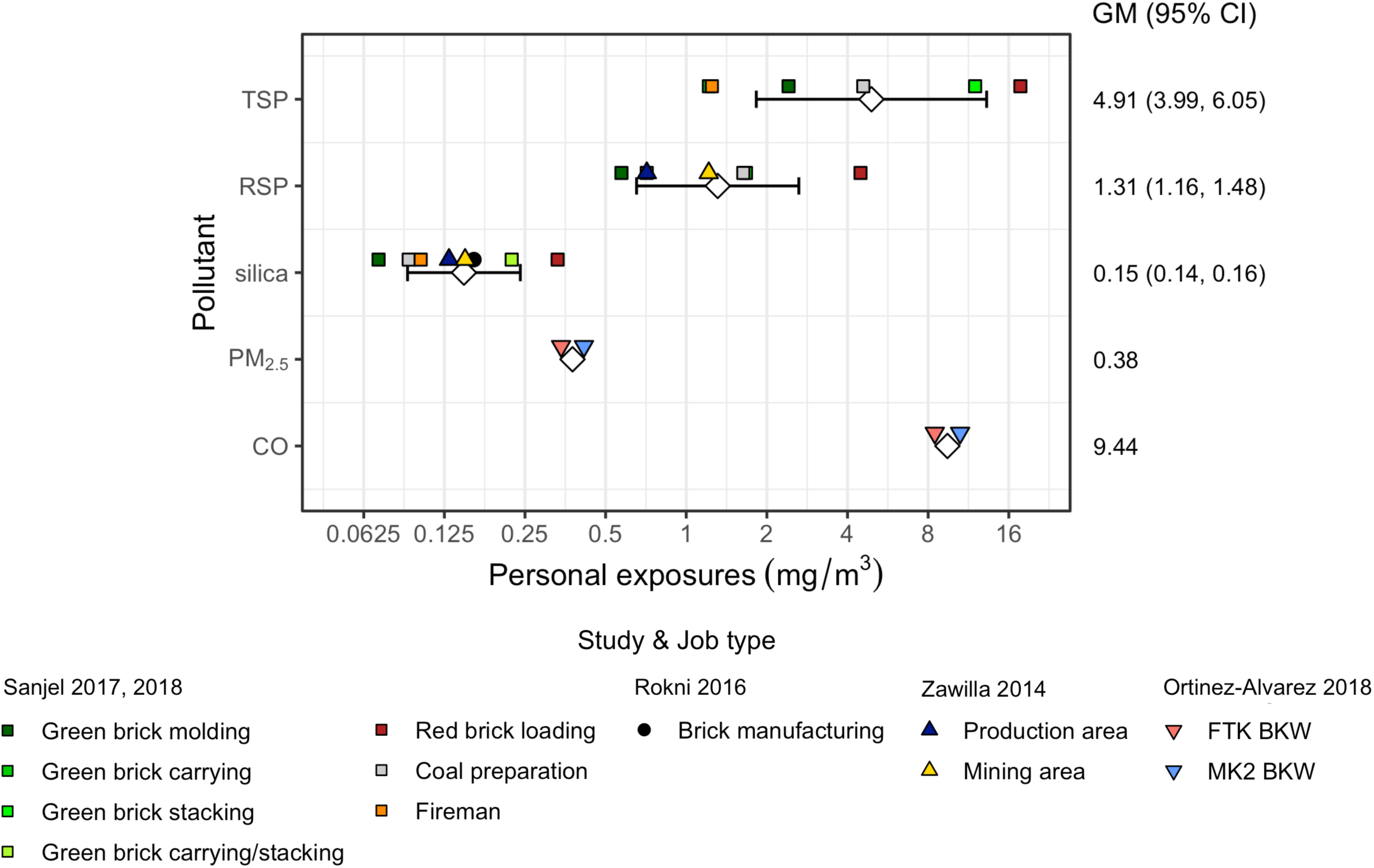
Geometric means and 95% confidence intervals of personal exposures by brick kiln job type. Diamonds and error bars represent the pooled geometric means and 95% confidence intervals, respectively, for each pollutant. Values displayed on the right represent pooled geometric means and 95% confidence intervals for each pollutant.

Similar levels of silica exposure were observed in a study among 12 workers across 2 brick kilns in the Mazandaran province in Iran (Figure 5).^53^ In brick and tile works in England and Scotland, mean quartz concentrations across 23 different work groups ranged between 40 μg/m^3^ for those with no direct exposure (office, canteen), to 620 μg/m^3^ for kiln demolition workers (SDs missing so not included in meta-analysis).^106^ Exposures to RSP were similar to those reported by Sanjel et al.^94^, with mean values ranging between 0.4 mg/m^3^ and 10 mg/m^3^ . Personal samples collected in a brick factory in Southern Cairo, Egypt, showed exposures were higher in the mining area than in the production area for both RSP and silica.^111^

Another study conducted in Durango, Mexico measured personal exposures to PM_2.5_ and CO among workers in one fixed traditional kiln (n=2) and one MK2 Marquez kiln (n=2).^48^ The assessments were conducted on the worker responsible for feeding the kilns and the assistant to the main operator. Despite the more energy-efficient design of the MK2 and lower emission concentrations, average personal exposures to PM_2.5_ and CO were higher in the MK2 vs the FTK (Figure 5). This was attributed to the presence of a flat shade roof and observational portholes, as well as mismanagement of the operational processes in the MK2, resulting in an accumulation of pollutants.

#### Exposure biomarkers

We identified 13 (17%) studies that measured exposure biomarkers among BKWs, children at kiln sites, and children living in communities near kilns (Table S3). BKWs and children near kilns had higher urinary concentrations of PAH exposure biomarkers (1-hydroxypyrene, α-naphthol, β-naphthol),^33,46,50,105^ and benzene exposure biomarkers (trans,trans-muconic acid)^33^ compared to controls or even individuals living near heavy traffic, waste landfill or metallurgical industry (Table S10). Heavy metal concentrations in blood among BKWs and nonworkers living near kilns were also higher compared to controls,^9,57,97,101,102^ but not compared to those in other urban marginalized communities.^33^

### Health outcomes

Forty-six studies evaluated health outcomes (Table S11). The most evaluated conditions were respiratory health, biomarkers, and musculoskeletal disorders. Overall study quality across health outcomes was low (Table S12).

#### Respiratory health

We summarized characteristics of studies reporting respiratory health data in the Online Supplement and Table S13. Forced expiratory volumes (FEVs) were lower in BKWs than in controls (Figure 6). Tobacco smoking may be an important effect modifier: BKWs who were smokers had lower FEVs when compared to non-smoking BKWs, non-BKWs smokers, or non-smoking non-BKWs (Table 1); however, this interaction was not formally evaluated by any of the studies and missing information on sample size in subgroups for most studies did not allow us to conduct a meta-analysis. Reported mean percent predicted values of FEV were lower in BKWs than in controls (Table 2) but there were too few studies to conduct a meta-analysis.

**Figure 6.**
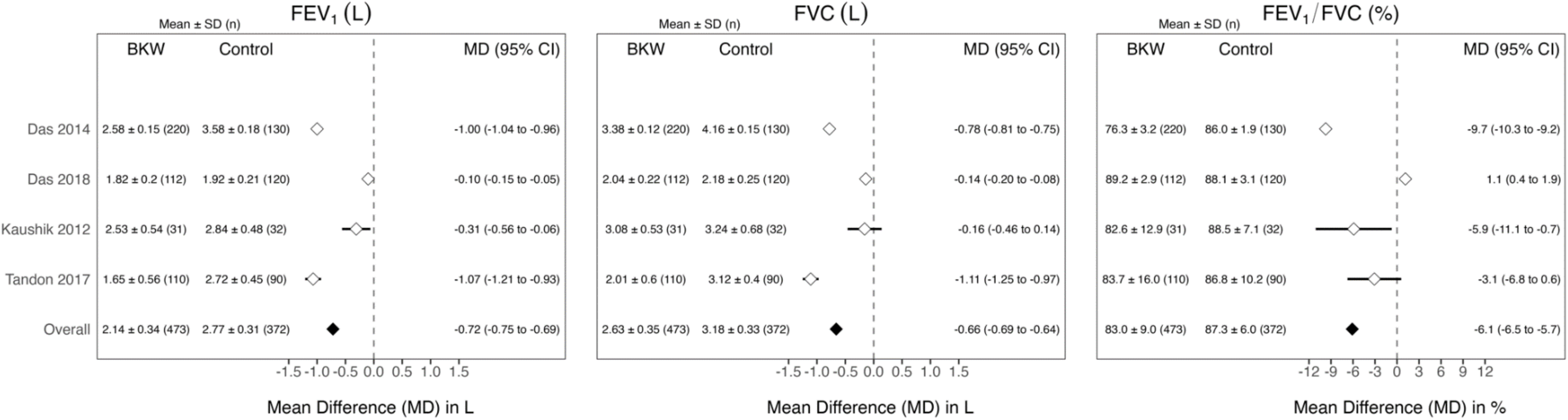
Mean values of lung function for brick kiln workers and controls, and mean difference between brick kiln workers and controls.

**Table 1.** Mean values of lung function for brick kiln workers and controls stratified by smoking status, and mean difference between brick kiln workers and controls stratified by smoking status.

| | Mean ± SD (n) | | | |
| --- | --- | --- | --- | --- |
|  | Brick kiln workers |  | Reference (or control) group |  |
|  | Smokers | Non-smokers | Smokers | Non-smokers |
| **FEV<sub>1</sub> (in L)** |  |  |  |  |
| Goel 2015<sup>76</sup> | 2.15 ± 0.50 (N/A) | N/A | 2.75 ± 0.73 (N/A) | 3.53 ± 0.24 (N/A) |
| Raza 2021<sup>108</sup> | 1.98 ± 0.53 (25) | 2.16 ± 0.62 (35) | N/A | N/A |
| Tandon 2017<sup>114</sup> | N/A | 1.65 ± 0.56 (110) | N/A | N/A |
| **FVC (in L)** |  |  |  |  |
| Goel 2015<sup>76</sup> | 2.56 ± 0.66 (N/A) | N/A | 3.17 ± 0.69 (N/A) | 3.86 ± 0.25 (N/A) |
| Raza 2021<sup>108</sup> | 2.27 ± 0.67 (25) | 2.53 ± 0.74 (35) | N/A | N/A |
| Tandon 2017<sup>114</sup> | N/A | 2.01 ± 0.60 (110) | N/A | N/A |
| **FEV<sub>1</sub>/FVC (in %)** |  |  |  |  |
| Goel 2015<sup>76</sup> | 82.8 ± 8.5 (N/A) | N/A | 85.9 ± 7.5 (N/A) | 91.4 ± 3.6 (N/A) |
| Srivastava 2002<sup>91</sup> | 74.2 ± 8.8 (47) | 70.4 ± 6.2 (23) | 79.4 ± 6.8 (52) | 78.6 ± 5.1 (54) |
| Tandon 2017<sup>114</sup> | N/A | 83.69 ± 16.02 (110) | N/A | N/A |

**Table 2.** Mean percent predicted values of lung function for brick kiln workers and controls, and mean difference between brick kiln workers and controls.

| **Lung function value** | **Brick kiln worker** | **Reference (or control) group** | **Mean difference (95% CI)** |
| --- | --- | --- | --- |
| **FEV<sub>1</sub> percent predicted** |  |  |  |
| Sheta 2015<sup>88</sup> | 74.1 ± 7.8 (173) | 81.8 ± 10.1 (170) | -7.7 (-9.6 to -5.8) |
| Srivastava 2002<sup>91</sup> (non-smokers) | N/A | N/A | N/A |
| Srivastava 2002<sup>91</sup> (smokers) | N/A | N/A | N/A |
| Tandon 2017<sup>114</sup> | 86.5 ± 12.0 (110) | 93.0 ± 8.5 (90) | -6.5 (-9.4 to -3.6) |
| **FVC percent predicted** |  |  |  |
| Sheta 2015<sup>88</sup> | 75.9 ± 6.7 (173) | 80.3 ± 6.1 (170) | -4.4 (-5.8 to -3.0) |
| Srivastava 2002<sup>91</sup> (non-smokers) | 81.2 ± 4.6 (52) | 83.6 ± 3.8 (54) | -2.4 (-4.0 to -0.8) |
| Srivastava 2002<sup>91</sup> (smokers) | 79.2 ± 8.8 (47) | 81.1 ± 4.8 (23) | -1.9 (-5.1 to 1.3) |
| Tandon 2017<sup>114</sup> | 88.9 ± 9.3 (110) | 94.4 ± 8.9 (90) | -5.5 (-8.0 to -3.0) |
| **FEV<sub>1</sub>/FVC percent predicted** |  |  |  |
| Sheta 2015<sup>88</sup> | N/A | N/A | N/A |
| Srivastava 2002<sup>91</sup> (non-smokers) | 79.4 ± 6.8 (52) | 78.6 ± 5.1 (54) | 0.8 (-1.5 to 3.1) |
| Srivastava 2002<sup>91</sup> (smokers) | 74.2 ± 8.8 (47) | 70.4 ± 6.2 (23) | 3.8 (0.2 to 7.4) |
| Tandon 2017<sup>114</sup> | 80.7 ± 4.8 (110) | 85.2 ± 5.0 (90) | -4.5 (-5.9 to -3.1) |

The prevalence of respiratory symptoms was higher in BKWs than in controls and was higher in participants living near kilns than in those living farther away from the kilns (Table 3). Both chronic diseases, like asthma, COPD and chronic bronchitis, and respiratory infections were also more common in BKWs when compared to controls (Table 3). One study found higher odds of tonsillitis and throat inflammation in children living near brick kilns compared to those living father away from bricks kilns.^103^ Another study found a higher incidence of pneumonia and upper respiratory infections when brick kilns was the source category of fine particulate matter.^87^ A study in India reported that 9.4% of adult and child BKWs had chest symptoms consisting of productive cough for two weeks or longer with or without chest pain, intermittent fever or hemoptysis.

**Table 3.**
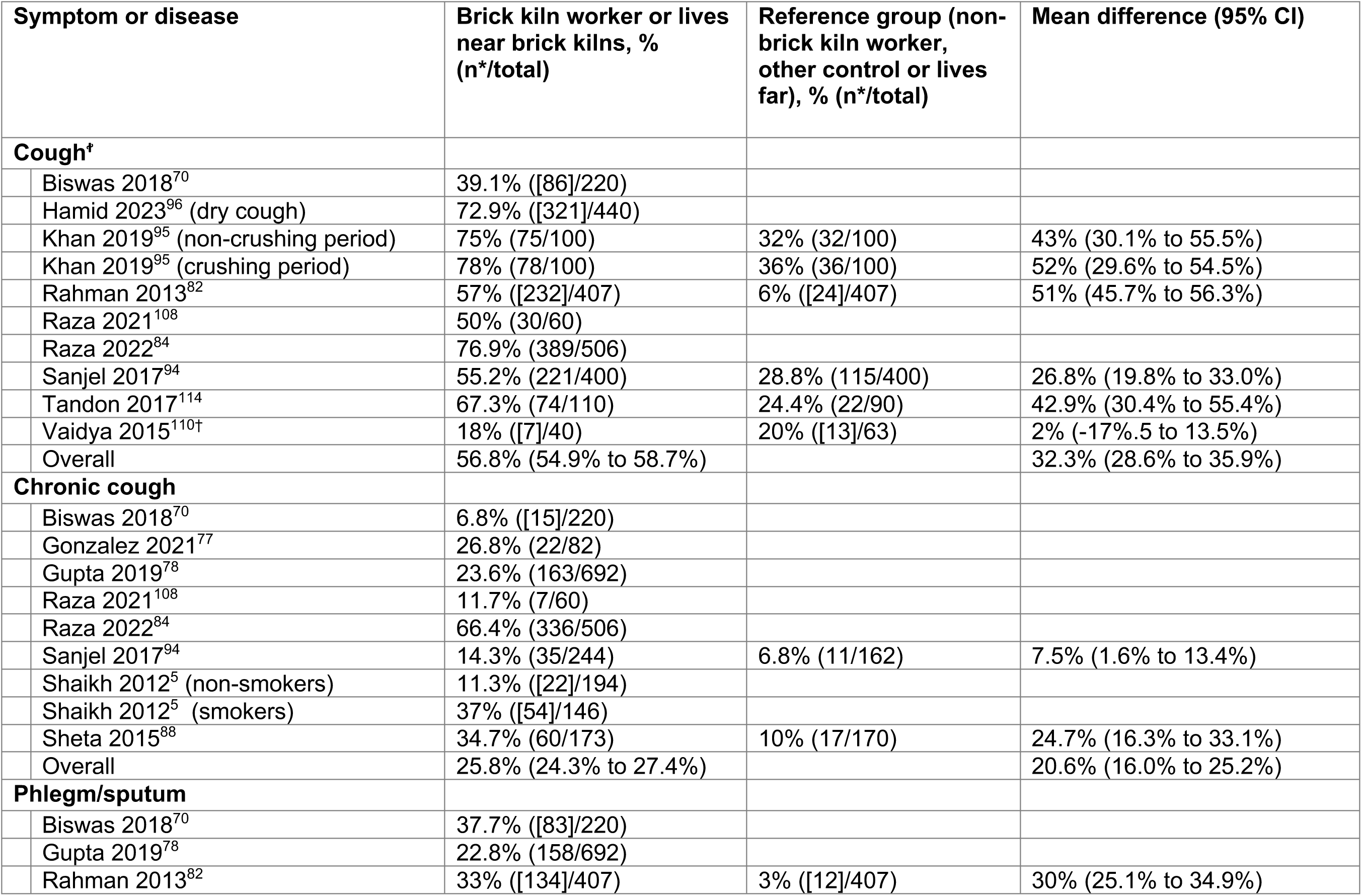
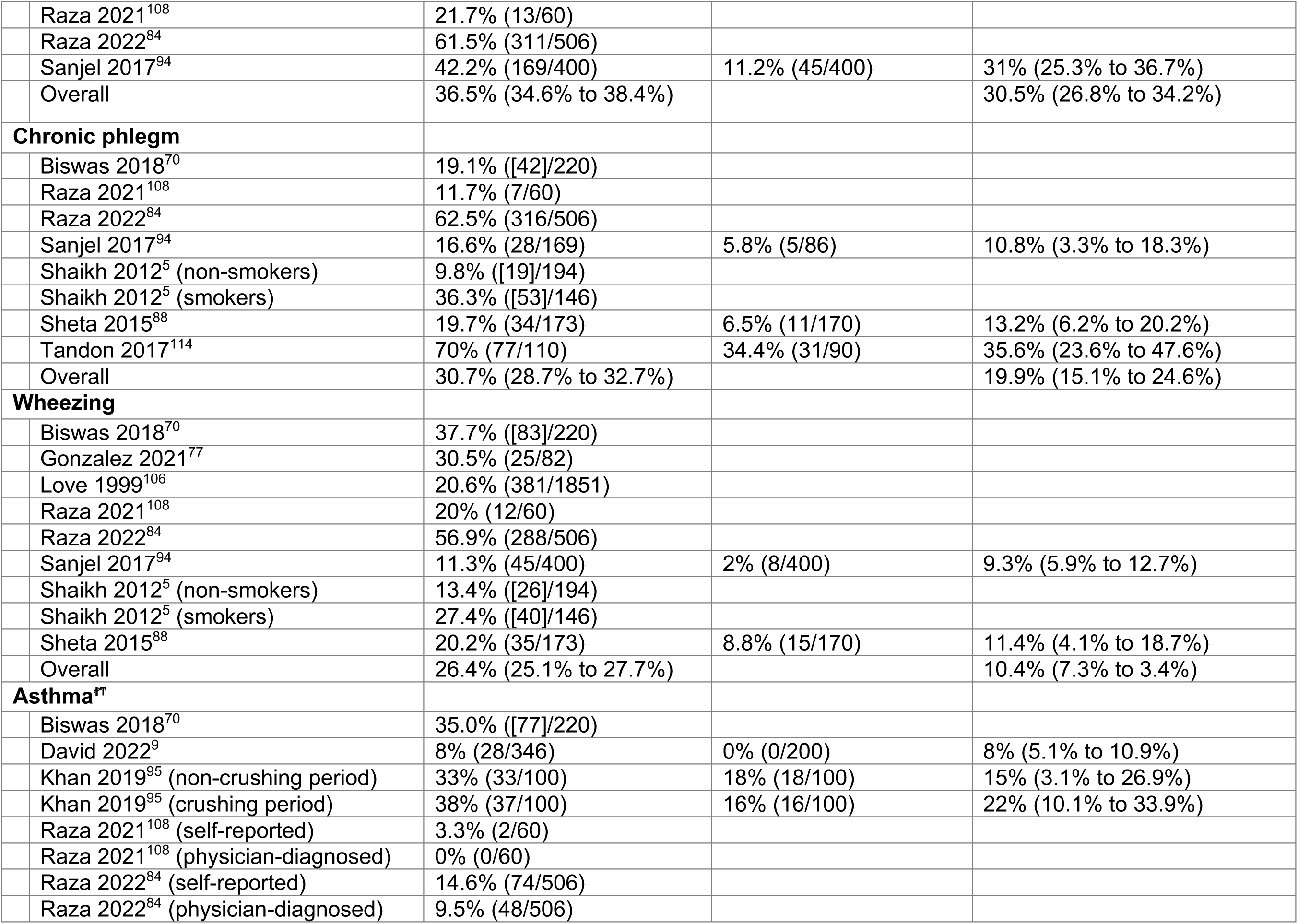
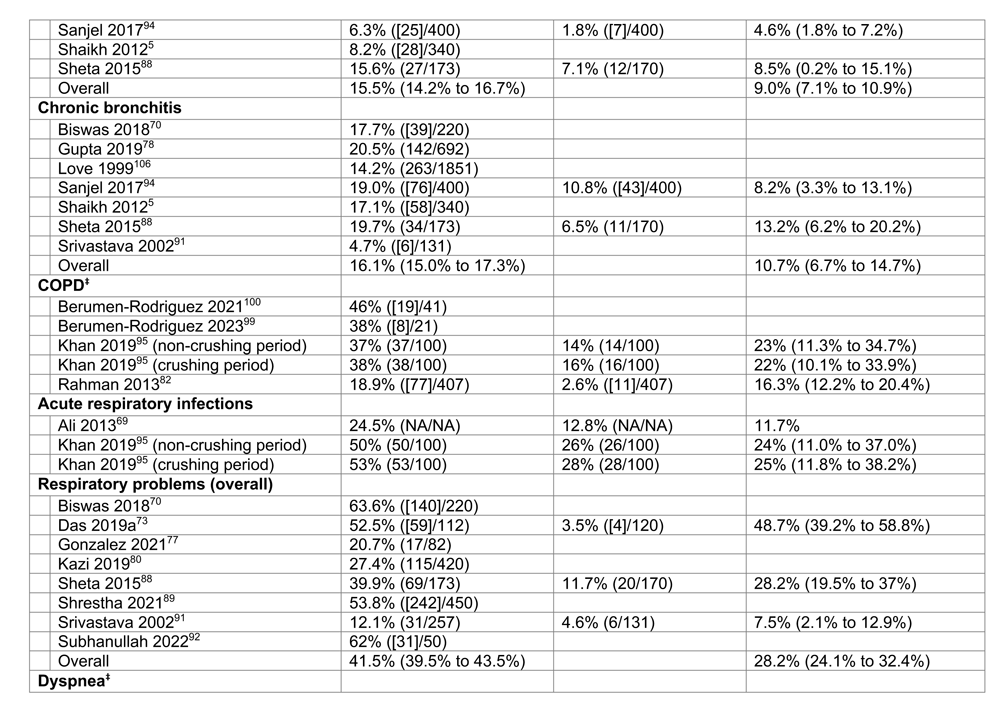
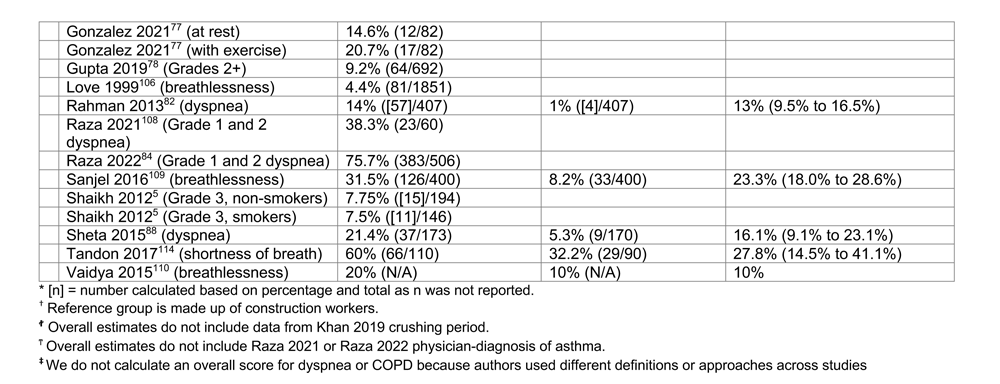
Prevalence of respiratory symptoms or chronic respiratory disease in brick kiln workers or controls (or participants living near or far from kilns) and mean difference between brick kilns workers and controls.

Ten studies evaluated the association between respiratory outcomes and number of years worked (Table S13): one study reported a positive correlation between the score obtained from a screening questionnaire for COPD and number of years worked (numerical value for correlation not given);^100^ one study found a higher prevalence of small opacities on chest X-ray (≥0/1) and either number of years worked or estimated cumulative exposure to respirable quartz;^106^ four studies did not find any relationship between respiratory symptoms or lung function and number of years worked;^70,79,89,107^ four studies found either a higher prevalence of respiratory symptoms or lower lung function with more years of work exposure.^5,80,88,114^ Ten studies evaluated the association between respiratory outcomes and BKW occupation: three found more respiratory symptoms (including conditions like chronic bronchitis and asthma) in bakers when compared to other BKW types;^5,77,88^ four did not find an association between lung function or respiratory symptoms and BKW type,^89,94,107,108^ one found that among workers in the modulation, loading, burning and unloading sites, obstructive and restrictive impairments were highest in workers at the loading and burning sites, respectively,^108^ one found that respiratory symptoms were lower in molders compared to carriers, stackers or firemen,^70^ and one found more respiratory symptoms in bigaaris (translates to carriers) when compared to other brick kiln types.^80^

#### Gastrointestinal disease

We identified 3 studies reporting data on gastrointestinal problems among BKWs,^80,91,111^ but only 1 related to kiln exposures.^111^ Zawilla et al.^111^ examined liver function among 87 silica-exposed BKWs and 45 non-exposed controls in Egypt. Mean liver function test concentrations (except albumin and bilirubin), MMP-9, and immunoglobulins (G and E) were all significantly higher in BKWs compared to controls; more than half of BKWs had abnormal AST and GGT and almost 20% had abnormal ALT and ALP levels, but no comparative data was reported for controls. The other studies, conducted among different BKW groups in India, reported proportions between 4% and 13.2% with gastrointestinal disorders but it was unclear if these were attributed to brick kilns or poor sanitary facilities and polluted drinking water.^80,91^

#### Reproductive health risks

Three studies reported data on reproductive health in BKWs.^9,74,102^ One study conducted in Pakistan compared health risks in 232 female BKWs to 113 controls and found a lower age at menarche for BKWs when compared to controls (11.1 ± 0.16 years vs. 14.1 ± 0.19 years, respectively).^74^ Female BKWs had a higher average number of pregnancies, abortions and stillbirths when compared to controls; however, controls were younger and fewer were married than BKWs. Another study in Pakistan examined testosterone and luteinizing hormone (LH) levels in 110 male BKWs with at least 10 years of work experience, 30 non-BKWs aged 19-45 years living in nearby communities to the kilns, and 57 adult males aged 21-40 years (distance from kilns not reported).^102^ The authors found that testosterone in BKWs was lower when compared to controls or non-BKWs who lived near the kilns. Testosterone levels were also lower in brick bakers compared to other BKW occupations. LH was lower in brick makers and brick carriers compared to controls or nonworkers who lived near the kilns. Brick bakers appear to have higher LH than controls and nonworkers who lived near the kilns, although the text and table report this finding inconsistently. The last study, also from Pakistan, compared 346 adult BKWs to 200 non-BKW controls of a similar age range.^9^ The authors found higher FSH and LH and lower testosterone in BKWs compared to controls

#### Musculoskeletal disorders

Nine studies reported data on musculoskeletal pain;^70–73,80,89,91,96,110^ however, none of these studies linked musculoskeletal pain directly to brick kiln pollution exposure. Four studies compared BKWs to controls and found that the prevalence of musculoskeletal problems was consistently higher in BKWs than in controls.^71,73,91,110^

#### Cardiovascular disease

We did not identify any papers reporting prevalence of cardiovascular disease among BKWs or any associations of cardiovascular outcomes with kiln pollutants. One study reported a prevalence of hypertension at 25.5% among Mexican BKWs,^100^ and two studies reported heart rate and blood pressure before and after completion of work in the brick kiln as an assessment of physiological stress.^71,73^ Comparing 220 male brick field workers from 12 brick fields in West Bengal, India and 130 controls engaged in office work, Das found no difference in resting heart rate between groups.^71^ Resting blood pressure was lower among BKWs although results presented in the table and text were inconsistent. Just after work, both heart rate and blood pressure were higher in BKWs than controls. In a later study among 112 child BKWs aged 9 to 16 years and 120 controls engaged in household jobs, the author found no difference in heart rate or blood pressure at rest between the two groups, but significantly higher values among BKWs than controls after completion of work.^73^ A total of 46% of kiln workers reported cardiovascular problems compared to 4% of controls.

#### Cancer

We identified 4 papers reporting cancer risk due to exposure to kiln emissions, namely PAHs, metals, and radionucleotides.^20,104,115,116^ However, all 4 of these studies were excluded from the health outcomes review as the risk assessment was based on models rather than collected health outcome data. In addition, one of the studies modelled the risk based on PAH concentrations in an agricultural and brick production area rather than at a kiln site.^115^

#### Biomarkers

Eleven studies evaluated biomarkers in BKWs against unexposed controls (Table S11). We summarized the findings for each study in the Online Supplement. Overall, there was evidence of higher reactive oxygen species, lower concentrations of superoxide dismutase and catalyse, and more DNA damage in BKWs when compared to controls (Table S14). There was also evidence of more inflammation as evidenced by a higher C-reactive protein and cortisol in BKWs when compared to controls, and elevated cytokine levels in exhaled breath condensate of BKWs. Two studies identified down-regulated expression of genes associated with DNA and protein repair in BKWs when compared to controls.

#### Linear growth

Three studies examined the role of brick kiln pollution on child linear growth (e.g., stunting).^7,85,90^ Nasir et al.^7^ examined 383 children aged 5-12 years from Pakistan, measured their height and calculated Z-scores using the 2007 World Health Organization (WHO) international reference. Mean height-for-age Z-score was -0.50 (prevalence of stunting not reported). The authors used propensity score matching on a probit regression using 4 different techniques to match on household and child characteristics for children living within 3 km of a brick kiln (exposed) when compared to children living farther away (controls). Using nearest neighbor and radius matching, exposed children had a Z-score that was -0.68 and -0.6 lower than controls, respectively (p<0.01). Z-scores were -0.43 and -0.42 lower with kernel and stratification matching, albeit not statistically significant. Roshania et al.^85^ conducted a large cross-sectional study with a cluster design and measured height and in weight in 2564 migrant children (aged 0-11 months and 12-23 months) from 1156 brick kilns in India. Z-scores were calculated using the 2006 WHO Multi Growth Reference studies. Overall prevalence of stunting was 51.6% (mean Z-score not reported). Among children whose first episode of migration to a kiln occurred before age 6 months, the odds of stunting were 1.6 (95% CI 1.17 – 2.19) and 2.1 (1.30 – 3.41) times higher in those with two and three migration episodes, respectively, when compared to one. Sinaga et al.^90^ evaluated 192 children from Indonesia aged 0-24 months who lived in villages with (n=101) and without (n=91) clay brick kilns. The overall prevalence of stunting was 19%. The authors did not find a difference in the prevalence of stunting between children who lived in villages with clay brick kilns and those who lived in villages without clay brick kilns (19.8% vs. 18.6%; p=0.45).

#### Other health outcomes

Other health outcomes reported in the literature included skin complaints,^73,75,80,89,91,92,109,110^ ocular problems,^73,80,89,91,92,96,109,110^ injuries,^80,109^ and nasal and otologic complications.^75^ The proportion of BKWs with skin complaints ranged between 5%^110^ and 19%^80^, with the major problems being callosities, eczema, dermatitis and itchy hands and feet.^80,91^ Results on eye problems varied widely, with ∼4% of BKWs reporting eye complaints in 2 studies,^80,91^ and 48% reporting problems in vision in another.^110^ Interestingly, a comparative study between BKWs and grocery workers as controls, found that the proportion experiencing eye problems was significantly lower among BKWs (14.8%) than controls (22.5%).^109^ On the other hand, the occurrence of injuries was significantly higher among BKWs (55.0%) than controls (44.2%). Among the 420 BKWs interviewed by Kazi et al.^80^ however, only 7% reported injuries. Lastly, an otorhinolaryngologic evaluation of 103 BKWs in Erbaa, Turkey, showed that 1.9% had structural otologic complications, whereas general otologic complications, including dust in the external ear canal or tympanic membrane, were observed in 25.2% of workers.^75^ Structural and general nasal complications were observed in 26.2% and 68.0% of BKWs, respectively, and were significantly higher among workers who did not wear a mask and those who had worked for more than 10 years when compared to those who did not wear masks or worked fewer than 10 years, respectively. The authors did not find differences in otologic or rhinologic complications by BKW occupation.

### Interventions

We identified 4 studies that compared different brick kiln technologies and provided recommendations for reducing emissions.^6,15,36,48^ A study which compared emission concentrations from a fixed traditional kiln and an improved MK2 Marquez kiln in Durango, Mexico, showed that average emissions of PM_2.5_, OC and EC from the FTK were a factor of two higher than the MK2, despite initial mismanagement by the MK2 operator which led to higher concentrations in the MK2 during the first sampling cycle.^48^ The authors highlighted the need for supervision by authorities, training, and good practice on implementation and operation to maximize the environmental benefits of improved technologies.

Among 18 brick kilns in Greater Dhaka, Bangladesh, consisting of FCKs, ZZKs and HKs, Haque et al. found average fuel-based emission factors of BC, PM_2.5_ and CO were highest in FCKs, whereas CO_2_ and SO_2_ emissions were highest in ZZKs and HKs, respectively.^36^ They noted, however, that SO_2_ concentrations not only depend on kiln technology but also on the sulfur content in the fuel used. Fuel consumption per fired brick was highest for FCKs. Similar results were observed in a comparative study conducted across 4 FCKs and 3 ZZKs in Kathmandu Valley, Nepal.^6^ Compared with ZZKs, FCKs had higher PM_2.5_ and BC, comparable SO_2_, and lower CO_2_ emissions based on fuel-based emission factors. Emissions per kilogram of fired brick were lower in ZZKs than FCKs for all pollutants, and brick production capacity was higher in ZZKs. Their results suggested that a conversion from FCKs to ZZKs could result in emission reductions in PM_2.5_ by ∼20% and BC by ∼30% per kilogram of fuel used, and ∼40% for PM_2.5_ and ∼55% for BC per kilogram of fired brick. Another study conducted in Punjab, Pakistan, found fuel-based emission factors were substantially higher in FCKs than ZZKs for all pollutants measured (SO_2_, CO, CO_2_, PM), except for NO_x_.^15^ In this case, emission reductions per kilogram of fuel used in ZZKs compared to BTKs were 98% for PM, 96% for SO_2_, 83% for CO, and 64% for CO_2_. While the emission factors and emission reduction estimates varied significantly across these studies, the results suggest that converting from FCKs to ZZKs or HKs, along with the use of high-grade coal with lower sulfur content, could be an effective solution for reducing emissions from brick kilns. The higher combustion efficiency of ZZKs also translates into higher private net benefits for ZZK owners, providing an incentive to shift from BTKs to ZZKs.^15^

An additional 2 studies offered similar recommendations to reduce emissions, namely optimizing airflow in existing kilns to improve combustion efficiency such as zig-zag firing, switching to cleaner brick-making technologies, and utilizing clay and coal with lower sulfur, carbon and metal contents.^21,44^ Additional recommendations on air quality management measures for clamp kilns based on findings from various firing campaigns in a model kiln included ensuring an even distribution of clay and fuel across the firing batch, ensuring a steady rise and fall in temperature, and ensuring adequate sun drying of bricks prior to firing to help reduce energy consumption.^21^

We did not find any studies on interventions to reduce exposures among BKWs, and only 5 studies that measured personal exposures and provided some recommendations on dust control and personal protection.^19,53,94,106,111^ These recommendations included the use of water to reduce dust when possible, a decrease in the long hours on the job leading to greater risk of overexposure, the use of respiratory protection with priority given to brick haulers and stackers, changing into clean clothes after work, and regular health surveillance. The authors also advocated for enforcement of stricter regulations on brick kilns.

## DISCUSSION

We conducted a comprehensive systematic review and meta-analysis of studies linking brick kiln pollution to environmental exposures and health. While published data presents on emissions related to brick kilns extensively, our review identified important knowledge gaps in personal exposures and evaluation of health outcomes among brick kiln workers. First, despite silica being one of the most important exposures to workers, we identified only four studies that measured personal exposures to silica and one that measured silica concentrations in kilns, two that provided data on silicosis and one that evaluated for profusions on chest X-rays out of 104 studies. Second, while most studies correctly focused on respiratory conditions, they largely ignored other important organ systems well known to be affected by pollution such as the cardiovascular and nervous systems. Indeed, hypertension and cardiovascular disease are the leading risk factor and cause of illness and death, respectively, worldwide,^117,118^ and long-term exposure to silica is linked to higher risk of cardiovascular disease mortality.^119^ Third, even among the studies that evaluated respiratory health, standardized questionnaires were not used consistently to report on respiratory symptoms, and guidelines and reference equations for spirometry were not reported in most studies. Most studies did not provide clear definitions of how respiratory symptoms were asked. Dyspnea, while generally higher in BKWs than in controls, was not consistently reported across studies. No studies reported Z-scores for lung function and only two studies out of 14 described the reference equations used to calculate percent predicted values. Lung function depends on age, sex, and height. Moreover, other risk factors including tobacco smoking, exposure to biomass smoke, and exposures to sources of air pollution other than brick kiln pollution also affect lung function. Without adjustment for these parameters, it is difficult to interpret whether differences in lung function between study groups are meaningful. Fourth, most studies were predominantly conducted in South Asia. While recognized as an important problem in South Asia, brick kilns are ubiquitous in low-and middle-income countries worldwide. Our review also identified potential research opportunities to improve our evidence base.

To our surprise, we found that only five studies (5%) measured silica concentrations or personal exposures,^8,19,53,106,111^ and only four reported personal exposures to other pollutants like PM_2.5_, RSP and CO.^48,94,106,111^ While the three studies reporting on silica exposures in low-and middle-income countries were limited in sample size and scope with no more than 48 participants,^19,53,111^ they nonetheless highlight the problem of silica exposure among brick kiln workers in countries where regulations may be less closely monitored. Silica exposures were severalfold higher than the American Conference of Governmental Industrial Hygienists threshold limit value (ACGIH TLV; 25 μg/m^3^) and the NIOSH recommended exposure limit (NIOSH REL; 50 μg/m^3^)^120^ and were similar or higher than other occupations where silica dust exposure is also prevalent like mining and pottery.^121,122^ Exposure to silica dust is associated with lung cancer, silicosis, kidney disease, chronic obstructive pulmonary disease and cardiovascular disease.^123^ Our review identified that better epidemiological data linking silica exposures in brick kilns with health outcomes is needed. Indeed, better studies estimating cumulative exposure to health outcomes will clearly help to drive policy for exposure mitigation in this neglected group of workers.

The most common study design was cross-sectional. Indeed, we did not identify a single study that collected longitudinal data to evaluate associations between long-term exposures and health outcomes or conducted a randomized controlled trial (RCT) to evaluate exposure mitigation strategies on health outcomes. Cross-sectional studies, while less expensive and faster to conduct than longitudinal studies, do not allow for causal or temporal relationships and may be prone to biases such as recall bias and the healthy worker effect.^124^ Common conditions including hypertension, chronic obstructive pulmonary disease, and chronic kidney disease would benefit from longitudinal studies to understand the relationship between cumulative exposures to silica and other brick kiln pollutants on health outcomes. Given that occupational exposure to brick kiln pollution is a preventable condition, individual-level interventions like the use of personal protective equipment or clustered interventions such as the use of water spray at kilns as strategies to mitigate personal exposures to silica and other pollutants may have measurable impacts on health outcomes. Moreover, RCTs can be used to study multiple primary outcomes including the impact of mitigation of brick kiln pollution exposures on musculoskeletal health where evidence is lacking. However, one difficulty with longitudinal studies or RCTs is that brick kiln workers tend to be migrants and temporary workers, which may complicate long-term follow-up due to high rate of loss to follow-up.

Other important observations identified in our review include the association between a higher number of seasons worked and health outcomes particularly for respiratory health; however, findings were not consistent across studies and were almost equally divided between higher prevalence and no effect. We also identified interaction effects between exposure to brick kiln pollution and tobacco smoking with worse lung function when compared to neither exposure or one exposure. The interaction between brick kiln pollution and tobacco smoking is important because of the well-recognized higher risk of mortality in individuals who are occupationally exposed to silica and who are also smokers.^125^ Our review also revealed a large variability in dust and silica exposures across job types within kilns.^19,94^ However, few studies examined the association between job type and health outcomes, and none showed a clear link between worse respiratory health and job type. More comprehensive, standardized research is needed to better understand the effect of job type on exposures and health outcomes and help inform exposure mitigation strategies.

Our review has several strengths. First, we performed a systematic search of all brick kiln studies ever conducted to provide one of the more comprehensive analyses of brick kiln pollution exposures and health. Second, our approach to our systematic review was registered a priori and we followed an explicit methodology to search for studies in existing databases. Third, our review has allowed us to identify potential opportunities for research and interventions to improve health in brick kiln workers. There are some potential shortcomings, however. Many studies were of low quality, some did not use comparison groups or reference equations that would allow for data comparisons, and standard questionnaires about respiratory symptoms were not used. Moreover, many studies had small sample size or did not specify whether sampling was done at random or by convenience, which may affect inferences. Multiple studies reporting pollution data did not specify the number of measurements collected or the sampling duration.

In conclusion, our review identified worse health outcomes, particularly respiratory health, in brick kiln workers when compared to controls but study quality supporting the evidence was low and methods were not reported consistently or accurately. Few studies have quantified personal exposure to silica, but the few that have suggest that exposures are high and may be similar or higher than in other occupations where silica dust is also a prevalent exposure. We identified knowledge gaps that can serve as research opportunities to better understand the relationship between brick kiln pollution and health among workers, and to evaluate the effectiveness of exposure mitigation strategies.

## Data Availability

All data produced in the present work are contained in the manuscript

