## Supplemental Material for "Brick kiln pollution and its impact on health: A systematic review and meta-analysis"

### List of abbreviations

Pollutants

| | |
|---|---|
| Ace | Acenaphthene |
| Acy | Acenaphthylene |
| Ant | Anthracene |
| B[b]A | Benzo[b]anthracene |
| B[b]F | Benzo[b]fluoranthene |
| B[b+k]F | Benzo[b+k]fluoranthene |
| B[k]F | Benzo[k]fluoranthene |
| B[ghi]P | Benzo[g,h,i]perylene |
| B[a]P | Benzo[a]pyrene |
| B[e]P | Benzo[e]pyrene |
| BC | Black carbon |
| BrC | Brown carbon |
| Chr | Chrysene |
| CO | Carbon monoxide |
| $CO_2$ | Carbon dioxide |
| Cor | Coronene |
| D[ah]A | Dibenzo[a,h]anthracene |
| EC | Elemental carbon |
| Fl | Fluorene |
| Fth | Fluoranthene |
| HCl | Hydrogen chloride |
| He | Helium |
| HF | Hydrogen fluoride |
| HCN | Hydrogen cyanide |
| Ind | Indeno[1,2,3-cd]pyrene |
| Nap | Naphthalene |
| $NH_3$ | Ammonia |
| NMVOC | Non-methane volatile organic compound |
| NO | Nitrogen oxide |
| $NO_2$ | Nitrogen dioxide |
| $NO_x$ | Nitrogen oxides |
| OC | Organic carbon |
| PAH | Polycyclic aromatic hydrocarbon |
| Phe | Phenanthrene |
| Py | Pyrene |
| PM | Particulate matter |
| $SO_2$ | Sulfur dioxide |
| $SO_3$ | Sulfur trioxide |
| TSP | Total suspended particulate |
| VOC | Volatile organic compound |
| WSOC | Water-soluble organic carbon |

Kilns

| | |
|---|---|
| BTK | Bull's trench kiln |
| CK | Clamp kiln |
| DDK | Downdraft kiln |
| FCBTK | Fixed-chimney Bull's trench kiln |
| FDZ | Forced-draft/induced-draft zigzag kiln |
| HK | Hoffmann kiln |
| MK | Marquez brick kiln |
| MK2 | Double-dome version of the original Marquez kiln |
| NDZ | Natural draft zigzag kiln |
| NS | Not specified |
| TCK | Traditional-campaign kiln |
| TFK | Traditional-fixed kiln |
| TIK | Traditional-improved kiln |
| TK | Tunnel kiln |
| VSBK | Vertical Shaft Brick Kiln |
| ZZK | Zigzag kiln |

### RESULTS

#### Literature search

We identified 1089 references through electronic searching. After removing duplicates, 1015 studies were available for screening. Of these, we excluded 807 based on title and abstract. Full texts for three studies could not be retrieved. A total of 205 full papers were assessed for eligibility, of which 101 met inclusion criteria and were included for review. An additional 6 studies were identified through manual searches, of which 3 were included in the review for a total of 104 studies (Figure 1). Most studies were conducted in South Asia (n = 74; 71%); primarily Pakistan (n = 30),[5,7,9,15,23,38–40,42,43,51,57,59,69,74,81–84,92,95–98,101,102,104,105,107,108] India (n = 20),[18,22,35,47,49,54,63,71–73,76,78–80,85,89,91,93,110,114] Nepal (n = 14),[6,8,19,34,41,44,55,56,60,94,103,109] and Bangladesh (n = 10);[20,24–26,28,29,29,36,70,87] 10 studies were conducted in Mexico,[30,31,33,46,48,50,65,86,99,100] 7 in China,[16,37,62,64,66–68] 4 in Vietnam,[18,32,45,63] 2 in Egypt,[88,111] 2 in Colombia,[61,77] 2 in Iran,[52,53] and 1 each in Indonesia,[90] Thailand,[58] South Africa,[21] Turkey,[75] England and Scotland.[106]

#### Brick kiln pollution

Thirty-two studies (41%) reported pollutant concentrations at the kilns, 20 (25%) reported emission factors, 16 (21%) reported source contributions, 14 (18%) reported exposure biomarkers, 6 (8%) reported personal exposures, 3 (4%) reported particle size distributions, and 2 (3%) reported radionuclide activity (Table S3). Pollutants measured included PM (n=47, 60%), $SO_2$ (n=24, 31%), CO (n=21, 27%), BC (n=14, 18%) $CO_2$ (n=13, 17%),$NO_x$ (n=12, 15%), PAHs (n=9, 12%), VOCs (n=8, 10%), $NO_2$ (n=8, 10%), EC (n=7, 9%), OC (n=6, 8%), $SiO_2$ (n=5, 6%), NO (n=5, 6%), $NH_3$ (n=4, 5%), HCl (n=4, 5%), HF (n=3, 4%), BrC (n=2, 3%),$SO_3$ (n=1, 1%), and

HCN (n=1, 1%). Some studies also reported data on the elemental composition (n=10, 13%)[24–29,54,60,62,67] and the content of metals (n=8, 11%),[30,41,47,49,52,61,68,96] and water-soluble inorganic ions (n=2, 3%)[41,65] of PM from kilns. Exposure biomarkers measured included metals/metalloids in urine and blood (n=8, 11%),[9,33,57,97,98,100–102] urinary hydroxylated AH metabolites (OH-PAHs) including 1-hydroxypyrene, α-naphthol and β-naphthol as biomarkers for PAH exposure (n=5, 7%),[33,46,50,100,105] urinary fluoride (n=2, 3%),[33,43] and trans,trans-muconic acid as a biomarker for benzene exposure (n=1, 1%).[33]

Most studies performed measurements at kiln sites (n=59, 76%). These included measurements of the flue gas (n=15, 20%), in-stack sampling (n=17, 23%), samples collected at various locations within the kiln site (16, 21%), and personal exposure assessments (6, 8%) and measurements of exposure biomarkers in BKWs (9, 12%) and children living at kiln sites (1, 1%). Of the studies reporting measurements outside kiln sites (n=14, 19%), 10 conducted source apportionment and 1 regressed pollutant concentrations against brick production to determine the contribution of brick kilns to air pollution in the surrounding areas, 1 compared ambient pollution concentrations during the brick kiln season and the pre-operational season, 1 compared PM levels in affected areas within a 3km radius of kilns and areas beyond the 3km radius, and 1 compared levels of air pollutants in an area near brick kilns and sugar mills and an area far from kilns and industry, during both the non-crushing period, when only brick kilns were operational, and the crushing period, when both sugar mills and brick kilns were operational. An additional 3 studies conducted outside of kiln sites reported exposure biomarkers among children living in communities with kilns.

***Pollutant emission factors from brick kilns***

The most reported emission factors were PM (n=18) and $SO_2$ (n=14), followed by CO (n=13), $CO_2$ (n=11), BC (n=7), VOCs (n=5), EC (n=4) OC (n=4), $NO_2$ (n=4), NO (n=4), $NO_x$ (n=4) and PAHs (n=3) (Table S4).

Five studies reported various VOC emission factors.[31,36,37,56,65] Christian et al.[31] sampled 3 traditional-fixed kilns in Central Mexico mostly burning biomass fuels and found similar NMOCs as those emitted from biomass burning. In a different study in Mexico, fuel-based emission factors for methane, methanol and acetic acid from a traditional-fixed kiln were 3-5, 2-8 and 5 times higher, respectively, than those reported by Christian et al.[31], consistent with a lower modified combustion efficiency.[65] Compared to an MK2 and a traditional-campaign kiln, emission factors from the traditional-fixed kiln were higher for methane, ethane, methanol and acetaldehyde, but lower for benzene and toluene. Hu et al.[37] analyzed total VOCs from pharmaceutical, brick and food industries in China and found much lower mean VOC emission factors from brick manufacturing factories (0.52 ± 0.92 kg/t fuel) compared to food (4.83 ± 10.9 kg/t fuel) and pharmaceutical (5.03 ± 8.35 kg/t fuel) companies. A study across three different kiln types in Bangladesh found VOC emission factors were lower for the fixed chimney kilns (1085 ± 947 g/kg fuel) compared to improved designs such as the zigzag (1371 ± 526 g/kg fuel) and Hoffman kiln (1953 ± 972 g/kg fuel) as VOC emissions depend not only on kiln design but also on quality of fuel used.[36] Lastly, Stockwell et al.[56] measured emissions from a clamp kiln and a zigzag kiln in Nepal and found drastic differences in the NMOC emission factors for the two kiln types with clamp-to-zigzag-kiln EF ratios of 223, 2604, 30, 203, 16, and 28 for methane, ethane, ethylene, benzene, methanol, and phenol, respectively. In addition, other species such as formaldehyde, furan, hydroxyacetone, and ammonia were also emitted at high levels from the clamp kiln but were below the detection limit in the zigzag kiln. Methane emissions from the clamp kiln were among the highest measured from any combustion source, with an emission factor of 19.5 g/kg fuel.

Three studies reported PAH emission factors.[16,34,41] Chen et al.[16] measured various types of PAHs in both stack and fugitive emissions from two typical types of annular brick kilns in China. The emission factors of PAHs and all other incomplete combustion products, including TSP, $PM_{10}$, $PM_{2.5}$, EC, and OC from the stack gas were significantly lower than those from the fugitive source for both kiln types. The authors attributed this reduction in the quantity of incomplete combustion products to the longer distance traveled at high temperatures (> 1000 □C) and hence longer reaction time along the chimney; however this is inconsistent with the higher ratio of CO to $CO_2$ observed in the stack (0.14 for K1 kilns; 0.16 for K2 kilns) compared to the fugitive emissions (0.05 for K1 kilns; 0.05 for K2 kilns).  Goetz et al.[34] sampled $PM_1$ emissions from a batch-style clamp kiln and a zigzag kiln using a mini aerosol mass spectrometer. PAHs were not observed above detection limits from either of the kilns.  On the other hand, Jayarathne et al.[41] did detect PAHs from the clamp kiln by filter-based measurements, with the most abundant PAHs being chrysene, benz(a)anthracene, benzo(e)pyrene, and 1-methylcrysene. Picene, a molecular marker for coal combustion, was also detected in all clamp kiln samples.

***Pollutant concentrations at brick kiln sites***

We identified 22 studies reporting PM concentrations, 14 reporting $SO_2$, 11 reporting CO, 9 reporting $NO_x$, 7 reporting $CO_2$, 5 reporting $NO_2$, 3 reporting BC, 3 reporting VOCs, 2 reporting NO, and 1 reporting OC and EC (Table S4).

We also identified 6 studies reporting various PAH concentrations.[30,39,40,54,62,104] Bruce et al.[30] measured PAH concentrations from an active and a filter MK burning motor oil, and an active MK burning wood in Northern Mexico. They found that most PAH concentrations were higher for samples taken during wood burning, except for benzo[b]fluoranthene, and benzo[k]fluoranthene which were higher when oil was used. Similar concentrations of pyrene and phenanthrene were observed with both fuels. PAH concentrations emitted from the filter kiln were significantly lower than those from the active kiln. PAHs extracted from soot samples from five kilns in Pakistan revealed a wide variability across kilns.[39] Mean concentration was highest for acenaphthylene (845 ± 1808 mg/kg), followed by phenanthrene (74.9 ± 163.0 mg/kg), fluoranthene (33.2 ± 45.5 mg/kg), naphthalene (26.8 ± 33.6 mg/kg), anthracene (22.3 ± 37.8 mg/kg), chrysene (20.0 ± 30.6 mg/kg), and fluorene (15.0 ± 18.6 mg/kg). Benzo[a]pyrene was not found in any of the samples. In a different study, PAHs were measured in surface soil dust samples from three kilns in Punjab province, Pakistan.[104] PAH concentrations in these samples were significantly lower than those found in soot, with the highest mean concentration being 0.301 ± 0.237 mg/kg for phenanthrene, and a mean total PAH concentration of 1.528 ± 1.416 mg/kg (Table S6). On the other hand, soil samples from a brick kiln in Peshawar, Pakistan, showed phenanthrene levels below the detection limit and naphthalene at the highest concentration (0.2167 mg/kg).[40] Finally, a study conducted in North East India, measured PAH concentrations in PM samples collected around a brick kiln.[54] Concentrations of all PAHs measured were below 0.133 ng/m$^3$ in $PM_{10}$ samples. In $PM_{2.5}$, fluorene showed the highest concentration (56.3 ng/m$^3$) followed by naphthalene (18.7 ng/m$^3$), acenaphthene (18.2 ng/m$^3$), and acenaphthylene (17 ng/m$^3$). All other PAH concentrations were below 0.5 ng/m$^3$. In a different study across 4 brick production enterprises in the Guanzhong Plain, China, the mean average total concentration of PAHs in $PM_{2.5}$ was 24.4 ng/m$^3$, with benzo[k]fluoranthene contributing the largest amount (2.76 ± 1.40 ng/m$^3$).[62]

Six studies reported metal concentrations.[30,47,49,52,61,96] Measurements by Bruce et al.[30] in the emissions of the active and filter MK burning motor oil revealed lead concentrations of 2.41 and 1.43 µg/m$^3$ and barium concentrations of 28.85 and 14.36 µg/m$^3$, respectively. No cadmium was detected in the emissions of either of the kilns. Ubaque et al.[61] measured concentrations of 10 heavy metals (Sb, As, Cd, Co, Sn, Cr, Cu, Mn, Ni) and mercury in kiln emissions during co-firing of municipal solid waste in Colombia. The mean heavy metal concentration across six measuring points along the HK vault was 0.0947 ± 0.01554 mg/m$^3$ (Table S6), with 77.8 – 86.1% of measurements below the threshold value of 0.5 mg/m$^3$ issued by the Ministry of Environment, Housing and Territorial Development (MAVDT). Mean mercury concentrations were all < 0.0002 mg/m$^3$, with > 99.8% of samples below the MAVDT threshold value of 0.2 mg/m$^3$. Pangtey et al.[49] measured metals in the soil used for making bricks across various kilns in Lucknow, India. The highest mean concentrations were for Fe (4494.6 µg/g), followed by Mn (230.03 µg/g), Pb (30.04 µg/g), Zn (28.66 µg/g), Cu (8.82 µg/g), Cr (6.19 µg/g) and Cd (1.05 µg/g) (Table S6). Another study in Isfahan Province, central Iran, where soil samples were collected at 0-500 m from a traditional kiln, detected lower Pb concentrations than those reported by Pangtey et al.[49], but otherwise similar concentrations for Zn, Cu and Cd.[52] On the other hand, most heavy metal concentrations in soil samples collected from 2 FCBTKs and 2 ZZKs in Pakistan were found to be lower than those reported by Pangtey et al.[49] or Ravankhah et al.[52], with Zn having the highest mean concentration (9.3 µg/g) followed by Cr (7.52 µg/g), Ni (5.09 µg/g), Mn (0.68 µg/g), Cd (0.34 µg/g) and Cu (0.14 µg/g).[96] Lastly, analysis of brick kiln bottom ash samples in West Bengal and Assam, India, revealed metal concentrations that were an order of magnitude higher than those measured in soil, with the exception of Mn which was similar.[47] As in the soil samples in Pangtey et al.[49], concentrations were highest for Mn, followed by Pb, Zn, Cu, Cr and Cd.

***Contribution of brick kiln emissions to surrounding ambient air pollution***

A total of 16 studies reported the contribution of brick kiln emissions to the surrounding ambient air pollution in various locations in South Asia, including Dhaka, Bangladesh (n=5),[24,26–29] Kathmandu Valley, Nepal (n=4),[44,55,112,113] Bhaktapur, Nepal (1),[103] Chittagong, Bangladesh (n=1),[25] Patna, India (n=1),[22] Ghaziabad, India (n=1),[35] Tehsil Darya Khan, Pakistan (n=1),[95] Peshawar district, Pakistan (n=1),[7] and Rawalpindi, Lahore and Gujranwala, Pakistan (n=1).[42]

Using two different methods for source apportionment (positive matrix factorization [PMF2] and multilinear-engine [ME-2] modeling), Begum et al.[24] identified biomass-burning and brick kilns as a source of fine, but not coarse PM, at an urban area in Dhaka, with estimated contributions to $PM_{2.2}$ in 2001-2002 of 29.1% and 37.5%, by ME-2 and PMF2, respectively. In a later study conducted at the same site, the authors[29] conducted PMF2 modeling using both total OC and EC concentrations (model 1) and the fractions of OC and EC (model 2). The estimated contributions of brick kilns to fine PM between 2010 and 2011, were 30% and 41%, using models 1 and 2, respectively, like those observed in 2001-2002. Similar measurements and modeling conducted at a semi-residential area in Dhaka between 2005 and 2006 identified biomass-burning/brick kilns as a source of both fine and coarse PM, contributing to 29.9% and 25.3% of fine and coarse PM, respectively, during winter.[26] Overall, brick kilns were the largest contributor to fine PM, responsible for 38.1% of the yearly concentration. Brick kilns contributed an estimated 22% of fine PM and 91% of BC at this site between 2007 and 2009,[27] and 21% of $PM_{2.5}$, 24% of BC, and 84% of sulfur between 1997 and 2005.[28] At a site in Chittagong, Bangladesh, located away from major sources of pollution, biomass-burning/brick kilns contributed an estimated 35.5% and 22.7% to fine and coarse PM, respectively, between February and July 2007.[25] In Ghaziabad, an industrial city adjoining New Delhi, India, the annual contribution of brick kilns to ambient air pollution between June 2018 and May 2019 was less than 10% for both $PM_{2.5}$ and $PM_{10}$, but up to 29% of $PM_{2.5}$ during the pre-monsoon season.[35] $PM_{10}$ and NMVOC measurements were conducted by two separate groups at Bode, a semi-urban site in the Kathmandu valley, during the winter of 2012-2013.[44,55] A large variation in the $PM_{10}$ concentrations was observed between the sampling periods before and after all brick kilns in the area were fully operational.[44] Brick kilns were the largest primary source contributor of $PM_{10}$ during the second sampling period (28%), and the largest primary source of EC, contributing 40% of the average EC concentration of both periods. Biomass co-fired brick kilns contributed 10.4% of total NMVOC concentrations, and were the largest contributor of benzene, at ~37%, and second largest contributor of all other measured NMVOCs, including acetonitrile, toluene, styrene, xylenes and trimethylbenzenes.[55]

Another study in Patna, India, measured BC, $PM_{2.5}$ and $PM_{10}$ concentrations at seven monitoring sites, including two brick kiln clusters.[22] Pollutant concentrations at the clusters were significantly higher during the winter and pre-monsoon seasons, when kilns were operational. Mean concentrations of BC, $PM_{2.5}$ and $PM_{10}$ at these sites were $13.27 \pm 3.34$ μg/m$^3$, $65.65 \pm 18.08$ μg/m$^3$ and $102.86 \pm 20.63$ μg/m$^3$ in the winter vs $4.74 \pm 1.54$ μg/m$^3$, $36.02 \pm 10.72$ μg/m$^3$ and $59.44 \pm 14.14$ μg/m$^3$ in the monsoon season. The estimated contribution of brick kiln emissions to BC, $PM_{2.5}$ and $PM_{10}$ in Patna during 2015 was $6.0 \pm 6.4$%, $19.8 \pm 2.5$% and $15.0 \pm 1.3$%, respectively.

Khan et al.[95] collected $PM_{10}$, $SO_2$ and $NO_x$ from areas in Bhakkar-Punjab, Pakistan, with brick kilns and sugar mills (affected), as well as controlled areas away from operational kilns and industry. Data were collected during both the non-crushing period, when only brick kilns were operational, and the crushing period, when both sugar mills and brick kilns were operational. During the non-crushing period, mean concentrations of $PM_{10}$, $SO_2$ and $NO_2$ in the affected areas were 115.3 μg/m$^3$, 2.3 μg/m$^3$ and 5.0 μg/m$^3$, respectively, compared to 17.9 μg/m$^3$, 0.5 μg/m$^3$ and 1.8 μg/m$^3$ in the controlled areas.

Data collected in an area with brick kilns in the Bhaktapur district, Nepal, revealed higher mean $PM_{10}$ and TSP concentrations during the brick kiln season compared to the monsoon season when brick kilns are closed (50 μg/m$^3$ vs 29 μg/m$^3$ for $PM_{10}$; 56 μg/m$^3$ vs 33 μg/m$^3$ for TSP).[103] Interestingly, no $SO_2$, CO or $NO_x$ was detected during either season.
Another study conducted in the Peshawar district in Pakistan, measured PM concentrations at sites within a 3km radius of brick kilns (treated) and control sites beyond the 3 km radius.[7] Both $PM_{10}$ and $PM_{2.5}$ were substantially higher within 3km of brick kilns, with mean ± SD concentrations of 524 ± 258 μg/m$^3$ and 158 ± 34 μg/m$^3$, respectively, compared to 113 ± 18 μg/m$^3$ and 34 ± 3 μg/m$^3$ at control sites.

Finally, Kamal et al.[42] evaluated PAH concentrations in three districts of the Punjab Province, Pakistan, and regressed them against number of bricks produced and total wood consumption per year. The authors presented the coefficients of determination for the two linear models and showed that 81.4% and 53.7% of the variance in ambient PAH concentrations could be explained by wood consumption and brick production, respectively. However, these models did not account for confounding. Moreover, the coefficient of variation provides a measure of goodness of fit but does not explain the contribution of independent variables to a dependent variable.

***Exposure biomarkers***
Three studies conducted in Mexico found that urinary concentrations of 1-hydroxypyrene (1-OHP), a biomarker for PAH exposure, were higher in children living near brick kilns than those living near heavy traffic, waste landfill or metallurgical industry (Table S11).[33,46,50] Urinary concentrations of trans,trans-muconic acid (t,t-MA), a biomarker for benzene exposure, were also highest among children living near brick kilns.[33] Another study among adult male BKWs and non-occupationally exposed controls in Pakistan reported median urinary concentrations of 1-OHP $\alpha$-naphthol and β-naphthol that were 2.5, 5.7 and 2.3 times higher, respectively, in BKWs than controls.[105] A different study conducted in Pakistan among BKWs and individuals working and living in areas with no exposure to fluoride found that urinary fluoride concentrations were significantly higher in BKWs compared to controls (Table S11).[43] Interestingly, fluoride concentrations reported by Flores-Ramirez et al.[33] for children living near kilns were an order of magnitude larger.

Measurements of metal and metalloid concentrations in urine among schoolchildren from a high-pollution and low-pollution area, children working in carpet weaving and in brick kilns in Lahore, Pakistan, revealed that BKWs had the highest values for many elements, with the most marked difference for arsenic and uranium.[57] The study by Flores-Ramirez et al.[33] on the other hand, found arsenic concentrations were similar across all four communities examined and below the reference value of 15 μg/L. In a different study conducted in Punjab, Pakistan, heavy metal (Pb, Cd, Cr) concentrations in blood were significantly higher in all BKW groups (brick makers, brick carriers, brick bakers) compared to controls (Table S11).[102] Nonworkers living near kiln sites also had significantly higher Cd concentrations compared to controls. Similarly, children living at kiln sites in the same area, were found to have higher heavy metal (Zn, Cd, Cr, Ni) concentrations in blood compared to controls living far from kiln sites (Table S11).[101] Male workers at those brick kilns had significantly higher levels of Cd, Cr and Ni, but not Zn, compared to control participants.[9] In a study[97] among workers in construction industries (n=250) and sex- and age-matched non-exposed controls (n=250), mean concentrations of arsenic in blood were highest in BKWs compared to furniture workers, painters, welders and controls (Table S11). The same values were reported in a follow-up study on a different sample of industrial workers (n=300) and controls (n=300).[98] Interestingly, blood lead concentrations measured among BKWs in Tercera Chica,[100] Mexico were lower than those among children living in the area, [33] and all below the established standard of 5 µg/dL.[33,100]

### Health outcomes

#### *Respiratory health*

We identified 34 studies that evaluated respiratory conditions in either BKWs alone or in BKWs and a reference group, or in groups of participants living near or far away from kilns (Table S13).[5,9,70–73,76–80,82,84,86–89,91–96,99,100,103,106–111,114,126] All studies were cross-sectional. Seven studies (21%) were conducted in both adults and children, 23 (68%) were conducted in adults only, 4 (12%) were conducted in children only, 11 (32%) were conducted in men only, 1 was conducted in women only, 17 (50%) were conducted on BKWs only, 12 (35%) compared respiratory outcomes between BKWs and some type of reference group, 3 (9%) were conducted in participants living near or far away from the kilns, 10 (29%) evaluated the association between respiratory health outcomes and number of years worked at the kilns, 10 (29%) examined health outcomes by type of BKW occupation, and 27 (79%) were reportedly conducted in multiple brick kilns (Table S13). Of the 12 studies that compared BKW to a reference group, 4 (31%) were matched on age, sex and/or other socioeconomic variables.[79,82,88,114]

A total of 14 studies (41%) reported data on lung function, 14 (41%) reported information on cough or chronic cough, 9 (26%) reported information on phlegm or chronic phlegm, 11 (32%) reported information on breathlessness, dyspnea or shortness of breath, 8 (24%) reported information on wheezing, 7 (21%) reported information on chronic bronchitis, 8 (24%) reported information on asthma, 4 (12%) reported information on COPD (Table S13). Additional studies reported respiratory conditions such as acute (or upper) respiratory infections, non-specific respiratory symptoms, tonsillitis, pharyngitis, self-reported history of tuberculosis, and silicosis.[91,111] One study reported the prevalence of chest symptoms as part of the Revised National Tuberculosis Control Programme in India.[93] Ten studies (29%) referenced the source of the questionnaire used to ask respiratory symptoms or provided operational definitions and illnesses.[5,70,77,78,88,94,99,100,106,114]

Of the 14 studies that reported data on lung function, 11 (79%) reported lung function values[71–73,76,79,84,86,88,100,108,114] and 7 (50%) reported percent predicted values for lung function values;[82,84,86,88,91,107,114] however, only two of the seven studies specified what reference population was used for the calculation of percent predicted values. Only 1 study conducted post-bronchodilator spirometry,[86,91,100] and none reported Z-scores. Only 5 (36%) studies explicitly reported following international guidelines for the conduct of spirometry.[82,84,91,100,114] Overall quality grading for all studies was low or not reported . One study,[111] not included in our count of studies that reported data on lung function, described collecting pulmonary function but did not report data in their paper or in a subsequent paper.

#### *Biomarkers*

A total of 11 studies evaluated biomarkers in BKWs against unexposed controls. All studies were cross-sectional.[9,74,79,81,83,97–99,101,102,105]

Ahmad et al.[97] evaluated 250 arsenic-exposed workers (65 of whom were BKWs with mean age 30.4 ± .2.1 years, 21% smokers) and 250 controls (mean age 27.0 ± 1.6 years, 55% smokers). BKWs also had down-regulated expression of Proteasome 26S Subunit, Non-ATPase 1 (PSMD1) when compared to unexposed controls. Workers with >10 years of exposure had a greater down-regulation of PSMD1 when compared to those with <10 years of exposure; however, the authors did not stratify the analysis by type of worker.

Akram et al.[98] evaluated blood samples of 300 occupational workers with ≥ 5 years of exposure (60 per industry, one category was brick kilns) and 300 age- and sex-matched unexposed controls for Variation in HPRT, OGG1 gene expression, and DNA damage in blood cells by comet assay. Mean age was 27,9 ± 21.1 years and 53% were smokers. Brick kiln workers had a significantly longer comet length (mean ± SE were 122 ± 6.0 vs 99.6 ± 2.3, units not reported), longer head length (98.3 ± 4.9 vs. 78.6 ± 2.04) and longer tail length (23.9 ± 2.2 vs. 16.92 ± 0.8) when compared to controls. While OGG1 and HPRT gene expression was significantly downregulated in occupational workers when compared to controls, the authors did not present their analysis by work group. Workers with >10 years of exposure had a greater down-regulation of OGG1 and HPRT when compared to those with <10 years of exposure; however, the authors did not stratify the analysis by type of worker.

Berumen-Rodriguez et al.[99] evaluated cytokine levels in exhaled breath condensate of 21 BKWs (no controls). They found high cytokine concentrations when compared to other occupational exposures.

David et al.[74] evaluated differences in the biochemical profile and cortisol in female BKWs (mean age 35.8 ± 1.9 years, 27.1% smokers) and unexposed controls (mean age 27.7 ± 1.4 years, 0% smokers). Female BKWs had lower levels of superoxide dismutase (SOD) and guaiacol peroxidase (POD) activity, higher levels of reactive oxidative species, low-density lipoprotein, triglycerides, and cortisol when compared to controls. Female BKWs had lower hemoglobin, red blood cells, mean corpuscular hemoglobin (MCH) and higher platelets when compared to controls, albeit non-statistically significant. David et al.[101] also evaluated children living and working at kilns and unexposed children who lived farther away from kilns to compare differences in reactive oxidative species, growth hormone and cortisol, hematological parameters and DNA damage. Children living and working at kilns had lower levels of SOD,

POD, catalase activity, growth hormone, MCH, and a higher average tail length and percent of DNA in the tail of single cell gel electrophoresis assay, and higher levels of hemoglobin, cortisol when compared to unexposed children. David et al.[9] subsequently evaluated 546 participants, 342 adult, male BKWs and 200 male non-BKW controls (mean age 24.4 ± 7.7 years) who lived at least 40 miles away from the nearest kiln, for serum antioxidant enzyme activity of SOD, POD, reactive oxidative species (ROS) and lipid peroxidation via malondialdehyde. Like other studies, SOD and POD were lower, and ROS was higher in BKWs when compared to controls. Jahan et al.[102] evaluated hematological, biochemical and hormone profiles in 110 BKWs (mean age 27.3 ± 12.1 years), 30 non-BKWs living in same vicinity as the kilns (27 ± 8.5 years) and 57 unexposed adult males (25.9 ± 5 years). Antioxidant enzymes (CAT, SOD, POD, GSH, and GR) and testosterone were reduced in BKWs as compared to unexposed adult males while thiobarbituric acid reactive substance was higher. Hematological profiles were, on the most part, similar between BKWs and the two groups of controls.

Kamal et al.[105] evaluated polycyclic aromatic hydrocarbon (PAH) exposure and other biomarkers in BKWs (mean age 39.6 ± 10.3 years) and a non-occupationally exposed group (mean age 42.2 ± 13 years). They found higher levels of C-reactive protein and lower levels of SOD, hemoglobin, red blood cells in BKWs when compared to controls. Brick bakers appeared to have lower CAT when compared to other BKW occupations.

Kaushik et al.[79] evaluated oxidative stress and DNA damage in 31 BKWs and 32 controls. They found that serum ferric reducing ability of plasma, glutathione S-transferase and reduced glutathione were all higher and serum malonedialdehyde was lower in BKWs when compared to controls. They also identified DNA fragmentation or smearing in 35.5% of BKWs and 12.5% of controls, however, this difference was not statistically significant.

Khisroon et al.[81] evaluated DNA damage in 100 male BKWs (mean age 28.4 ± 6.6 years, 26% smokers) and 50 nearby male residents without a history of brick kiln work (mean age 28.6 ± 5.4 years, 25% smokers). Using an alkaline comet assay, they found a significantly higher total comet score (indicating a longer tail when compared to the diameter of the comet nucleus) in BKWs when compared to controls. BKWs with >15 years of occupation also had a significantly higher total comet score when compared to those with 15 years or less.

Raza et al.[83] evaluated 200 workers exposed to arsenic (including 50 BKWs with mean age 30.4 ± 2.1 years) and 200 unexposed controls (mean age 27.0 ± 1.6 years) and found the BKWs had the highest level of DNA fragmentation when compared to other workers (data in controls not reported). They also reported significantly lower levels of CAT, SOD, glutathione peroxidase in BKWs when compared to controls; however, they did not find differences in CAT, SOD and glutathione peroxidase between BKWs with >10 years of work exposure and those with <10 years of exposure.

**Table S1. Characteristics of included studies with brick kiln pollution data.**

| Study | Country | Dates | Methods | Measurement locations | Sample size | Number of measurements | Sampling duration | Pollutants measured | Values reported |
|---|---|---|---|---|---|---|---|---|---|
| Abedin 2020[20] | Bangladesh | NA | Radionucleotides in coal and ash samples measured using a co-axial high-purity germanium (HPGe) gamma- ray detector (GC 2018, CANBERRA, USA). | Brick kilns | 3 coal-fired brick kilns | 30 samples (10 feed coal, 10 fly ash, and 10 bottom ash) | NA | 226Ra, 232Th, 40K | Radionuclide activity |
| Ahmad 2020[97] | Pakistan | NA | Levels of inorganic arsenic, lead, and cadmium measured in blood samples of control and exposed workers (brick kiln, wood, paint, welding) using flame atomic absorption spectrophotometer (Varian AA240FS) equipped with hydride generator. | Workers | 65 brick kiln workers, 250 controls | 1 sample/worker | NA | As, Pb, Cd | Exposure biomarkers |
| Akinshipe 2018[21] | South Africa | NA | Gaseous pollutants measured with electrochemical sensors. PM measured using the SidepakTM Personal Aerosol Monitor Model AM510 and the DustTrakTM DRX Handheld Aerosol Monitor Model 8534. | Flue gas | 1 model CK | 13 firing cycles | Hourly samples over 8-14 days | $SO_2$, $NO_2$, NO, $NO_x$, PM, CO, $CO_2$ | Pollutant concentrations; Emission factors |
| Akram 2022[98] | Pakistan | NA | Total arsenic, cadmium and lead content measured in blood samples of control and industrial workers using a flame atomic absorption spectrophotometer (Varian (AA240FS). | Workers and unexposed control subjects | 60 brick kiln workers; 300 controls | 1 sample/participant | NA | Metals (As, Cd, Pb) | Exposure biomarkers |
| Arif 2018[22] | India | Jan-Dec 2015 | Continuous BC measurements with a 7 channel micro-aethalometer (Model AE- 42, Magee Scientific, USA) and $PM_{2.5}$, $PM_{10}$ measurements with a portable scattering Nephelometer (pDR-1500, Thermo Scientific, USA). | Seven monitoring sites in the city, including 2 brick kiln clusters | 2 brick kiln clusters | NA | 5-minute intervals | BC, $PM_{2.5}$, $PM_{10}$ | Source contributions |

| | | | | | | | | | |
|---|---|---|---|---|---|---|---|---|---|
| Bashir 2023[23] | Pakistan | NA | Ratnam device used to measure real-time concentrations of SO2, CO, CO2 and PM2.5 and collect PM2.5 on filters for gravimetric analysis. | Stack | 1 FCBTK, 1 ZZK | NA | 4-6 hr sampling | CO, $CO_2$, $SO_2$, $PM_{2.5}$, BC | Pollutant concentrations; Emission factors |
| Beard 2022[8] | Nepal | May 2018 | Respirable dust samples collected using SKC AirLite (SKC Inc., Eighty Four, PA) sampling pumps calibrated to 2.5 L/min and an aluminum cyclone with a 4.0-µm cut-point. Filters were analyzed for silica via X-ray diffraction. CO and CO2 measured using Drager Pac 7000 Personal Gas Detectors (SKC). NO2 and SO2 cocnetrations determined using UMEX200 Passive Samplers (SKC). | Inside and outside homes of fire masters and workers, approx. 100m away from kiln | 16 indoor and 16 outdoor locations across 4 BTKs | 1 sample/location | Mean sampling time of 6.67 hrs | Silica, CO, $CO_2$, $NO_2$, $SO_2$ | Pollutant concentrations |
| Begum 2005[24] | Bangladesh | Jul 2001 - Mar 2002 | $PM_{2.2}$ & $PM_{10-2.2}$ samples with a "Gent" stacked filter. Concentration of BC in PM2.2 determined by reflectance measurement using an EEL Smoke Stain Reflectometer. Multielement analyses of the samples made with Proton Induced X-ray Emission (PIXE). Analysed by positive matrix factorization (PMF) and multilinear engine (ME-2) to estimate sources contributions. | Traffic hotspot site in Dhaka, 4.8 m above the ground | 1 site | 132 samples | 8-20 hr distributed uniformly over 24 hr a day | BC and elemental concentrations in $PM_{10-2.2}$ and $PM_{2.2}$ | Source contributions |

| | | | | | | | | | |
|---|---|---|---|---|---|---|---|---|---|
| Begum 2009[25] | Bangladesh | Feb-Jul 2007 | $PM_{10-2.5}$ and $PM_{2.5}$ gravimetric samples using a Thermo Andersen dichotomous sampler. BC measured using an EEL (Evans Electroselenium Limited) Smoke Stain Reflectometer. Multielemental analyses of the samples using proton induced X-ray emission (PIXE) and proton elastic scattering analysis (PESA). PMF modeling used to calculate average source contribution. | Chittagong Television Station Campus, 100 m above ground | 1 site | 104 samples | NA | BC, and elemental concentrations in $PM_{10-2.5}$ and $PM_{2.5}$ | Source contributions |
| Begum 2011[26] | Bangladesh | Feb 2005-Dec 2006 | $PM_{10-2.5}$ and $PM_{2.5}$ samples collected with size-fractionating aerosol sampler (Gent stacked filter unit). BC measured using an EEL Smoke Stain Reflectometer. Multielement analysis using PIXE. Source apportionment resolved with PMF. | Semi-residential area in Dhaka | 1 site | 161 samples | 24-hr sampling | $PM_{10-25}$, $PM_{2.5}$, and BC and elemental concentrations in $PM_{10-2.5}$ and $PM_{2.5}$ | Source contributions |
| Begum 2013a[27] | Bangladesh | 1996-2011 | $PM_{2.2}$ & $PM_{10-2.2}$ samples with a "Gent" stacked filter. Concentration of BC in $PM_{2.2}$ determined by reflectance measurement using an EEL Smoke Stain Reflectometer. Multielement analyses of the samples made with PIXE. Analyzed by positive matrix factorization (PMF) to estimate sources contributions. | Semi-residential area in Dhaka, 6.8 m above ground | 1 site | 100 samples/year | 6-20 h (depending on seasons) distributed uniformly over 24 h a day | $PM_{2.2}$, and EC and elemental concentrations in $PM_{2.2}$ | Source contributions |
| Begum 2013b[29] | Bangladesh | Feb 2010-Feb 2011 | $PM_{2.5}$ samples collected on Teflon and Quartz filters using Air Metrics MiniVol samplers. BC measured using an EEL Smoke Stain Reflectometer. Multielemental analyses made using ion beam analysis (IBA). Source apportionment by PMF modeling. | Traffic hotspot site in Dhaka, 10 m above the ground | 1 site | 126 samples | 24-hr sampling | $PM_{2.5}$, and OC, EC and elemental concentrations in $PM_{2.5}$ | Source contributions |

| | | | | | | | | | |
|---|---|---|---|---|---|---|---|---|---|
| Begum 2019[28] | Bangladesh | Dec 1996 - Sep 2015 | $PM_{2.5}$ samples collected with a "Gent" stacked filter. Concentration of BC determined by reflectance measurement using an EEL Smoke Stain Reflectometer. Multielement analyses of the samples made with Ion Beam Analysis. Analyzed by positive matrix factorization (PMF) to estimate sources contributions. | Semi-residential area in Dhaka, 6.8 m above ground | 1 site | NA | 24-hr sampling | $PM_{2.5}$, and BC and elements in $PM_{2.5}$ | Source contributions |
| Berumen-Rodriguez 2021[100] | Mexico | Nov 2020 | $PM_{10}$ collected with a high-volume sampler (Hi-Vol) equipped with quartz filter and $PM_{2.5}$ collected with a low volume sampler (Mini-Vol) with PVC filters. Blood lead concentration determined using an atomic absorption spectrophotometer. OH-PAH concentration in urine determined using Isotope Dilution Gas Chromatography/Tandem Mass Spectrometry. | 10 m from source; brick kiln workers | NA; 42 brick kiln workers | 6-7 samples; 1 sample/participant | 8-hr sampling; NA | $PM_{2.5}$, $PM_{10}$; Pb, OH-PAH | Pollutant concentrations; Exposure biomarkers |
| Berumen-Rodriguez 2023[99] | Mexico | NA | Exposure to PAHs and toluene evaluated using hydroxylated markers of PAHs and hippuric acid in urine, respectively. Urine samples analyzed using a gas chromatograph (Agilent 6890 GC) coupled to a mass spectrometry detector (Agilent 5975 MSD). | Brick kiln workers | 21 brick kiln workers | 1 sample/participant | NA | OH-PAHs | Exposure biomarkers |

| | | | | | | | | | |
|---|---|---|---|---|---|---|---|---|---|
| Bruce 2007[30] | Mexico | NA | Gravimetric & nephelometric sampling of PM. PAHs from 3 filters extracted using ultrasonic extraction (EPA method 3550B). Samples analysed for metals using inductively induced plasma (EPA method 6010B). Active carbon filters analyzed via GC/mass spectrometry for VOCs. Size distributions from 0.07 to 16 µm measured using optical sizing spectrometers (Particle Measuring Systems). | Flue gas | Several TFK, 2 MK, 2 MK2 | NA | NA | PM, PAHs, metals | Pollutant concentrations; Size distributions |
| Chen 2017[16] | China | Jul 2013 (Xiamen, Southern China) and Dec 2014 (Xi'an-Baoji-Weinan, Northern China) | $PM_{2.5}$ and $PM_{10}$ samples collected on glass fiber filters using impaction samplers (PEM, SKC, USA). TSP, EC, OC and particulate phase PAHs samples trapped on quartz fiber filters using regular samplers (Pall Co. USA). Polyurethane foam used to collect gaseous phase PAHs. | Flue gas and fugitive emissions | 8 K-1 (Xiamen) HK, 5 K-2 (Xi'an-Baoji-Weinan) HK | 2 samples/kiln | 40-min measurements | $CO_2$, CO, EC, OC, $SO_2$, $NO_2$, NO, TSP, $PM_{10}$, $PM_{2.5}$, PAHs | Emission factors |
| Christian 2010[31] | Mexico | Spring 2007 | Measured trace gas emissions with a mobile, rolling cart-based Fourier transform infrared spectrometer. Used an integrating nephelometer (Radiance Research M903) to determine $PM_{2.5}$. Measured $CO_2$ with the LICOR LI-7000 gas analyzer. | Flue gas | 3 TFK | NA | NA | $PM_{2.5}$, $CO_2$, CO, NH3, HCl, VOCs (CH4, CH3OH, C2H4, C2H2, C3H6, HAc, HFo, HCHO) | Emission factors |
| Co 2009[32] | Vietnam | 11-16 Apr 2006 | PM from stack gas collected on glass fiber filters using the STL-Combi dust sampler. CO and $SO_2$ measured using an automatic gas analyzer (Quintox, KM9006). | Stack | 1 TIK, 3 sites | NA | 2-3 hr/day for 4 days | CO, $SO_2$, PM | Emission factors |

| | | | | | | | | | |
|---|---|---|---|---|---|---|---|---|---|
| David 2020 | Pakistan | March 2018 - March 2019 | Metal concentrations in whole blood measured using an AA 40 FS Fast Sequential Atomic Absorption Spectrometer (Varian, AA240FS, USA). | Adult female brick kiln workers and controls | 25 kilns, 118 workers (18–55 years); 114 controls (18–58 years) | 1 sample/participant | NA | Metals (Cr, Cd, Ni) | Exposure biomarkers |
| David 2021[101] | Pakistan | Jun 2018 - Jun 2019 | Metal concentrations in whole blood measured using an AA 40 FS Fast Sequential Atomic Absorption Spectrometer (Varian, AA240FS, USA). | Children living at kiln sites | 25 kilns, 134 children (4 - 17 years old) | 1 sample/child | NA | Metals (Pb, Cd, Zn, Ni) | Exposure biomarkers |
| David 2022[9] | Pakistan | Mar - Nov 2018 | Metal concentrations in whole blood measured using a Fast Sequential Atomic Absorption Spectrometer (Varian, AA240FS, USA). | Adult male brick kiln workers and controls | 25 kilns, 346 workers (18–55 years; working for at least 1–5 years); 200 controls | 1 sample/participant | NA | Metals (Cr, Cd, Zn, Ni) | Exposure biomarkers |
| Flores-Ramirez 2018[33] | Mexico | 2010-2012 | 1-OHP in urine quantified using High Performance Liquid Chromatography (HPLC; HP1100, Agilent Technologies), as a biomarker of exposure to PAHs. Urinary trans, trans-muconic acid (t,t-MA) quantified using HPLC, as a biomarker of benzene exposure. Manganese and fluoride in urine determined using a Perkin-Elmer 3110 atomic absorption spectrophotometer and a potentiometric method, respectively. Lead in blood measured using a Perkin- | Children in area with brick kilns | 1 site, 40 children (6 - 12 years old) | 1 sample/child | NA | As, Mn, Pb, F-, PAHs (1-OHP), benzene (t,t-MA) | Exposure biomarkers |

| | | | | | | | | | |
|---|---|---|---|---|---|---|---|---|---|
| | | | Elmer 3110 atomic absorption spectrophotometer. | | | | | | |
| Goetz 2018[34] | Nepal | Apr 2015 | Mass, composition (organics, sulfates, nitrates, chlorides, and ammonium) and size distribution of $PM_1$ measured using a mini aerosol mass spectrometer (mAMS; Aerodyne Research Inc.). Magee Scientific AE33 aethalometer used to measure BC concentrations, absorption coefficients, and absorption Ångström exponents. Gases measured using a Picarro cavity ring-down spectrometer (CRDS) model G2401 (CO, $CO_2$, CH4), a LI-COR $CO_2$ and H2O monitor (Li840A), a Vaisala $CO_2$ monitor (GMP343), and a Gaslab Inc. high-range $CO_2$ monitor. | Flue gas and fugitive emissions | 1 CK, 1 FDZ | 1 sample/kiln | 4-hr sampling | $PM_1$ and components (BC, organics, sulfate, nitrate, ammonium, chloride, PAHs) | Emission factors; Size distributions |

| | | | | | | | | | |
|---|---|---|---|---|---|---|---|---|---|
| Gupta 2023[35] | India | Jun 2018 - May 2019 | 24-hr integrated $PM_{2.5}$ and $PM_{10}$ samples collected using two medium-volume samplers (URG 3000ABC, USA) at two sites in the city. Water-soluble inorganic ions quantified by ion chromatography (Thermo Scientific, Dionex-Aquion). Carbon fractions quantified using Desert Research Institute (DRI) Thermal/-Optical carbon analyzer (Model 2015). Elements analyzed using a dispersive X-Ray Fluorescence (ED-XRF) spectrometer. US EPA PMF v5.0 model used to apportion PM mass. | 2 sites in Ghaziabad | 2 sites | every 3rd day over 1 year | 24-hr sampling | $PM_{2.5}$, $PM_{10}$ | Source contributions |
| Hamid 2023[96] | Pakistan | Aug 2020 - Jan 2021 | TSP measured using a calibrated Microdust Pro Casella Cell (CEL-712) at 4 points in each brick kiln. Concentration of heavy metals in soil determined using atomic absorption spectrometry (Buck model 210 VGP). | Kiln site | 2 FCBTK, 2 ZZK | 2 per day, 3 times a month | NA | TSP, metals | Pollutant concentrations |
| Haque 2018[36] | Bangladesh | NA | Gravimetric samples of $PM_{2.5}$ collected on quartz fiber filters. Filters analyzed for BC using a Magee Scientific OT-21 Soot scan Transmissometer. $CO_2$, CO, $SO_2$, VOC, $NO_x$, and O3 measured by low volume (0.5 LPM) Aeroqual 500 real-time sampler through an electro-chemical gas sensor. | Stack | 10 FCBTK, 6 ZZK, 2 HK | NA | NA | $PM_{2.5}$, BC, $CO_2$, CO, $SO_2$, $NO_x$, VOCs | Pollutant concentrations; Emission factors |

| | | | | | | | | | |
|---|---|---|---|---|---|---|---|---|---|
| Hu 2019[37] | China | NA | $SO_2$, $NO_x$ and PM analyzed by a Flue gas analyzer (Laoying-3012H, Qingdao LaoYing Environmental Science and Technology, Co., Ltd.) installed at outlet of Flue gas. VOCs collected and analyzed by a GC-MS system. | Flue gas | 34 brick manufacturing companies | NA | NA | $SO_2$, $NO_x$, PM, VOCs | Emission factors |
| Hussain 2022[38] | Pakistan | NA | Stack emissions of $NO_x$, $SO_2$ and CO measured with a gas analyzer PG-250 (Horiba, Japan). | Stack | 3 kilns | NA | NA | $NO_x$, $SO_2$, CO | Pollutant concentrations |
| Iqbal 2013[39] | Pakistan | NA | Soot samples collected from brick kiln stacks. PAHS extracted via the soxhlet extraction method. | Stack | 5 kilns | 1 sample/kiln | NA | PAHs | Pollutant concentrations |
| Jahan 2016[102] | Pakistan | Mar 2015 | Metal concentrations in whole blood measured using an AA 40 FS Fast Sequential Atomic Absorption Spectrometer (Varian, AA240FS, USA). | Workers in different exposure groups, nonworkers living in the same area | NA | 1 sample/participant | | Metals (Pb, Cd, Cr) | Exposure biomarkers |
| Jan 2014[40] | Pakistan | NA | Soil samples collected from brick kilns and at various distances in four directions. PAHs extracted using soxhlet extraction and analyzed with a reversed phase high-performance liquid chromatograph (PerkinElmer, USA). | Kiln site | 1 kiln | NA | NA | PAHs | Pollutant concentrations |

| | | | | | | | | | |
|---|---|---|---|---|---|---|---|---|---|
| Jayarathne 2018[41] | Nepal | 11 – 25 Apr 2015 | $PM_{2.5}$ collected using a custom-built, dual-channel PM sampler on both Quartz and Teflon filters. $PM_{2.5}$ determined from Teflon filters. OC and EC determined following the NIOSH 5040 method on 1.0 cm2 punches of Quartz fiters (Sunset OC-EC Aerosol Analyzer, Sunset Laboratories, Tigard, OR). Inorganic ions quantified in aqueous extracts of filter samples by ion exchange chromatography with conductivity detection (Dionex-ICS 5000). Filter extracts analyzed for metals using a Thermo X-Series II quadrupole inductively coupled plasma mass spectrometry (ICP-MS) instrument (Thermo Fisher Scientific Inc., Waltham, MA, USA). Filter extracts analyzed for organic species using gas chromatography (GC; Agilent Technologies 7890A) coupled to mass spectrometry (MS; Agilent Technologies 5975). | Flue gas, 2–3 m downwind of the stack; fugitive emissions | 1 FDZ, 1 CK | 3 samples/kiln | NA | $PM_{2.5}$, EC, OC, metals, water-soluble inorganic ions, PAHs | Emission factors |
| Joshi 2008[103] | Nepal | Jun 2004 - Sep 2005 | Gravimetric sampling of TSP and $PM_{10}$. $SO_2$, $NO_x$, CO measured using a manual gas analyzer (Drager pump). | Areas with brick kilns (stationary & mobile) | NA | 2 samples/site | 4-hr sampling | TSP, $PM_{10}$, $SO_2$, $NO_x$, CO | Source contributions |
| Kamal 2014a[104] | Pakistan | NA | Soil dust samples collected from brick kiln units. Extracted samples analyzed by gas chromatography-mass spectrometer. | Six locations in the inner area of brick kiln units | 26 brick kilns | 1 sample/kiln | NA | PAHs | Pollutant concentrations |
| Kamal 2014b[105] | Pakistan | NA | Post-shift urine samples analysed for PAH biomarkers using high pressure liquid chromatography (HPLC-FD, XS PD-20A VP-Shimadzu) | Male brick kiln workers and non-occupationall | 46 workers, 34 controls | 1 sample/participant | NA | PAHs (1-OHP, α-naphthol, β-naphthol) | Exposure biomarkers |

| | | | | | | | | | |
|---|---|---|---|---|---|---|---|---|---|
| | | | | y exposed control group | | | | | |
| Kamal 2016[42] | Pakistan | NA | Polyurethane foam (PUF) passive air samplers used. PAHs extracted from the PUF disks using Soxhlet extraction assembly, and further concentrated using rotary evaporator and purified on a column, packed with alumina/silica. Analysis of PAHs carried out using a gas-chromatograph equipped with a mass-spectrometer (GC-MS). | Residential, industrial/agricultural, and industrial/traffic areas | 3 sites | NA | 56-day sampling period | PAHs | Source contributions |
| Khan 2019[95] | Pakistan | May 2016 - Apr 2017 | Seasonal data collected from both affected and controlled areas during non-crushing period, when only brick kilns were operational, and crushing period, when both sugar mills and brick kilns were operational. Gravimetric samples of $PM_{10}$ collected on glass fiber filters using High volume Air Sampler (VOPV-12). | Areas near (affected) and far (controlled) from brick kilns and sugar mills | NA | NA | NA | $PM_{10}$, $SO_2$, $NO_x$ | Source contributions |
| Khanoranga 2019[43] | Pakistan | Aug-Sep 2017 | Fluoride concentration in urine determined using F-ion selective electrode. | Brick kiln workers | 3 districts, 100 male brick kiln workers (17-45 years old) | 1 sample/worker | NA | F- | Exposure biomarkers |

| | | | | | | | | | |
|---|---|---|---|---|---|---|---|---|---|
| Kim 2015[44] | Nepal | Dec 21 2012-Jan 3 2013 (1st period), Feb 13-21 2013 (2nd period) | $PM_{10}$ gravimetric samples collected using a sequential sampler (PMS-103, APM Engineering). Trace elements analyzed using energy-dispersive X-ray fluorescence following the USEPA IO-3.6 method. Ions analyzed by ion chromatography following the USEPA IO-4.1 method. Quartz filters analyzed for OC and EC by a thermal optical transmittance carbon analyzer following the USEPA NIOSH-5040 method. Source contributions quantified using a the multivariate receptor model SMP (Solver for Mixture Problem). | Site in Bode, 15 m above ground | 1 site | NA | 24-hr sampling | $PM_{10}$, OC, EC | Source contributions |
| Le 2010[45] | Vietnam | 24–31 Jan 2007 | PM from stack gas collected on glass fiber filters using the STL-Combi dust sampler. CO and $SO_2$ measured using an automatic gas analyzer (Quintox, KM9006). | Stack | 1 kiln, 3 sites | NA | 1 hr/day for 7 days | CO, $SO_2$, PM | Pollutant concentrations; Emission factors |
| Love 1999[106] | England & Scotland | NA | Typical concentrations of repirable dust and silica in 12 occupational groups determined based on personal sampling supplemented by static samples in areas where people passed through but did not carry out specific tasks. Employees carried personal dust samplers (Casella cyclone). Quartz content derived by infrared spectrophotometry, or in some cases by x-ray diffraction, or by the potassium bromide disc method of infrared spectroscopy. | Workers in 12 different exposure groups at 18 Heavy clay factories | 1407 workers | NA | NA | RSP, silica | Personal exposures |

| | | | | | | | | | |
|---|---|---|---|---|---|---|---|---|---|
| Martinez-Salinas 2010[46] | Mexico | NA | 1-OHP in urine quantified using High Performance Liquid Chromatography (HPLC; HP1100, Agilent Technologies), as a biomarker of exposure to PAHs. | Children in area with brick kilns | 2 communities, 65 children (3 - 13 years old) | 1 sample/child | NA | PAHs (1-OHP) | Exposure biomarkers |
| Mondal 2017[47] | India | NA | Brick kiln bottom ash (BKBA) samples collected from 32 BKBA dumping sites in India and analysed for toxic metals using energy dispersive X-ray spectroscopy. | Brick-kiln bottom ash dumping sites | 20 sites in West Bengal, 12 sites in Assam | NA | NA | Metals (Pb, Cd, Zn, Cr, Mn, Cu) | Pollutant concentrations |
| Nasim 2020[15] | Pakistan | Apr-May 2018 | PM samples obtained in an isokinetic manner following US EPA Method 17, and measured gravimetrically. Real time monitoring of gaseous emissions carried out using the Horiba PG-350E gas analyzer. | Stack, 10-12m above ground | 1 FDZ, 1 BTK | 1 sample/site | 1-hr sampling (ZZK), 30-min feeding and 45-min non-feeding sampling (BTK) | $SO_2$, CO, $NO_x$, $CO_2$, PM | Emission factors |
| Nasir 2021[7] | Pakistan | Apr 2019 | $PM_{2.5}$ and $PM_{10}$ measured at one "treated" and one "control" site each in the Umar, Pandu and Badaber areas of the Peshawar district using the HAZ Dust Environmental Particulate Air Monitoring Equipment with Model (EPAM-5000). | Sites within 3km radius of kilns (treated) and sites beyond 3km radius (control) | 3 treated sites; 3 control sites | NA | NA | $PM_{2.5}$, $PM_{10}$ | Source contributions |

| | | | | | | | | | |
|---|---|---|---|---|---|---|---|---|---|
| Nepal 2019[6] | Nepal | Apr 29-May 5 2017, May 24-Jun 1 2017 | Real-time concentrations of $CO_2$, CO, $SO_2$ and BC measured using the Ratnoze1 (Mountain Air Engineering. USA) portable sampling system. PM mass determined gravimetrically using filter papers. BC measured using a microAeth (AE-51, AethLabs, CA), installed inside the Ratnoze1. | Stack, 10-15m above ground | 4 FCBTK, 3 FDZ | NA | 6-hr sampling | $CO_2$, $SO_2$, BC, $PM_{2.5}$ | Pollutant concentrations; Emission factors |
| Ortinez-Alvarez 2018[48] | Mexico | NA | $PM_{2.5}$ samples collected following reference method RFPS-0498-116 with a BGI Incorporated PQ200 Air Sampler and sharp-cut cyclone. CO and CH4 were measured with a spectrophotometer (Gasmet Technologies Oy, Model DX4000, www.gasmet. com) following, respectively, US-EPA method NSPS RM 3A and American Society for Testing and Materials (ASTM) method D 6348-03. Quartz filters containing $PM_{2.5}$ analyzed for OC and EC by the coulombimetry method. Personal exposures to CO and $PM_{2.5}$ measured with a Langan T15 CO meter using an electrochemical cell and a personal impactor (MSP Corp. Model 200) respectively. | Flue gas, fugitive emissions, brick kiln workers | 1 TFK, 1 MK2, 2 workers/kiln | NA | 2-hr sampling in kilns, 3-4 hr sampling for personal exposures | $PM_{2.5}$, OC, EC, CO | Pollutant concentrations; Emission factors; Personal exposures |

| | | | | | | | | | |
|---|---|---|---|---|---|---|---|---|---|
| Pangtey 2004[49] | India | NA | Sampling of work environment for airborne dust done using a personal dust sampler (Rotheroe and Michell Ltd), at 1-2 L/min depending on dust concentration. Respirable fraction collected using cyclone. $SO_2$, $NO_x$ and CO measured by data analogger (model 190, Data logger). Soil samples analysed for heavy metals using Atomic Absorption Spectrophotometer Perkin Elemer-5000 and for silica using gravimetric method. | Kiln site | 18 BTK, 4 FCBTK | NA | NA | TSP, RSP, SO2, NOx, CO2, metals (Pb, Cd, Zn, Cr, Mn, Cu, Fe, Ni) | Pollutant concentrations |
| Perez-Maldonado 2019[50] | Mexico | 2016-2018 | 1-OHP in urine quantified using High Performance Liquid Chromatography (HPLC; HP1100, Agilent Technologies), as a biomarker of exposure to PAHs. | Children in area with brick kilns | 1 site, 30 children (6 - 12 years old) | 1 sample/child | NA | PAHs (1-OHP) | Exposure biomarkers |
| Rajarathnam 2014[18] | India, Vietnam | NA | PM collected in Glass Fibre (GFC of Whatman make) thimbles and measured gravimetrically. Gaseous samples collected in impinger tube and analyzed for $SO_2$ following BIS/EPA method IS11255. CO and $CO_2$ measurements carried out using Flue gas analyzer. | Stack, 10-15m above ground | 17 kilns | ≥3 samples/kiln | NA | PM, $SO_2$, CO, $CO_2$ | Emission factors |
| Rauf 2022[51] | Pakistan | NA | Concentration of various pollutants measured in 3 different types of brick kilns and emission factors calculated. PM sampling conducted using an isokinetic sampling console-Apex− 572 following US EPA method 17. | Stack | 1 FCBTK, 1 IDZZK, 1 HK | alternate days over 2 weeks | 2-hr sampling | CO, $PM_{2.5}$, BC, $CO_2$, SO2 | Emission factors |

| | | | | | | | | | |
|---|---|---|---|---|---|---|---|---|---|
| | | | Stack emissions measured using Horiba PG-350E following US EPA method 2. | | | | | | |
| Ravankhah 2017[52] | Iran | NA | Surface soil samples collected from various sites in the vicinity of brick kilns and grouped by distance from the kiln. Cd concentration determined by graphite furnace atomic absorption spectroscopy (GFAAS, Shimadzu AA-670G, Japan). Pb, Ni, Zn and Cu concentrations determined by flame atomic absorption spectroscopy (FAAS, Shimadzu AA-670G, Japan). | Sites within 0-500 m from brick kiln | 12 sites | 5 samples/site | NA | Metals (Pb, Cd, Zn, Ni, Cu) | Pollutant concentrations |
| Raza 2014[107] | Pakistan | Fall 2009 – Spring 2010 | $PM_{2.5}$ and $PM_{10}$ measured using the dust monitor, DUST TRAK PRO II model 8530. | Different regions of brick kilns (modulation and loading, burning, and unloading sections) | NA | NA | NA | $PM_{10}$, $PM_{2.5}$ | Pollutant concentrations |
| Raza 2021[108] | Pakistan | Jan - Apr 2018 | Real-time monitoring of $PM_{2.5}$, $PM_{10}$ using a dust particle counter (Dylos DC-1700). $NO_2$, $SO_2$, VOCs measured using a portable gas sampler (Aeroqual-500). | Working sites (modulation area and near burning sites of kiln area) of brick kilns | 3 kilns | NA | 8-hr sampling | $PM_{2.5}$, $PM_{10}$, $NO_2$, $SO_2$, VOC | Pollutant concentrations |
| Rokni 2016[53] | Iran | NA | Respirable dust samples collected on PVC filters using a personal sampling pump (Apex Pro personal sampling pump, Casella, UK) to sample air through a size-selective cyclone (Respirable Dust Aluminum Cyclone, SKC, UK) at a flow rate of 2.2 L per min, following NIOSH Method 7602. Filters then analysed for silica | Workers at 2 brick kilns | 12 workers | 1 sample/worker | < 8-hr sampling | Silica | Personal exposures |

| | | | | | | | | | |
|---|---|---|---|---|---|---|---|---|---|
| | | | by IR spectroscopy (FT-IR, Perkin Elmer, Boston, USA). | | | | | | |
| Saikia 2018[54] | India | NA | Gravimetric $PM_{2.5}$ samples collected using a Fine Particulate Sampler (APM-550, Envirotech, India). Gravimetric $PM_{10}$ and TSPM samples collected using a Respirable Dust Sampler (APM 460 NL, Envirotech, India) and a High Volume Sampler (Envirotech APM430), respectively. Gas samples collected by an Impinger Gas Sampler (Ecotech, India) using the modified West and Gaeke method for $SO_2$, sodium arsenite for $NO_2$ and the Weather burn method for NH3.PM samples analyzed for ions using an Ion Chromatography system (882 Compact Ion chromatography, Metrohm AG), and trace elements using an Atomic Absorption Spectrophotometer (AAnalyst-700, Perkin Elmer). PAHs extracted in a Soxhlet extractor and analyzed in a Shimadzu HPLC–PDA system attached to a chromatographic column. | Kiln site | 1 kiln | NA | 8-hr sampling | $PM_{2.5}$, $PM_{10}$, TSP, $SO_2$, $NO_2$, NH3, PAHs, elemental concentrations in $PM_{2.5}$ and $PM_{10}$ | Pollutant concentrations |
| Sanjel 2016[109] | Nepal | Jan - Mar 2015 and Mar - Apr 2016 | Direct-reading PM concentrations determined using the Dusttrak model 8533. | 5 work stations: green brick molding, green brick stacking, red brick loading, coal | 16 kilns | NA | 2-hr sampling | TSP, $PM_{10}$, PM4, $PM_{2.5}$, $PM_1$ | Pollutant concentrations |

| | | | | | | | | | |
|---|---|---|---|---|---|---|---|---|---|
| | | | | crushing/carrying and firing | | | | | |
| Sanjel 2017[94] | Nepal | Feb-Mar 2015 | Personal samples collected on PVC filters following NIOSH Method 0500 for TSP and NIOSH Method 0600 for RSP, using SKC AirChek 52 air sampling pumps. | Workers in different exposure groups at 16 brick kilns | 72 workers (RSP), 89 workers (TSP) | NA | 120-min sampling for TSP, 160-min sampling for RSP | TSP, PM4 | Personal exposures |
| Sanjel 2018[19] | Nepal | Feb-Mar 2015 | Personal silica samples collected on PVC filters following NIOSH Method 7500, using SKC AirChek 52 (SKC Inc., Eighty Four, PA, USA) sampling pumps and aluminum cyclones with a 4 μm cut-point. | Workers in different positions at 16 brick kilns | 46 workers | 1 sample/worker | 170±10 min sampling | silica | Personal exposures |
| Sarkar 2017[55] | Nepal | Dec 19 2012-Jan 30 2013 | NMVOC measurements performed using a high-sensitivity proton-transfer-reaction time-of-flight mass spectrometer (PTR-TOF-MS model 8000, Ionicon Analytik GmbH, Innsbruck, Austria). Source contributions to NMVOCs estimated using a positive matrix factorization model (US EPA PMF v5.0). | Suburban site in Bhaktapur (Bode) | 1 site | 1006 samples | hourly averaged | NMVOCs | Source contributions |
| Stockwell 2016[56] | Nepal | Apr 2015 | Trace gas and aerosol measurements by Fourier transform infrared (FTIR) spectroscopy. The FTIR system included side-by-side Teflon and quartz fiber filters preceded by cyclones to collect gravimetric samples of $PM_{2.5}$. Real-time absorption and scattering resulting from BC and (indirectly) BrC, monitured using two photoacoustic extinctiometers (PAX, Droplet Measurement Technologies, Inc., CO) at 405 and 870 nm. | Flue gas and fugitive emissions | 1 ZZK, 1 CK | NA | 5-hr sampling | $CO_2$, CO, $SO_2$, NO, $NO_2$, VOCs, NH3, HCl, HF, HCN, BC, BrC | Emission factors |

| | | | | | | | | | |
|---|---|---|---|---|---|---|---|---|---|
| Sughis 2014[57] | Pakistan | Jan-Apr 2009 | $PM_1$, $PM_{2.5}$ and $PM_{10}$ concentrations estimated using a portable laser-operated aerosol mass analyzer (Aerocet 531, Met One Instrument Inc., USA). Concentrations of metals in urine samples quantified by inductively coupled plasma-mass spectrometry (Agilent 7500 ce instrument). | Outdoors on kiln site; brick kiln workers | 1 kiln; 80 brick kiln workers (8-12 years old) | NA; 1 sample/child | ≥24 hours at kiln | $PM_1$ , $PM_{2.5}$, $PM_{10}$; metals | Pollutant concentrations; Exposure biomarkers |
| Suksuwan 2023[58] | Thailand | NA | Concentrations of CO, $NO_x$ and $SO_2$ emitted from the smoke hole and chimneys of a beehive kiln measured using a flue gas analyzer (Testo 350, German). | Flue gas | 1 DDK | 3 firing stages | NA | CO, $NO_x$, $SO_2$ | Pollutant concentrations |
| Tabinda 2019[59] | Pakistan | NA | Concentrations of gaseous emissions (CO, $SO_2$, NO, $NO_2$, O2) measured using a portable exhaust gas analyzer (TESTO-350). | Stack and Flue gas | 2 BTK (1 using good quality coal, 1 Low-quality coal with wood) | 3 samples/location | NA | CO, $NO_x$, NO, $NO_2$, $CO_2$, $SO_2$, He | Pollutant concentrations |
| Tandon 2017[114] | India | 2010-2012 | Assessment of the ambient air quality of the brick kilns carried out using a portable air monitoring apparatus (Lamotte, USA). | Brick kiln site | NA | NA | NA | TSP, $SO_x$, $NO_x$, CO, $CO_2$ | Pollutant concentrations |
| Thygerson 2019[60] | Nepal | May 2018 | Gravimetric $PM_{2.5}$ samples collected on Teflon filters using SKC AirLite sampling pumps (SKC nc., Eighty Four, PA) and personal exposure monitors (PEM, SKC) with a cut-point of 2.5 µm. Optical analysis of samples to determine concentration of BC and BrC. Samples analysed for 33 elements following the IO3.3 compendium method, using the ARL energy-dispersive X- | Inside and outside homes located on kiln site, ~100m away from kiln | 16 indoor, 16 outdoor | 1 sample/location | NA | $PM_{2.5}$, BC, BrC, elemental concentratio ns in $PM_{2.5}$ | Pollutant concentrations |

| | | | | | | | | | |
|---|---|---|---|---|---|---|---|---|---|
| | | | ray fluorescence (EDXRF) instrument (Thermo Fisher Scientific, Waltham, MA, USA). | | | | | | |
| Ubaque 2010[61] | Colombia | NA | Gravimetric samples of PM collected on a fibre-glass filter. $SO_2$ and $NO_x$ measured following US EPA Methods 8 and 7, respectively. Samples analyzed for organic compounds by gas chromatography. Hydrochloride and hydrofluoride acids analyzed by ionic chromatography following US EPA Method 26A. US EPA Method 29 used for polychlorinated dibenzo dioxins and furans. Samples analyzed for metals by atomic absorption spectrometry. | 6 measuring points along Hoffman kiln vault | 1 kiln | 3 tests, 6 measuring points/test | 24-hr sampling | PM, $SO_2$, SO3, $NO_x$, metals, HF, HCl, VOCs ($CH_4$), polychlorinated dioxins, furans | Pollutant concentrations |
| Vaidya 2015[110] | India | 2008-2010 | CO, $SO_2$ and $NO_2$ measured using a Gasteck Detector Pump model 800. Dust measured gravimetrically with an air sampling pump (SKC Air Check–52). | Close to the kiln site and 35-40 feet away from site | 6 kilns | 2 samples/day, 3 days/season (summer & winter)*<br><br>*Different values reported in Table 3 | NA | CO, $NO_2$, $SO_2$, PM | Pollutant concentrations |
| Wang 2022[62] | China | Apr - Oct 2020 | $PM_{2.5}$ samples collected on pre-fired quartz-fiber filters using medium-volume samplers (HY-100SFB, Qingdao, China) at 100 L/min. Background values in surrounding area measured and subtracted from kiln concentrations. Elements on filters quantified using inductively coupled plasma-mass spectroscopy (ICP-MS) (NexION 350D, PerkinElmer, Shelton, Connecticut, USA). PAHs analyzed using a | Flue gas | 4 brick production enterprises | 1 sample/kiln | 4-5 hr sampling | $PM_{2.5}$, PAHs, elements | Pollutant concentrations |

| | | | | | | | | | |
|---|---|---|---|---|---|---|---|---|---|
| | | | thermal desorption-gas chromatography-mass spectrometer (TD-GC–MS) (7980GC/5975MS; Agilent Technology, Santa Clara, CA, USA). | | | | | | |
| Werden 2022[112] | Nepal | 13 - 24 Apr 2015 | Concentration and chemical composition of ambient nonrefractory $PM_1$ (organic aerosol, sulfate, nitrate, chloride, and ammonium) measured by a mini aerosol mass spectrometer (mAMS). Analyzed using PMF to differentiate sources. Simultaneous measurements of brown carbon and BC (Aethalometer AE 33, Magee Scientific, USA). BC assumed to be submicron. | Suburban site in Kathmandu Valley (Bode) | 1 site | NA | Continuous, 1-min intervals | $PM_1$ and components (organics, sulfate, nitrate, ammonium, chloride) | Source contributions |
| Werden 2023[113] | Nepal | Jan - Feb 2018 | Concentration and chemical composition of ambient nonrefractory $PM_1$ (organic aerosol, sulfate, nitrate, chloride, and ammonium) measured by a mini aerosol mass spectrometer (mAMS). Analyzed using PMF to differentiate sources. Colocated gravimetric samples of $PM_{2.5}$ collected twice a day for 11-hr sample times analyzed for BC using a dual-spot aethalometer (AE33, Magee Scientific). BC measured assumed to be predominantly submicron and total $PM_1$ computed as the mass of nonrefractory $PM_1$ from the mAMS plus BC from the aethalometer. | Rural, urban and suburban sites in Kathmandu Valley | 3 sites | NA | NA | $PM_1$ and components (organics, sulfate, nitrate, ammonium, chloride) | Source contributions |

| | | | | | | | | | |
|---|---|---|---|---|---|---|---|---|---|
| Weyant 2014[63] | India, Vietnam | 2011-2012 | CO and $CO_2$ concentrations measured in real-time using an electrochemical sensor (SS1128, Senko) and a nondispersive infrared sensor (Telaire T6615, GE), respectively. $PM_{2.5}$ collected on Teflon filters using a 2.5 µm cut cyclone (URG 2000−30ED). EC and OC collected on quartz fiber filters and measured using thermal-optical analysis with a Sunset Laboratory OC/ EC analyzer. | Stack | 1 DDK, 3 BTK, 2 VSBK, 3 NDZ, 3 FDZ, 1 TK | 3 samples/kiln | 20-60 min sampling | CO, $PM_{2.5}$, EC | Emission factors |
| Ying 2021[64] | China | NA | Pollutants in exhaust gas measured using a flue gas sampler (M5, KNJ Engineering, Japan). PCDD/Fs in samples analyzed by high-resolution gas chromatography/high-resolution mass spectrometry (JMS-800D, JEOL, Japan). | Flue gas | 1 TK | 8/9 samples | 1-hr sampling | PM, $SO_2$, $NO_x$, HCl, Fluorides, $H_2S$, $NH_3$, PCDD/Fs | Pollutant concentrations; Emission factors |
| Zavala 2018[65] | Mexico | 12-16 Mar 2013 | $PM_{2.5}$ samples collected on quart filters using an inertial mass separator with a cut-point of 2.5 µm. EC and OC composition analyzed using thermal-optical transmittance (TOT) and reflectance (TOR) analysis. Analyses of anions and cations by ion chromatography. $CO_2$ and CO measured using a Fourier-transform infrared spectrometer (FTIR). Flame ionization detector (FID) analyzer used to measure total gaseous organic compounds (TOGs). | Flue gas and fugitive emissions | 1 TFK, 1 TCK, 1 MK2 | NA | NA | $CO_2$, CO, NO, $NO_2$, $SO_2$, $PM_{2.5}$, BC, OC,VOCs, inorganic components | Emission factors |
| Zawilla 2014[111] | Egypt | NA | Total dust samples collected over 8-hr period at 14 L/min from 2 sites each in the production, mining and control | Production and mining areas of | 1 brick factory | 48 samples | ≥ 8-hr sampling | TSP; RSP, silica | Pollutant concentrations; Personal exposures |

| | | | | | | | | | |
|---|---|---|---|---|---|---|---|---|---|
| | | | areas. Respirable dust samples collected with personal sampler devices attached to the belts and collars of workers. Pumps operated at 1.7 L/min for at least 8 hours. Free silica content of the collected dust determined by X-ray diffraction (Philips X-ray diffraction equipment model PW/1710). | factory; workers | | | | | |
| Zhang 2020[66] | China | Sep−Oct 2014 | Particle sizes and mixing state of BC obtained using a single-particle soot photometer (SP2, Droplet Measurement Technologies, Boulder, CO). | Flue gas | NA | NA | 0.5-2 hr sampling | Count and mass median diameters of BC | Size distributions |
| Zhou 2014[67] | China | NA | Coal gangue, brick, fly ash and fuel gas samples collected from the molding room, products after firing, dust-collector and stack, respectively. Flue gas samples collected following American Standard Method EPA 29. Trace elements collected in impingers and particulates obtained in a filter. Samples analyzed for Hg and other elements by cold vapor atomic absorption spectroscopy (CVAAS) and inductively coupled plasma mass spectrometry (ICP-MS), repsectively. Concentration of 238U, 232Th, 226Ra, 210Pb and 40K coal gangue, brick, fly ash, and particulates in Flue gas samples determined by a high-purity germanium gamma ray spectrometer. Alpha activity of 210Po counted by alpha spectrometry. | Stack | 1 coal gangue brick making plant | NA | 1-hr sampling | Elemental concentrations in coal gangue and its combustion products, 238U, 232Th, 226Ra, 210Pb, 210Po, 40K, 222Rn | Pollutant concentrations; Radionuclide activity |

| | | | | | | | | | |
|---|---|---|---|---|---|---|---|---|---|
| Zhou 2015[68] | China | NA | Simulation experiment conducted in fixed bed reactor using coal, coal gangue, coal ash and clay samples from 3 tunnel kilns. Flue gas sampled following the Ontario Hydro Method. Flue gas samples and combustion residues analyzed for As, Cd, and Pb by inductively coupled plasma mass spectrometry (ICP-MS) and for Hg by cold vapour atomic absorption spectrometry (CV-AAS). | Stack | 3 TK | NA | 1-hr sampling | As, Cd, Hg, Pb | Emission factors |

**Table S2. Quality control of studies reporting brick kiln pollution data.**

| Study | Methodology | | | | Results | Total score |
|---|---|---|---|---|---|---|
| | **Representativeness of sample** | **Ascertainment of exposure** | **Quality control** | **Overall quality** | | |
| | a) Truly representative of the average for the target exposure * (adequate sampling duration and number of samples)<br><br>b) Somewhat representative of the average in the target exposure *<br><br>c) Not representative of the average for the target exposure/No description of the sampling strategy | a) Validated measurement tools and detailed description of methods **<br><br>b) Some description of measurement tools and methods *<br><br>c) Inadequate description of measurement tools and methods | a) Detailed description of calibration and quality control *<br><br>b) Some description of calibration and quality control *<br><br>c) Inadequate description of calibration and quality control | a) High quality **<br><br>b) Moderate quality *<br><br>c) Low quality | a) High quality (results and statistical tests are clearly described and appropriate) **<br><br>b) Moderate quality (results and statistical tests are somewhat described and appropriate) *<br><br>c) Low quality (results and statistical tests are not appropriate, not described or incomplete) | |
| Abedin 2020[20] | b | a | b | b | b | 7 |
| Ahmad 2020[97] | b | a | a | a | c | 6 |
| Akinshipe 2018[21] | c | b | c | c | c | 1 |
| Akram 2022[98] | b | b | a | b | a | 6 |
| Arif 2018[22] | b | b | c | b | b | 4 |
| Bashir 2023[23] | c | b | c | c | c | 1 |
| Beard 2022[33]8 | b | a | b | a | a | 8 |
| Begum 2005[24] | b | b | c | b | b | 4 |
| Begum 2009[25] | b | a | b | b | a | 7 |
| Begum 2011[26] | b | a | b | b | a | 7 |
| Begum 2013a[27] | b | a | c | b | a | 6 |
| Begum 2013b[29] | b | a | b | b | a | 7 |
| Begum 2019[28] | b | a | b | b | a | 7 |
| Berumen-Rodriguez 2021[100] | c | b | c | c | c | 1 |
| Berumen-Rodriguez 2023[99] | c | b | b | b | c | 3 |
| Bruce 2007[30] | c | a | b | b | c | 4 |
| Chen 2017[16] | a | b | b | b | b | 5 |
| Christian 2010[31] | b | b | b | b | b | 5 |

| | | | | | | |
|---|---|---|---|---|---|---|
| Co 2009[32] | c | b | c | c | c | 1 |
| David 2020 | b | a | b | b | b | 6 |
| David 2021[101] | a | a | a | a | b | 7 |
| David 2022[9] | a | a | b | a | b | 7 |
| Flores-Ramirez 2018[33] | b | b | b | b | b | 5 |
| Goetz 2018[34] | a | a | b | a | a | 8 |
| Gupta 2023[35] | b | a | b | b | b | 6 |
| Hamid 2023[96] | b | b | c | b | b | 4 |
| Haque 2018[36] | c | a | c | b | b | 4 |
| Hu 2019[37] | c | a | a | b | b | 5 |
| Hussain 2022[38] | b | c | b | c | c | 2 |
| Iqbal 2013[39] | b | b | c | c | c | 2 |
| Jahan 2016[102] | c | b | b | b | b | 4 |
| Jan 2014[40] | b | b | b | b | b | 5 |
| Jayarathne 2018[41] | b | a | a | a | a | 8 |
| Joshi 2008[103] | b | b | c | b | c | 3 |
| Kamal 2014a[104] | a | a | a | a | a | 8 |
| Kamal 2014b[105] | b | a | a | a | a | 8 |
| Kamal 2016[42] | b | a | a | a | c | 6 |
| Khan 2019[95] | b | b | c | b | c | 3 |
| Khanoranga 2019[43] | b | a | a | b | b | 6 |
| Kim 2015[44] | b | a | c | b | a | 6 |
| Le 2010[45] | c | b | c | c | b | 2 |
| Love 1999[106] | b | b | c | b | b | 4 |
| Martinez-Salinas 2010[46] | a | a | c | b | b | 5 |
| Mondal 2017[47] | b | b | b | b | b | 5 |
| Nasim 2020[15] | c | a | b | b | b | 5 |
| Nasir 2021[7] | b | c | c | c | c | 1 |
| Nepal 2019[6] | b | a | b | b | a | 7 |
| Ortinez-Alvarez 2018[48] | c | a | c | b | c | 3 |
| Pangtey 2004[49] | b | b | c | b | c | 3 |
| Perez-Maldonado 2019[50] | c | b | c | c | b | 2 |
| Rajarathnam 2014[18] | b | a | c | b | b | 5 |
| Rauf 2022[51] | b | a | c | b | c | 4 |

| | | | | | | |
|---|---|---|---|---|---|---|
| Ravankhah 2017[52] | a | a | a | a | a | 8 |
| Raza 2014[107] | c | b | c | c | c | 1 |
| Raza 2021[108] | b | b | c | c | c | 2 |
| Rokni 2016[53] | b | a | c | b | b | 5 |
| Saikia 2018[54] | b | a | b | b | c | 5 |
| Sanjel 2016[109] | a | b | b | b | c | 4 |
| Sanjel 2017[94] | a | a | a | a | a | 8 |
| Sanjel 2018[19] | b | a | b | b | b | 6 |
| Sarkar 2017[55] | b | a | b | b | b | 6 |
| Stockwell 2016[56] | c | a | a | a | a | 7 |
| Sughis 2014[57] | b | b | c | b | b | 4 |
| Suksuwan 2023[58] | c | c | c | c | b | 1 |
| Tabinda 2019[59] | c | c | c | c | c | 0 |
| Tandon 2017[114] | c | c | c | c | c | 0 |
| Thygerson 2019[60] | c | b | b | b | a | 5 |
| Ubaque 2010[61] | c | b | b | b | b | 4 |
| Vaidya 2015[110] | b | b | c | c | c | 2 |
| Wang 2022[62] | b | a | b | b | a | 7 |
| Werden 2022[112] | b | a | a | a | a | 8 |
| Werden 2023[113] | b | a | a | a | a | 8 |
| Weyant 2014[63] | b | a | b | b | b | 6 |
| Ying 2021[64] | b | b | c | b | b | 4 |
| Zavala 2018[65] | b | a | a | a | b | 7 |
| Zawilla 2014[111] | b | b | b | b | c | 4 |
| Zhang 2020[66] | c | a | b | b | a | 6 |
| Zhou 2014[67] | b | a | b | b | b | 6 |
| Zhou 2015[68] | b | a | b | b | b | 6 |

**Table S3. Summary of pollution data reported in the studies.**

| | | |
|---|---|---|
| **Pollution data** | **n (%)** | **References** |
| Pollutant concentrations at kilns | 32 (41%) | 6,8,21,23,30,36,38–40,45,47–49,52,54,57–62,64,67,96,100,104,107–111,114 |
| Emission factors | 20 (25%) | 6,15,16,18,21,23,31,32,34,36,37,41,45,48,51,56,63–65,68 |
| Source contributions | 16 (21%) | 7,22,24–29,35,42,44,55,95,103,112,113 |
| Exposure biomarkers | 14 (18%) | 9,33,43,46,50,57,74,97–102,105 |
| Personal exposures | 6 (8%) | 19,48,53,94,106,111 |
| Particle size distribution | 3 (4%) | 30,34,66 |
| Radionuclide activity | 2 (3%) | 20,67 |
| **Pollutants** | | |
| PM | 48 (61%) | 6,7,15,16,18,21–32,34–37,41,44,45,48,49,51,54,57,60–65,94–96,100,103,107–114 |
| $SO_2$ | 24 (30%) | 6,8,15,16,18,21,23,32,36–38,45,49,51,54,56,58,59,61,64,65,95,103,108 |
| CO | 22 (27%) | 8,15,16,18,21,23,31,32,36,38,45,48,49,51,56,58,59,63,65,103,110,114 |
| BC | 14 (18%) | 6,22–26,28,34,36,51,56,60,65,66 |
| $CO_2$ | 14 (18%) | 6,8,15,16,18,21,23,31,36,51,56,59,65,114 |
| $NO_x$ | 13 (16%) | 15,21,36–38,49,58,59,61,64,95,103,114 |
| PAHs | 10 (13%) | 16,30,34,39–42,54,62,104 |
| VOCs | 8 (10%) | 31,36,37,55,56,61,65,108 |
| $NO_2$ | 8 (10%) | 8,16,21,41,54,59,65,108 |
| EC | 7 (9%) | 16,27,29,41,44,48,63 |
| OC | 6 (8%) | 16,28,41,44,48,65 |
| $SiO_2$ | 5 (6%) | 8,19,53,106,111 |
| NO | 5 (6%) | 16,21,56,59,65 |
| $NH_3$ | 4 (5%) | 31,54,56,64 |
| HCl | 4 (5%) | 31,56,61,64 |
| HF | 3 (4%) | 31,56,61 |
| BrC | 2 (3%) | 56,60 |
| $SO_3$ | 1 (1%) | 61 |
| HCN | 1 (1%) | 56 |
| **Measurement location** | | |
| Kiln sites | 60 (76%) | |
| Stack | 17 (22%) | 6,15,18,23,32,36,38,39,41,45,51,59,61,63,67,68,109 |

| | | | | |
|---|---|---|---|---|
| | | Locations on kiln site | 17 (22%) | [8,20,40,49,52,54,57,60,96,100,104,107–111,114] |
| | | Flue gas | 15 (19%) | [16,21,31,34,37,41,48,56,58,59,62,64–66,126] |
| | | Exposure biomarkers in BKWs | 10 (13%) | [9,43,57,74,97–100,102,105] |
| | | Personal exposures in BKWs | 6 (8%) | [19,48,53,94,106,111] |
| | | Exposure biomarkers in children living at kiln sites | 1 (1%) | [101] |
| | Outside kiln sites | | 19 (24%) | |
| | | Source apportionment | 12 (15%) | [22,24–29,35,44,55,112,113] |
| | | Exposure biomarkers among children living in communities near kilns | 3 (4%) | [33,46,50] |
| | | Pollutant concentrations regressed against brick production | 1 (1%) | [42] |
| | | Comparison of ambient pollution during brick kiln and pre-operational seasons | 1 (1%) | [103] |
| | | Comparison of PM levels within and beyond a 3km radius of kilns | 1 (1%) | [7] |
| | | Comparison of ambient pollution near and far from brick kilns and sugar mills, during the non-crushing period, when only brick kilns were operational, and the crushing period, when both sugar mills and brick kilns were operational | 1 (1%) | [95] |

**Table S4. Summary of type of pollutant emission factors and concentrations at brick kiln sites.**

| | | | |
|---|---|---|---|
| **Pollutant emission factors** | | **n (%)** | **References** |
| | PM | 18 (90%) | 3,9,10,31,37,51,55,56,69,73,76,78,82,83,85,89,90,99 |
| | $SO_2$ | 14 (70%) | 83,9,10,31,51,59,69,78,82,83,85,89,90,99 |
| | CO | 13 (65%) | 3,9,10,31,37,59,59,69,73,78,82,90,99 |
| | $CO_2$ | 11 (55%) | 3,9,10,31,51,59,69,73,78,82,99 |
| | BC | 7 (35%) | 3,10,51,55,59,69,78 |
| | VOCs | 5 (25%) | 29,34,35,54,63 |
| | EC | 4 (20%) | 37,56,76,82 |
| | OC | 4 (20%) | 56,76,78,82 |
| | $NO_2$ | 4 (20%) | 59,78,82,99 |
| | NO | 4 (20%) | 59,78,82,99 |
| | $NO_x$ | 4 (20%) | 9,83,85,99 |
| | PAHs | 3 (15%) | 28,32,39 |
| **Pollutant concentrations** | | **n (%)** | **References** |
| | PM | 22 (69%) | 1–17,17–21 |
| | $SO_2$ | 14 (44%) | 1–14 |
| | CO | 11 (34%) | 8,19,21,22,36,38,45,50,59,60,110 |
| | $NO_x$ | 9 (28%) | 1–9 |
| | $CO_2$ | 7 (22%) | 1–7 |
| | PAHs | 6 (19%) | 27,37,38,52,60,103 |
| | Metals | 6 (19%) | 27,45,47,50,59,96 |
| | $NO_2$ | 5 (16%) | 16,19,22,55,60 |
| | BC | 3 (9%) | 8,21,61 |
| | VOCs | 3 (9%) | 16,36,62 |
| | NO | 2 (6%) | 19,60 |
| | OC | 1 (3%) | 49 |
| | EC | 1 (3%) | 49 |
| | $SiO_2$ | 1 (3%) | 1 |

**Table S5. Emission factors of brick kiln pollutants reported in studies.**

| Study | Pollutant | Units | Brick kiln type | Fuels used | Fuel category | Measurement location | Sample size | Mean | SD | Notes |
|---|---|---|---|---|---|---|---|---|---|---|
| Akinshipe 2018 | $SO_2$ | g/brick | CK | Coal | Coal | Flue gas | 1778 | 1.07 | 0.66 | Based on hourly emission concentrations from Batches 3-5 and 8-12 |
| Akinshipe 2018 | $NO_2$ | g/brick | CK | Coal | Coal | Flue gas | 1778 | 0 | 0 | Based on hourly emission concentrations from Batches 3-6 and 8-12 |
| Akinshipe 2018 | NO | g/brick | CK | Coal | Coal | Flue gas | 1778 | 0.14 | 0.1 | |
| Akinshipe 2018 | $NO_x$ | g/brick | CK | Coal | Coal | Flue gas | 1778 | 0.14 | 0.1 | |
| Akinshipe 2018 | $PM_1$ | g/brick | CK | Coal | Coal | Flue gas | 1778 | 0.96 | 0 | |
| Akinshipe 2018 | $PM_{2.5}$ | g/brick | CK | Coal | Coal | Flue gas | 1778 | 0.96 | 0 | |
| Akinshipe 2018 | $PM_{10}$ | g/brick | CK | Coal | Coal | Flue gas | 1778 | 0.96 | 0.5 | |
| Akinshipe 2018 | CO | g/brick | CK | Coal | Coal | Flue gas | 1778 | 22.5 | 18.8 | |
| Akinshipe 2018 | $CO_2$ | g/brick | CK | Coal | Coal | Flue gas | 1778 | 378 | 223 | |
| Akinshipe 2018 | $SO_2$ | g/kg brick | CK | Coal | Coal | Flue gas | 1778 | 0.38 | 0.25 | |
| Akinshipe 2018 | $NO_2$ | g/kg brick | CK | Coal | Coal | Flue gas | 1778 | 0 | 0 | |
| Akinshipe 2018 | NO | g/kg brick | CK | Coal | Coal | Flue gas | 1778 | 0.05 | 0.03 | |
| Akinshipe 2018 | $NO_x$ | g/kg brick | CK | Coal | Coal | Flue gas | 1778 | 0.05 | 0.03 | |
| Akinshipe 2018 | $PM_1$ | g/kg brick | CK | Coal | Coal | Flue gas | 1778 | 0.33 | 0 | |
| Akinshipe 2018 | $PM_{2.5}$ | g/kg brick | CK | Coal | Coal | Flue gas | 1778 | 0.34 | 0 | |
| Akinshipe 2018 | $PM_{10}$ | g/kg brick | CK | Coal | Coal | Flue gas | 1778 | 0.34 | 0.5 | |
| Akinshipe 2018 | CO | g/kg brick | CK | Coal | Coal | Flue gas | 1778 | 7.83 | 6.65 | |
| Akinshipe 2018 | $CO_2$ | g/kg brick | CK | Coal | Coal | Flue gas | 1778 | 132 | 79.7 | |
| Bashir 2023 | CO | g/kg fuel | BTK | Coal | Coal | Stack | 1 | 24 | | |
| Bashir 2023 | $CO_2$ | g/kg fuel | BTK | Coal | Coal | Stack | 1 | 1630 | | |
| Bashir 2023 | $SO_2$ | g/kg fuel | BTK | Coal | Coal | Stack | 1 | 14.2 | | |
| Bashir 2023 | $PM_{2.5}$ | g/kg fuel | BTK | Coal | Coal | Stack | 1 | 5.4 | | |
| Bashir 2023 | BC | g/kg fuel | BTK | Coal | Coal | Stack | 1 | 0.37 | | |

| | | | | | | | | | | |
|---|---|---|---|---|---|---|---|---|---|---|
| Bashir 2023 | CO | g/kg fuel | ZZK | Coal | Coal | Stack | 1 | 13.8 | | |
| Bashir 2023 | $CO_2$ | g/kg fuel | ZZK | Coal | Coal | Stack | 1 | 1410 | | |
| Bashir 2023 | $SO_2$ | g/kg fuel | ZZK | Coal | Coal | Stack | 1 | 1.14 | | |
| Bashir 2023 | $PM_{2.5}$ | g/kg fuel | ZZK | Coal | Coal | Stack | 1 | 3.5 | | |
| Bashir 2023 | BC | g/kg fuel | ZZK | Coal | Coal | Stack | 1 | 0.06 | | |
| Chen 2017 | $CO_2$ | g/kg fuel | HK | Coal | Coal | Flue gas | 18 | 1940 | 260 | K-1 kilns (Xiamen) |
| Chen 2017 | CO | g/kg fuel | HK | Coal | Coal | Flue gas | 18 | 267 | 161 | K-1 kilns (Xiamen) |
| Chen 2017 | EC | g/kg fuel | HK | Coal | Coal | Flue gas | 18 | 0.002 | 0.003 | K-1 kilns (Xiamen) |
| Chen 2017 | OC | g/kg fuel | HK | Coal | Coal | Flue gas | 18 | 0.044 | 0.086 | K-1 kilns (Xiamen) |
| Chen 2017 | $SO_2$ | g/kg fuel | HK | Coal | Coal | Flue gas | 18 | 2.22 | 1.4 | K-1 kilns (Xiamen) |
| Chen 2017 | $NO_2$ | g/kg fuel | HK | Coal | Coal | Flue gas | 18 | 0.091 | 0.091 | K-1 kilns (Xiamen) |
| Chen 2017 | NO | g/kg fuel | HK | Coal | Coal | Flue gas | 18 | 8.27 | 6.25 | K-1 kilns (Xiamen) |
| Chen 2017 | $PM_{2.5}$ | g/kg fuel | HK | Coal | Coal | Flue gas | 18 | 0.605 | 0.957 | K-1 kilns (Xiamen) |
| Chen 2017 | $PM_{10}$ | g/kg fuel | HK | Coal | Coal | Flue gas | 18 | 1.25 | 1.44 | K-1 kilns (Xiamen) |
| Chen 2017 | TSP | g/kg fuel | HK | Coal | Coal | Flue gas | 18 | 1.87 | 1.77 | K-1 kilns (Xiamen) |
| Chen 2017 | $CO_2$ | g/kg fuel | HK | Coal | Coal | Flue gas | 10 | 1920 | 240 | K-2 kilns (Xi'an-Baoji-Weinan) |
| Chen 2017 | CO | g/kg fuel | HK | Coal | Coal | Flue gas | 10 | 395 | 160 | K-2 kilns (Xi'an-Baoji-Weinan) |
| Chen 2017 | EC | g/kg fuel | HK | Coal | Coal | Flue gas | 10 | 0.056 | 0.04 | K-2 kilns (Xi'an-Baoji-Weinan) |
| Chen 2017 | OC | g/kg fuel | HK | Coal | Coal | Flue gas | 10 | 0.143 | 0.121 | K-2 kilns (Xi'an-Baoji-Weinan) |
| Chen 2017 | $SO_2$ | g/kg fuel | HK | Coal | Coal | Flue gas | 10 | 1.99 | 1.06 | K-2 kilns (Xi'an-Baoji-Weinan) |
| Chen 2017 | $NO_2$ | g/kg fuel | HK | Coal | Coal | Flue gas | 10 | 0.019 | 0.039 | K-2 kilns (Xi'an-Baoji-Weinan) |
| Chen 2017 | NO | g/kg fuel | HK | Coal | Coal | Flue gas | 10 | 5.05 | 2.49 | K-2 kilns (Xi'an-Baoji-Weinan) |
| Chen 2017 | $PM_{2.5}$ | g/kg fuel | HK | Coal | Coal | Flue gas | 10 | 2.67 | 2.62 | K-2 kilns (Xi'an-Baoji-Weinan) |
| Chen 2017 | $PM_{10}$ | g/kg fuel | HK | Coal | Coal | Flue gas | 10 | 3.09 | 1.63 | K-2 kilns (Xi'an-Baoji-Weinan) |
| Chen 2017 | TSP | g/kg fuel | HK | Coal | Coal | Flue gas | 10 | 4.57 | 2.35 | K-2 kilns (Xi'an-Baoji-Weinan) |

| | | | | | | | | | | |
|---|---|---|---|---|---|---|---|---|---|---|
| Chen 2017 | $CO_2$ | g/kg fuel | HK | Coal | Coal | Fugitive emissions | 8 | 578 | 11.9 | K-1 kilns (Xiamen) |
| Chen 2017 | CO | g/kg fuel | HK | Coal | Coal | Fugitive emissions | 8 | 26.9 | 7.67 | K-1 kilns (Xiamen) |
| Chen 2017 | EC | g/kg fuel | HK | Coal | Coal | Fugitive emissions | 8 | 0.04 | 0.03 | K-1 kilns (Xiamen) |
| Chen 2017 | OC | g/kg fuel | HK | Coal | Coal | Fugitive emissions | 8 | 0.63 | 0.36 | K-1 kilns (Xiamen) |
| Chen 2017 | $SO_2$ | g/kg fuel | HK | Coal | Coal | Fugitive emissions | 8 | 2.34 | 1.03 | K-1 kilns (Xiamen) |
| Chen 2017 | $NO_2$ | g/kg fuel | HK | Coal | Coal | Fugitive emissions | 8 | 0.01 | 0.01 | K-1 kilns (Xiamen) |
| Chen 2017 | NO | g/kg fuel | HK | Coal | Coal | Fugitive emissions | 8 | 0.72 | 0.85 | K-1 kilns (Xiamen) |
| Chen 2017 | $PM_{2.5}$ | g/kg fuel | HK | Coal | Coal | Fugitive emissions | 8 | 3.71 | 1.58 | K-1 kilns (Xiamen) |
| Chen 2017 | $PM_{10}$ | g/kg fuel | HK | Coal | Coal | Fugitive emissions | 8 | 8.36 | 3.16 | K-1 kilns (Xiamen) |
| Chen 2017 | TSP | g/kg fuel | HK | Coal | Coal | Fugitive emissions | 8 | 15.5 | 11.7 | K-1 kilns (Xiamen) |
| Chen 2017 | $CO_2$ | g/kg fuel | HK | Coal | Coal | Fugitive emissions | 5 | 403 | 92.7 | K-2 kilns (Xi'an-Baoji-Weinan) |
| Chen 2017 | CO | g/kg fuel | HK | Coal | Coal | Fugitive emissions | 5 | 123 | 48.7 | K-2 kilns (Xi'an-Baoji-Weinan) |
| Chen 2017 | EC | g/kg fuel | HK | Coal | Coal | Fugitive emissions | 5 | 0.16 | 0.21 | K-2 kilns (Xi'an-Baoji-Weinan) |
| Chen 2017 | OC | g/kg fuel | HK | Coal | Coal | Fugitive emissions | 5 | 1.66 | 1.88 | K-2 kilns (Xi'an-Baoji-Weinan) |
| Chen 2017 | $SO_2$ | g/kg fuel | HK | Coal | Coal | Fugitive emissions | 5 | 2.6 | 3.74 | K-2 kilns (Xi'an-Baoji-Weinan) |
| Chen 2017 | $NO_2$ | g/kg fuel | HK | Coal | Coal | Fugitive emissions | 5 | 0.02 | 0.01 | K-2 kilns (Xi'an-Baoji-Weinan) |
| Chen 2017 | NO | g/kg fuel | HK | Coal | Coal | Fugitive emissions | 5 | 1.33 | 1.46 | K-2 kilns (Xi'an-Baoji-Weinan) |
| Chen 2017 | $PM_{2.5}$ | g/kg fuel | HK | Coal | Coal | Fugitive emissions | 5 | 31.1 | 40.7 | K-2 kilns (Xi'an-Baoji-Weinan) |
| Chen 2017 | $PM_{10}$ | g/kg fuel | HK | Coal | Coal | Fugitive emissions | 5 | 71.5 | 93.8 | K-2 kilns (Xi'an-Baoji-Weinan) |
| Chen 2017 | TSP | g/kg fuel | HK | Coal | Coal | Fugitive emissions | 5 | 79.2 | 102 | K-2 kilns (Xi'an-Baoji-Weinan) |
| Christian 2010 | $CO_2$ | g/kg fuel | TFK | Biomass (mainly sawdust and crop waste) | Biomass | Flue gas | 1 | 1736 | NA | |

| | | | | | | | | | | |
|---|---|---|---|---|---|---|---|---|---|---|
| Christian 2010 | $CO_2$ | g/kg fuel | TFK | Biomass (mainly sawdust and crop waste) | Biomass | Flue gas | 1 | 1780 | NA | |
| Christian 2010 | $CO_2$ | g/kg fuel | TFK | Biomass (mainly sawdust and crop waste) | Biomass | Flue gas | 1 | 1787 | NA | |
| Christian 2010 | CO | g/kg fuel | TFK | Biomass (mainly sawdust and crop waste) | Biomass | Flue gas | 1 | 55.7 | NA | |
| Christian 2010 | CO | g/kg fuel | TFK | Biomass (mainly sawdust and crop waste) | Biomass | Flue gas | 1 | 30.2 | NA | |
| Christian 2010 | CO | g/kg fuel | TFK | Biomass (mainly sawdust and crop waste) | Biomass | Flue gas | 1 | 25.7 | NA | |
| Christian 2010 | $PM_{2.5}$ | g/kg fuel | TFK | Biomass (mainly sawdust and crop waste) | Biomass | Flue gas | 1 | 1.24 | NA | Estimated from measurements of OC, EC, metals, and ions (but not sulfate) |
| Christian 2010 | $PM_{2.5}$ | g/kg fuel | TFK | Biomass (mainly sawdust and crop waste) | Biomass | Flue gas | 1 | 1.96 | NA | Estimated from measurements of OC, EC, metals, and ions (but not sulfate) |
| Christian 2010 | $CH_4$ | g/kg fuel | TFK | Biomass (mainly sawdust and crop waste) | Biomass | Flue gas | 3 | 1.66 | 0.51 | |
| Christian 2010 | $NH_3$ | g/kg fuel | TFK | Biomass (mainly sawdust and crop waste) | Biomass | Flue gas | 3 | 0.02 | 0.01 | |
| Christian 2010 | $CH_3OH$ | g/kg fuel | TFK | Biomass (mainly sawdust and crop waste) | Biomass | Flue gas | 2 | 0.90 | NA | |
| Christian 2010 | $C_2H_4$ | g/kg fuel | TFK | Biomass (mainly sawdust and crop waste) | Biomass | Flue gas | 3 | 0.32 | 0.05 | |
| Christian 2010 | $C_2H_2$ | g/kg fuel | TFK | Biomass (mainly sawdust and crop waste) | Biomass | Flue gas | 3 | 0.09 | 0.07 | |
| Christian 2010 | $C_3H_6$ | g/kg fuel | TFK | Biomass (mainly sawdust and crop waste) | Biomass | Flue gas | 2 | 0.22 | NA | |

| | | | | | | | | | | |
|---|---|---|---|---|---|---|---|---|---|---|
| Christian 2010 | Hac | g/kg fuel | TFK | Biomass (mainly sawdust and crop waste) | Biomass | Flue gas | 1 | 0.21 | NA | |
| Christian 2010 | HFo | g/kg fuel | TFK | Biomass (mainly sawdust and crop waste) | Biomass | Flue gas | 3 | 0.02 | 0.01 | |
| Christian 2010 | HCHO | g/kg fuel | TFK | Biomass (mainly sawdust and crop waste) | Biomass | Flue gas | 3 | 0.05 | 0.02 | |
| Christian 2010 | NMOC | g/kg fuel | TFK | Biomass (mainly sawdust and crop waste) | Biomass | Flue gas | 3 | 1.3 | 0.92 | |
| Co 2009 | CO | g/brick | TIK | Coal briquettes | Coal | Stack | 1 | 12.3 | NA | |
| Co 2009 | $SO_2$ | g/brick | TIK | Coal briquettes | Coal | Stack | 1 | 5.9 | NA | |
| Co 2009 | PM | g/brick | TIK | Coal briquettes | Coal | Stack | 1 | 1.4 | NA | |
| Co 2009 | CO | g/kg brick | TIK | Coal briquettes | Coal | Stack | 1 | 5.59 | NA | 2.2 ± 0.02 kg per piece |
| Co 2009 | $SO_2$ | g/kg brick | TIK | Coal briquettes | Coal | Stack | 1 | 2.68 | NA | 2.2 ± 0.02 kg per piece |
| Co 2009 | PM | g/kg brick | TIK | Coal briquettes | Coal | Stack | 1 | 0.64 | NA | 2.2 ± 0.02 kg per piece |
| Goetz 2018 | $PM_1$ | g/kg fuel | CK | Coal, sawdust, hardwood | Coal & biomass | Fugitive emissions | 1 | 1.759 | NA | |
| Goetz 2018 | $PM_1$ | g/kg fuel | FDZ | Coal, bagasse | Coal & biomass | Flue gas | 1 | 1.823 | NA | |
| Goetz 2018 | BC | g/kg fuel | CK | Coal, sawdust, hardwood | Coal & biomass | fugitive emissions | 1 | 0.014 | NA | |
| Goetz 2018 | BC | g/kg fuel | FDZ | Coal, bagasse | Coal & biomass | Flue gas | 1 | 0.466 | NA | |
| Goetz 2018 | SO4 | g/kg fuel | CK | Coal, sawdust, hardwood | Coal & biomass | fugitive emissions | 1 | 0.484 | NA | |
| Goetz 2018 | SO4 | g/kg fuel | FDZ | Coal, bagasse | Coal & biomass | Flue gas | 1 | 0.955 | NA | |
| Goetz 2018 | NH4 | g/kg fuel | CK | Coal, sawdust, hardwood | Coal & biomass | fugitive emissions | 1 | 0.168 | NA | |
| Goetz 2018 | NH4 | g/kg fuel | FDZ | Coal, bagasse | Coal & biomass | Flue gas | 1 | 0.108 | NA | |
| Goetz 2018 | Chl | g/kg fuel | CK | Coal, sawdust, hardwood | Coal & biomass | fugitive emissions | 1 | 0.094 | NA | |

| | | | | | | | | | |
|---|---|---|---|---|---|---|---|---|---|
| Haque 2018 | $PM_{2.5}$ | g/kg fuel | BTK | Coal and biomass | Coal & biomass | Stack | 10 | 6.12 | 4.3 |
| Haque 2018 | $PM_{2.5}$ | g/kg fuel | FDZ | Coal | Coal | Stack | 6 | 5.88 | 3.2 |
| Haque 2018 | $PM_{2.5}$ | g/kg fuel | HK | Coal | Coal | Stack | 2 | 4.73 | 2.4 |
| Haque 2018 | $PM_{2.5}$ | g/kg brick | BTK | Coal and biomass | Coal & biomass | Stack | 10 | 0.44 | 0.31 |
| Haque 2018 | $PM_{2.5}$ | g/kg brick | FDZ | Coal | Coal | Stack | 6 | 0.42 | 0.29 |
| Haque 2018 | $PM_{2.5}$ | g/kg brick | HK | Coal | Coal | Stack | 2 | 0.34 | 0.18 |
| Haque 2018 | $PM_{2.5}$ | g/MJ | BTK | Coal and biomass | Coal & biomass | Stack | 10 | 0.27 | 0.21 |
| Haque 2018 | $PM_{2.5}$ | g/MJ | FDZ | Coal | Coal | Stack | 6 | 0.25 | 0.18 |
| Haque 2018 | $PM_{2.5}$ | g/MJ | HK | Coal | Coal | Stack | 2 | 0.21 | 0.11 |
| Haque 2018 | BC | g/kg fuel | BTK | Coal and biomass | Coal & biomass | Stack | 10 | 0.43 | 0.18 |
| Haque 2018 | BC | g/kg fuel | FDZ | Coal | Coal | Stack | 6 | 0.31 | 0.09 |
| Haque 2018 | BC | g/kg fuel | HK | Coal | Coal | Stack | 2 | 0.25 | 0.07 |
| Haque 2018 | BC | g/kg brick | BTK | Coal and biomass | Coal & biomass | Stack | 10 | 0.03 | 0.2 |
| Haque 2018 | BC | g/kg brick | FDZ | Coal | Coal | Stack | 6 | 0.02 | 0.01 |
| Haque 2018 | BC | g/kg brick | HK | Coal | Coal | Stack | 2 | 0.01 | 0.01 |
| Haque 2018 | BC | g/MJ | BTK | Coal and biomass | Coal & biomass | Stack | 10 | 0.02 | 0.01 |
| Haque 2018 | BC | g/MJ | FDZ | Coal | Coal | Stack | 6 | 0.014 | 0.004 |
| Haque 2018 | BC | g/MJ | HK | Coal | Coal | Stack | 2 | 0.011 | 0.003 |
| Haque 2018 | $CO_2$ | g/kg fuel | BTK | Coal and biomass | Coal & biomass | Stack | 10 | 242 | 92 |
| Haque 2018 | $CO_2$ | g/kg fuel | FDZ | Coal | Coal | Stack | 6 | 379 | 144 |
| Haque 2018 | $CO_2$ | g/kg fuel | HK | Coal | Coal | Stack | 2 | 237 | 162 |
| Haque 2018 | $CO_2$ | g/kg brick | BTK | Coal and biomass | Coal & biomass | Stack | 10 | 14.9 | 5.8 |
| Haque 2018 | $CO_2$ | g/kg brick | FDZ | Coal | Coal | Stack | 6 | 21.4 | 8.6 |
| Haque 2018 | $CO_2$ | g/kg brick | HK | Coal | Coal | Stack | 2 | 12.6 | 5.9 |
| Haque 2018 | $CO_2$ | g/MJ | BTK | Coal and biomass | Coal & biomass | Stack | 10 | 10.7 | 4.1 |

| | | | | | | | | | |
|---|---|---|---|---|---|---|---|---|---|
| Haque 2018 | $CO_2$ | g/MJ | FDZ | Coal | Coal | Stack | 6 | 16.9 | 6.4 |
| Haque 2018 | $CO_2$ | g/MJ | HK | Coal | Coal | Stack | 2 | 10.9 | 4.9 |
| Haque 2018 | CO | g/kg fuel | BTK | Coal and biomass | Coal & biomass | Stack | 10 | 7.5 | 6.8 |
| Haque 2018 | CO | g/kg fuel | FDZ | Coal | Coal | Stack | 6 | 6.8 | 5.9 |
| Haque 2018 | CO | g/kg fuel | HK | Coal | Coal | Stack | 2 | 5.6 | 3.1 |
| Haque 2018 | CO | g/kg brick | BTK | Coal and biomass | Coal & biomass | Stack | 10 | 0.44 | 0.41 |
| Haque 2018 | CO | g/kg brick | FDZ | Coal | Coal | Stack | 6 | 0.41 | 0.38 |
| Haque 2018 | CO | g/kg brick | HK | Coal | Coal | Stack | 2 | 0.31 | 0.18 |
| Haque 2018 | CO | g/MJ | BTK | Coal and biomass | Coal & biomass | Stack | 10 | 0.33 | 0.3 |
| Haque 2018 | CO | g/MJ | FDZ | Coal | Coal | Stack | 6 | 0.31 | 0.26 |
| Haque 2018 | CO | g/MJ | HK | Coal | Coal | Stack | 2 | 0.24 | 0.16 |
| Haque 2018 | $SO_2$ | g/kg fuel | BTK | Coal and biomass | Coal & biomass | Stack | 10 | 26.7 | 17.6 |
| Haque 2018 | $SO_2$ | g/kg fuel | FDZ | Coal | Coal | Stack | 6 | 18.5 | 13.4 |
| Haque 2018 | $SO_2$ | g/kg fuel | HK | Coal | Coal | Stack | 2 | 33.7 | 15.3 |
| Haque 2018 | $SO_2$ | g/kg brick | BTK | Coal and biomass | Coal & biomass | Stack | 10 | 1.8 | 1.6 |
| Haque 2018 | $SO_2$ | g/kg brick | FDZ | Coal | Coal | Stack | 6 | 1.1 | 0.87 |
| Haque 2018 | $SO_2$ | g/kg brick | HK | Coal | Coal | Stack | 2 | 1.8 | 0.87 |
| Haque 2018 | $SO_2$ | g/MJ | BTK | Coal and biomass | Coal & biomass | Stack | 10 | 1.2 | 0.8 |
| Haque 2018 | $SO_2$ | g/MJ | FDZ | Coal | Coal | Stack | 6 | 0.8 | 0.6 |
| Haque 2018 | $SO_2$ | g/MJ | HK | Coal | Coal | Stack | 2 | 1.5 | 0.78 |
| Haque 2018 | VOCs | g/kg fuel | BTK | Coal and biomass | Coal & biomass | Stack | 10 | 1085 | 947 |
| Haque 2018 | VOCs | g/kg fuel | FDZ | Coal | Coal | Stack | 6 | 1371 | 526 |
| Haque 2018 | VOCs | g/kg fuel | HK | Coal | Coal | Stack | 2 | 1953 | 972 |
| Haque 2018 | VOCs | g/kg brick | BTK | Coal and biomass | Coal & biomass | Stack | 10 | 76.7 | 47.2 |
| Haque 2018 | VOCs | g/kg brick | FDZ | Coal | Coal | Stack | 6 | 77.6 | 35.9 |
| Haque 2018 | VOCs | g/kg brick | HK | Coal | Coal | Stack | 2 | 98 | 48.3 |

| | | | | | | | | | |
|---|---|---|---|---|---|---|---|---|---|
| Haque 2018 | VOCs | g/MJ | BTK | Coal and biomass | Coal & biomass | Stack | 10 | 48.4 | 41.6 |
| Haque 2018 | VOCs | g/MJ | FDZ | Coal | Coal | Stack | 6 | 61.2 | 23.4 |
| Haque 2018 | VOCs | g/MJ | HK | Coal | Coal | Stack | 2 | 87.4 | 41.8 |
| Hu 2019 | $SO_2$ | g/kg fuel | NS | Coal, clay, coal fly ash | Coal | Flue gas | 34 | 7.17 | 7.49 |
| Hu 2019 | $NO_x$ | g/kg fuel | NS | Coal, clay, coal fly ash | Coal | Flue gas | 34 | 1.09 | 0.9 |
| Hu 2019 | PM | g/kg fuel | NS | Coal, clay, coal fly ash | Coal | Flue gas | 34 | 0.56 | 1.13 |
| Hu 2019 | VOCs | g/kg fuel | NS | Coal, clay, coal fly ash | Coal | Flue gas | 34 | 0.52 | 0.92 |
| Hu 2019 | $SO_2$ | kg/MY output | NS | Coal, clay, coal fly ash | Coal | Flue gas | 34 | 56.5 | 94.1 |
| Hu 2019 | $NO_x$ | kg/MY output | NS | Coal, clay, coal fly ash | Coal | Flue gas | 34 | 2.74 | 2.08 |
| Hu 2019 | PM | kg/MY output | NS | Coal, clay, coal fly ash | Coal | Flue gas | 34 | 0.86 | 0.82 |
| Hu 2019 | VOCs | kg/MY output | NS | Coal, clay, coal fly ash | Coal | Flue gas | 34 | 1.15 | 0.89 |
| Hu 2019 | $SO_2$ | g/kg brick | NS | Coal, clay, coal fly ash | Coal | Flue gas | 34 | 5.48 | 6.25 |
| Hu 2019 | $NO_x$ | g/kg brick | NS | Coal, clay, coal fly ash | Coal | Flue gas | 34 | 0.59 | 0.8 |
| Hu 2019 | PM | g/kg brick | NS | Coal, clay, coal fly ash | Coal | Flue gas | 34 | 0.19 | 0.27 |
| Hu 2019 | VOCs | g/kg brick | NS | Coal, clay, coal fly ash | Coal | Flue gas | 34 | 0.27 | 0.45 |
| Jayarathne 2018 | $PM_{2.5}$ | g/kg fuel | FDZ | Coal, bagasse | Coal & biomass | Flue gas | 1 | 19.06 | NA |
| Jayarathne 2018 | $PM_{2.5}$ | g/kg fuel | FDZ | Coal, bagasse | Coal & biomass | Flue gas | 1 | 11.75 | NA |
| Jayarathne 2018 | $PM_{2.5}$ | g/kg fuel | FDZ | Coal, bagasse | Coal & biomass | Flue gas | 1 | 14.52 | NA |
| Jayarathne 2018 | EC | g/kg fuel | FDZ | Coal, bagasse | Coal & biomass | Flue gas | 1 | <0.01 | NA |
| Jayarathne 2018 | EC | g/kg fuel | FDZ | Coal, bagasse | Coal & biomass | Flue gas | 1 | <0.01 | NA |
| Jayarathne 2018 | EC | g/kg fuel | FDZ | Coal, bagasse | Coal & biomass | Flue gas | 1 | <0.01 | NA |
| Jayarathne 2018 | OC | g/kg fuel | FDZ | Coal, bagasse | Coal & biomass | Flue gas | 1 | 1.12 | NA |

| | | | | | | | | | | |
|---|---|---|---|---|---|---|---|---|---|---|
| Jayarathne 2018 | OC | g/kg fuel | FDZ | Coal, bagasse | Coal & biomass | Flue gas | 1 | 1.26 | NA | |
| Jayarathne 2018 | OC | g/kg fuel | FDZ | Coal, bagasse | Coal & biomass | Flue gas | 1 | 0.63 | NA | |
| Jayarathne 2018 | $PM_{2.5}$ | g/kg fuel | CK | Coal, hardwood | Coal & biomass | Fugitive emissions | 1 | 8.02 | NA | |
| Jayarathne 2018 | $PM_{2.5}$ | g/kg fuel | CK | Coal, hardwood | Coal & biomass | Fugitive emissions | 1 | 10.54 | NA | |
| Jayarathne 2018 | $PM_{2.5}$ | g/kg fuel | CK | Coal, hardwood | Coal & biomass | Fugitive emissions | 1 | 13.41 | NA | |
| Jayarathne 2018 | EC | g/kg fuel | CK | Coal, hardwood | Coal & biomass | Fugitive emissions | 1 | <0.02 | NA | |
| Jayarathne 2018 | EC | g/kg fuel | CK | Coal, hardwood | Coal & biomass | Fugitive emissions | 1 | <0.1 | NA | |
| Jayarathne 2018 | EC | g/kg fuel | CK | Coal, hardwood | Coal & biomass | Fugitive emissions | 1 | <0.1 | NA | |
| Jayarathne 2018 | OC | g/kg fuel | CK | Coal, hardwood | Coal & biomass | Fugitive emissions | 1 | 4.68 | NA | |
| Jayarathne 2018 | OC | g/kg fuel | CK | Coal, hardwood | Coal & biomass | Fugitive emissions | 1 | 7.26 | NA | |
| Jayarathne 2018 | OC | g/kg fuel | CK | Coal, hardwood | Coal & biomass | Fugitive emissions | 1 | 8.37 | NA | |
| Le 2009 | CO | g/brick | TIK | Coal briquettes | Coal | Stack | NA | 6.35 | 0.3 | |
| Le 2009 | $SO_2$ | g/brick | TIK | Coal briquettes | Coal | Stack | NA | 0.52 | 0.02 | |
| Le 2009 | PM | g/brick | TIK | Coal briquettes | Coal | Stack | NA | 0.64 | NA | |
| Le 2009 | CO | g/kg brick | TIK | Coal briquettes | Coal | Stack | NA | 2.886 | 0.136 | 2.2 ± 0.02 kg per piece |
| Le 2009 | $SO_2$ | g/kg brick | TIK | Coal briquettes | Coal | Stack | NA | 0.236 | 0.009 | 2.2 ± 0.02 kg per piece |
| Le 2009 | PM | g/kg brick | TIK | Coal briquettes | Coal | Stack | NA | 0.291 | NA | 2.2 ± 0.02 kg per piece |
| Nasim 2020 | $SO_2$ | g/kg fuel | BTK | Coal | Coal | Stack | 1 | 213 | NA | |
| Nasim 2020 | $SO_2$ | g/kg fuel | FDZ | Coal, rice husk, poultry waste | Coal & biomass | Stack | 1 | 7.97 | NA | |
| Nasim 2020 | CO | g/kg fuel | BTK | Coal | Coal | Stack | 1 | 168 | NA | |
| Nasim 2020 | CO | g/kg fuel | FDZ | Coal, rice husk, poultry waste | Coal & biomass | Stack | 1 | 27.97 | NA | |
| Nasim 2020 | $NO_x$ | g/kg fuel | BTK | Coal | Coal | Stack | 1 | 1.42 | NA | |
| Nasim 2020 | $NO_x$ | g/kg fuel | FDZ | Coal, rice husk, poultry waste | Coal & biomass | Stack | 1 | 1.80 | NA | |

| | | | | | | | | | | |
|---|---|---|---|---|---|---|---|---|---|---|
| Nasim 2020 | $CO_2$ | g/kg fuel | BTK | Coal | Coal | Stack | 1 | 7891 | NA | Non-physical value excluded from meta-analysis |
| Nasim 2020 | $CO_2$ | g/kg fuel | FDZ | Coal, rice husk, poultry waste | Coal & biomass | Stack | 1 | 2836 | NA | |
| Nasim 2020 | PM | g/kg fuel | BTK | Coal | Coal | Stack | 1 | 42.22 | NA | |
| Nasim 2020 | PM | g/kg fuel | FDZ | Coal, rice husk, poultry waste | Coal & biomass | Stack | 1 | 1.01 | NA | |
| Nasim 2020 | $SO_2$ | g/MJ | BTK | Coal | Coal | Stack | 1 | 9.20 | NA | |
| Nasim 2020 | $SO_2$ | g/MJ | FDZ | Coal, rice husk, poultry waste | Coal & biomass | Stack | 1 | 0.298 | NA | |
| Nasim 2020 | CO | g/MJ | BTK | Coal | Coal | Stack | 1 | 7.24 | NA | |
| Nasim 2020 | CO | g/MJ | FDZ | Coal, rice husk, poultry waste | Coal & biomass | Stack | 1 | 1.04 | NA | |
| Nasim 2020 | $NO_x$ | g/MJ | BTK | Coal | Coal | Stack | 1 | 0.061 | NA | |
| Nasim 2020 | $NO_x$ | g/MJ | FDZ | Coal, rice husk, poultry waste | Coal & biomass | Stack | 1 | 0.067 | NA | |
| Nasim 2020 | $CO_2$ | g/MJ | BTK | Coal | Coal | Stack | 1 | 341 | NA | Excluded from meta-analysis |
| Nasim 2020 | $CO_2$ | g/MJ | FDZ | Coal, rice husk, poultry waste | Coal & biomass | Stack | 1 | 106 | NA | |
| Nasim 2020 | PM | g/MJ | BTK | Coal | Coal | Stack | 1 | 1.83 | NA | |
| Nasim 2020 | PM | g/MJ | FDZ | Coal, rice husk, poultry waste | Coal & biomass | Stack | 1 | 0.038 | NA | |
| Nepal 2019 | $CO_2$ | g/kg fuel | BTK | Coal, rice husk, briquette | Coal & biomass | Stack | 4 | 1633 | 134 | |
| Nepal 2019 | $SO_2$ | g/kg fuel | BTK | Coal, rice husk, briquette | Coal & biomass | Stack | 4 | 22 | 22 | |
| Nepal 2019 | $PM_{2.5}$ | g/kg fuel | BTK | Coal, rice husk, briquette | Coal & biomass | Stack | 4 | 3.8 | 2.6 | |
| Nepal 2019 | BC | g/kg fuel | BTK | Coal, rice husk, briquette | Coal & biomass | Stack | 4 | 0.6 | 0.2 | |
| Nepal 2019 | $CO_2$ | g/kg fuel | FDZ | Coal, rice husk, sawdust | Coal & biomass | Stack | 3 | 1981 | 232 | |
| Nepal 2019 | $SO_2$ | g/kg fuel | FDZ | Coal, rice husk, sawdust | Coal & biomass | Stack | 3 | 24 | 22 | |
| Nepal 2019 | $PM_{2.5}$ | g/kg fuel | FDZ | Coal, rice husk, sawdust | Coal & biomass | Stack | 3 | 3.1 | 1 | |
| Nepal 2019 | BC | g/kg fuel | FDZ | Coal, rice husk, sawdust | Coal & biomass | Stack | 3 | 0.4 | 0.2 | |

| | | | | | | | | | | |
|---|---|---|---|---|---|---|---|---|---|---|
| Nepal 2019 | $CO_2$ | g/kg brick | BTK | Coal, rice husk, briquette | Coal & biomass | Stack | 4 | 96 | 16 | |
| Nepal 2019 | $SO_2$ | g/kg brick | BTK | Coal, rice husk, briquette | Coal & biomass | Stack | 4 | 1.2 | 1.2 | |
| Nepal 2019 | $PM_{2.5}$ | g/kg brick | BTK | Coal, rice husk, briquette | Coal & biomass | Stack | 4 | 0.2 | 0.1 | |
| Nepal 2019 | BC | g/kg brick | BTK | Coal, rice husk, briquette | Coal & biomass | Stack | 4 | 0.03 | 0.01 | |
| Nepal 2019 | $CO_2$ | g/kg brick | FDZ | Coal, rice husk, sawdust | Coal & biomass | Stack | 3 | 82 | 19 | |
| Nepal 2019 | $SO_2$ | g/kg brick | FDZ | Coal, rice husk, sawdust | Coal & biomass | Stack | 3 | 0.9 | 0.5 | |
| Nepal 2019 | $PM_{2.5}$ | g/kg brick | FDZ | Coal, rice husk, sawdust | Coal & biomass | Stack | 3 | 0.1 | 0.0 | |
| Nepal 2019 | BC | g/kg brick | FDZ | Coal, rice husk, sawdust | Coal & biomass | Stack | 3 | 0.01 | 0.00 | |
| Ortinez-Alvarez 2018 | $PM_{2.5}$ | g/Mg brick | TFK | Pine wood | Biomass | Fugitive emissions | 1 | 632 | NA | Units in paper incorrectly shown as g/mg brick. Confirmed with authors that the correct units are g/Mg brick. |
| Ortinez-Alvarez 2018 | $PM_{2.5}$ | g/Mg brick | MK2 | Pine wood | Biomass | Flue gas | 1 | 386 | NA | |
| Ortinez-Alvarez 2018 | $PM_{10}$ | g/Mg brick | TFK | Pine wood | Biomass | Fugitive emissions | 1 | 735 | NA | |
| Ortinez-Alvarez 2018 | $PM_{10}$ | g/Mg brick | MK2 | Pine wood | Biomass | Flue gas | 1 | 448 | NA | |
| Ortinez-Alvarez 2018 | EC | g/Mg brick | TFK | Pine wood | Biomass | Fugitive emissions | 1 | 51 | NA | |
| Ortinez-Alvarez 2018 | EC | g/Mg brick | MK2 | Pine wood | Biomass | Flue gas | 1 | 38 | NA | |
| Ortinez-Alvarez 2018 | OC | g/Mg brick | TFK | Pine wood | Biomass | Fugitive emissions | 1 | 345 | NA | |
| Ortinez-Alvarez 2018 | OC | g/Mg brick | MK2 | Pine wood | Biomass | Flue gas | 1 | 287 | NA | |
| Pangtey 2004 | $SO_2$ | ppm | BTK | Coal, firewood | Coal & biomass | BKS | 30 | 0.35 | NA | |
| Pangtey 2004 | $NO_x$ | ppm | BTK | Coal, firewood | Coal & biomass | BKS | 30 | 0.23 | NA | |
| Pangtey 2004 | CO | ppm | BTK | Coal, firewood | Coal & biomass | BKS | 30 | 3.44 | NA | |
| Pangtey 2004 | TSP | ppm | BTK | Coal, firewood | Coal & biomass | BKS | 30 | 93.3 | NA | |
| Pangtey 2004 | RSP | ppm | BTK | Coal, firewood | Coal & biomass | BKS | 30 | 4.66 | NA | |
| Rajarathnam 2014 | PM | g/MJ | BTK | Coal, rubber tires, wood logs | Coal & biomass | Stack | 15 | 0.66 | 0.594 | India |

| | | | | | | | | | | |
|---|---|---|---|---|---|---|---|---|---|---|
| Rajarathnam 2014 | $SO_2$ | g/MJ | BTK | Coal, rubber tires, wood logs | Coal & biomass | Stack | 15 | 0.39 | 0.359 | India |
| Rajarathnam 2014 | CO | g/MJ | BTK | Coal, rubber tires, wood logs | Coal & biomass | Stack | 15 | 2.96 | 2.69 | India |
| Rajarathnam 2014 | $CO_2$ | g/MJ | BTK | Coal, rubber tires, wood logs | Coal & biomass | Stack | 15 | 140 | 79.8 | India |
| Rajarathnam 2014 | PM | g/MJ | NDZ | Coal, biomass, sawdust | Coal & biomass | Stack | 15 | 0.21 | 0.1911 | India |
| Rajarathnam 2014 | $SO_2$ | g/MJ | NDZ | Coal, biomass, sawdust | Coal & biomass | Stack | 15 | 0.06 | 0.0912 | India |
| Rajarathnam 2014 | CO | g/MJ | NDZ | Coal, biomass, sawdust | Coal & biomass | Stack | 15 | 0.32 | 0.3104 | India |
| Rajarathnam 2014 | $CO_2$ | g/MJ | NDZ | Coal, biomass, sawdust | Coal & biomass | Stack | 15 | 113 | 33.9 | India |
| Rajarathnam 2014 | PM | g/MJ | FDZ | Coal | Coal | Stack | 9 | 0.23 | 0.1932 | India |
| Rajarathnam 2014 | $SO_2$ | g/MJ | FDZ | Coal | Coal | Stack | 9 | 0.23 | 0.23 | India |
| Rajarathnam 2014 | CO | g/MJ | FDZ | Coal | Coal | Stack | 9 | 1.96 | 1.4896 | India |
| Rajarathnam 2014 | $CO_2$ | g/MJ | FDZ | Coal | Coal | Stack | 9 | 92 | 74.52 | India |
| Rajarathnam 2014 | PM | g/MJ | VSBK | Coal | Coal | Stack | 4 | 0.10 | 0.018 | India |
| Rajarathnam 2014 | $SO_2$ | g/MJ | VSBK | Coal | Coal | Stack | 4 | 0.11 | 0.0209 | India |
| Rajarathnam 2014 | CO | g/MJ | VSBK | Coal | Coal | Stack | 4 | 4.39 | 1.7121 | India |
| Rajarathnam 2014 | $CO_2$ | g/MJ | VSBK | Coal | Coal | Stack | 4 | 126 | 35.28 | India |
| Rajarathnam 2014 | PM | g/MJ | DDK | Biomass, eucalyptus branches | Biomass | Stack | 3 | 0.54 | 0.486 | India |
| Rajarathnam 2014 | $SO_2$ | g/MJ | DDK | Biomass, eucalyptus branches | Biomass | Stack | 3 | <0.1 | 0.003 | India |
| Rajarathnam 2014 | CO | g/MJ | DDK | Biomass, eucalyptus branches | Biomass | Stack | 3 | 5.17 | 0.2068 | India |
| Rajarathnam 2014 | $CO_2$ | g/MJ | DDK | Biomass, eucalyptus branches | Biomass | Stack | 3 | 181 | 61.54 | India |
| Rajarathnam 2014 | PM | g/MJ | VSBK | Coal | Coal | Stack | 3 | 0.22 | 0.0352 | Vietnam |

| | | | | | | | | | | |
|---|---|---|---|---|---|---|---|---|---|---|
| Rajarathnam 2014 | $SO_2$ | g/MJ | VSBK | Coal | Coal | Stack | 3 | 1.78 | 0.0178 | Vietnam |
| Rajarathnam 2014 | CO | g/MJ | VSBK | Coal | Coal | Stack | 3 | 2.93 | 0.3516 | Vietnam |
| Rajarathnam 2014 | $CO_2$ | g/MJ | VSBK | Coal | Coal | Stack | 3 | 146 | 10.22 | Vietnam |
| Rajarathnam 2014 | PM | g/MJ | TK | Coal | Coal | Stack | 3 | 0.21 | 0.0252 | Vietnam |
| Rajarathnam 2014 | $SO_2$ | g/MJ | TK | Coal | Coal | Stack | 3 | 0.49 | 0.0147 | Vietnam |
| Rajarathnam 2014 | CO | g/MJ | TK | Coal | Coal | Stack | 3 | 1.56 | 0.4056 | Vietnam |
| Rajarathnam 2014 | $CO_2$ | g/MJ | TK | Coal | Coal | Stack | 3 | 109 | 10.9 | Vietnam |
| Rajarathnam 2014 | PM | g/kg brick | BTK | Coal, rubber tires, wood logs | Coal & biomass | Stack | 15 | 0.89 | 0.8633 | India |
| Rajarathnam 2014 | $SO_2$ | g/kg brick | BTK | Coal, rubber tires, wood logs | Coal & biomass | Stack | 15 | 0.52 | 0.5096 | India |
| Rajarathnam 2014 | CO | g/kg brick | BTK | Coal, rubber tires, wood logs | Coal & biomass | Stack | 15 | 3.63 | 2.9766 | India |
| Rajarathnam 2014 | $CO_2$ | g/kg brick | BTK | Coal, rubber tires, wood logs | Coal & biomass | Stack | 15 | 179 | 100.24 | India |
| Rajarathnam 2014 | PM | g/kg brick | NDZ | Coal, biomass, sawdust | Coal & biomass | Stack | 15 | 0.22 | 0.1958 | India |
| Rajarathnam 2014 | $SO_2$ | g/kg brick | NDZ | Coal, biomass, sawdust | Coal & biomass | Stack | 15 | 0.06 | 0.0888 | India |
| Rajarathnam 2014 | CO | g/kg brick | NDZ | Coal, biomass, sawdust | Coal & biomass | Stack | 15 | 0.35 | 0.3675 | India |
| Rajarathnam 2014 | $CO_2$ | g/kg brick | NDZ | Coal, biomass, sawdust | Coal & biomass | Stack | 15 | 119 | 38.08 | India |
| Rajarathnam 2014 | PM | g/kg brick | FDZ | Coal | Coal | Stack | 9 | 0.24 | 0.2016 | India |
| Rajarathnam 2014 | $SO_2$ | g/kg brick | FDZ | Coal | Coal | Stack | 9 | 0.24 | 0.2376 | India |
| Rajarathnam 2014 | CO | g/kg brick | FDZ | Coal | Coal | Stack | 9 | 2.04 | 1.5504 | India |
| Rajarathnam 2014 | $CO_2$ | g/kg brick | FDZ | Coal | Coal | Stack | 9 | 96 | 76.8 | India |
| Rajarathnam 2014 | PM | g/kg brick | VSBK | Coal | Coal | Stack | 4 | 0.09 | 0.0162 | India |
| Rajarathnam 2014 | $SO_2$ | g/kg brick | VSBK | Coal | Coal | Stack | 4 | 0.10 | 0.019 | India |
| Rajarathnam 2014 | CO | g/kg brick | VSBK | Coal | Coal | Stack | 4 | 4.14 | 1.6146 | India |

| | | | | | | | | | | |
|---|---|---|---|---|---|---|---|---|---|---|
| Rajarathnam 2014 | $CO_2$ | g/kg brick | VSBK | Coal | Coal | Stack | 4 | 118 | 33.04 | India |
| Rajarathnam 2014 | PM | g/kg brick | DDK | Biomass, eucalyptus branches | Biomass | Stack | 3 | 1.56 | 1.404 | India |
| Rajarathnam 2014 | $SO_2$ | g/kg brick | DDK | Biomass, eucalyptus branches | Biomass | Stack | 3 | 0.00 | 0 | India |
| Rajarathnam 2014 | CO | g/kg brick | DDK | Biomass, eucalyptus branches | Biomass | Stack | 3 | 5.01 | 0.2004 | India |
| Rajarathnam 2014 | $CO_2$ | g/kg brick | DDK | Biomass, eucalyptus branches | Biomass | Stack | 3 | 526 | 178.84 | India |
| Rajarathnam 2014 | PM | g/kg brick | VSBK | Coal | Coal | Stack | 3 | 0.12 | 0.0192 | India |
| Rajarathnam 2014 | $SO_2$ | g/kg brick | VSBK | Coal | Coal | Stack | 3 | 0.97 | 0.0097 | Vietnam |
| Rajarathnam 2014 | CO | g/kg brick | VSBK | Coal | Coal | Stack | 3 | 1.59 | 0.1908 | Vietnam |
| Rajarathnam 2014 | $CO_2$ | g/kg brick | VSBK | Coal | Coal | Stack | 3 | 79 | 5.53 | Vietnam |
| Rajarathnam 2014 | PM | g/kg brick | TK | Coal | Coal | Stack | 3 | 0.31 | 0.0372 | Vietnam |
| Rajarathnam 2014 | $SO_2$ | g/kg brick | TK | Coal | Coal | Stack | 3 | 0.72 | 0.0288 | Vietnam |
| Rajarathnam 2014 | CO | g/kg brick | TK | Coal | Coal | Stack | 3 | 2.28 | 0.5928 | Vietnam |
| Rajarathnam 2014 | $CO_2$ | g/kg brick | TK | Coal | Coal | Stack | 3 | 149 | 14.9 | Vietnam |
| Rauf 2022 | CO | g/kg brick | BTK | Coal, sawdust, wood chips, waste tires | Coal & biomass | Stack | 1 | 2 | NA | |
| Rauf 2022 | $PM_{2.5}$ | g/kg brick | BTK | Coal, sawdust, wood chips, waste tires | Coal & biomass | Stack | 1 | 1.06 | NA | |
| Rauf 2022 | BC | g/kg brick | BTK | Coal, sawdust, wood chips, waste tires | Coal & biomass | Stack | 1 | 0.043 | NA | |
| Rauf 2022 | $CO_2$ | g/kg brick | BTK | Coal, sawdust, wood chips, waste tires | Coal & biomass | Stack | 1 | 131 | NA | |

| | | | | | | | | | |
|---|---|---|---|---|---|---|---|---|---|
| Rauf 2022 | $SO_2$ | g/kg brick | BTK | Coal, sawdust, wood chips, waste tires | Coal & biomass | Stack | 1 | 1.9 | NA |
| Rauf 2022 | CO | g/kg brick | FDZ | Coal, sawdust | Coal & biomass | Stack | 1 | 1.62 | NA |
| Rauf 2022 | $PM_{2.5}$ | g/kg brick | FDZ | Coal, sawdust | Coal & biomass | Stack | 1 | 0.24 | NA |
| Rauf 2022 | BC | g/kg brick | FDZ | Coal, sawdust | Coal & biomass | Stack | 1 | 0.02 | NA |
| Rauf 2022 | $CO_2$ | g/kg brick | FDZ | Coal, sawdust | Coal & biomass | Stack | 1 | 101.6 | NA |
| Rauf 2022 | $SO_2$ | g/kg brick | FDZ | Coal, sawdust | Coal & biomass | Stack | 1 | 0.41 | NA |
| Rauf 2022 | CO | g/kg brick | HK | Coal | Coal | Stack | 1 | 1.35 | NA |
| Rauf 2022 | $PM_{2.5}$ | g/kg brick | HK | Coal | Coal | Stack | 1 | 0.19 | NA |
| Rauf 2022 | BC | g/kg brick | HK | Coal | Coal | Stack | 1 | 0.04 | NA |
| Rauf 2022 | $CO_2$ | g/kg brick | HK | Coal | Coal | Stack | 1 | 93.4 | NA |
| Rauf 2022 | $SO_2$ | g/kg brick | HK | Coal | Coal | Stack | 1 | 0.79 | NA |
| Stockwell 2016 | $CO_2$ | g/kg fuel | CK | Coal, hardwood | Coal & biomass | Fugitive emissions | 1 | 2102 | NA |
| Stockwell 2016 | CO | g/kg fuel | CK | Coal, hardwood | Coal & biomass | Fugitive emissions | 1 | 70.9 | NA |
| Stockwell 2016 | $SO_2$ | g/kg fuel | CK | Coal, hardwood | Coal & biomass | Fugitive emissions | 1 | 13.0 | NA |
| Stockwell 2016 | NO | g/kg fuel | CK | Coal, hardwood | Coal & biomass | Fugitive emissions | 1 | BDL | NA |
| Stockwell 2016 | $NO_2$ | g/kg fuel | CK | Coal, hardwood | Coal & biomass | Fugitive emissions | 1 | 0.297 | NA |
| Stockwell 2016 | BC | g/kg fuel | CK | Coal, hardwood | Coal & biomass | Fugitive emissions | 1 | 0.0172 | 0.0075 |
| Stockwell 2016 | CH4 | g/kg fuel | CK | Coal, hardwood | Coal & biomass | Fugitive emissions | 1 | 19.5 | NA |
| Stockwell 2016 | NH3 | g/kg fuel | CK | Coal, hardwood | Coal & biomass | Fugitive emissions | 1 | 0.317 | NA |
| Stockwell 2016 | $CO_2$ | g/kg fuel | FDZ | Coal, bagasse | Coal & biomass | Flue gas | 1 | 2620 | NA |
| Stockwell 2016 | CO | g/kg fuel | FDZ | Coal, bagasse | Coal & biomass | Flue gas | 1 | 10.1 | NA |
| Stockwell 2016 | $SO_2$ | g/kg fuel | FDZ | Coal, bagasse | Coal & biomass | Flue gas | 1 | 12.7 | NA |

| | | | | | | | | | | |
|---|---|---|---|---|---|---|---|---|---|---|
| Stockwell 2016 | NO | g/kg fuel | FDZ | Coal, bagasse | Coal & biomass | Flue gas | 1 | 1.28 | NA | |
| Stockwell 2016 | $NO_2$ | g/kg fuel | FDZ | Coal, bagasse | Coal & biomass | Flue gas | 1 | 0.0821 | NA | |
| Stockwell 2016 | BC | g/kg fuel | FDZ | Coal, bagasse | Coal & biomass | Flue gas | 1 | 0.112 | 0.063 | |
| Stockwell 2016 | CH4 | g/kg fuel | FDZ | Coal, bagasse | Coal & biomass | Flue gas | 1 | 0.087 | NA | |
| Stockwell 2016 | NH3 | g/kg fuel | FDZ | Coal, bagasse | Coal & biomass | Flue gas | 1 | BDL | NA | |
| Stockwell 2016 | $CO_2$ | g/kg fuel | FDZ | Coal, bagasse | Coal & biomass | Stoke holes | 1 | 2234 | NA | |
| Stockwell 2016 | CO | g/kg fuel | FDZ | Coal, bagasse | Coal & biomass | Stoke holes | 1 | 230 | NA | |
| Stockwell 2016 | $SO_2$ | g/kg fuel | FDZ | Coal, bagasse | Coal & biomass | Stoke holes | 1 | 28.5 | NA | |
| Stockwell 2016 | NO | g/kg fuel | FDZ | Coal, bagasse | Coal & biomass | Stoke holes | 1 | 10.4 | NA | |
| Stockwell 2016 | $NO_2$ | g/kg fuel | FDZ | Coal, bagasse | Coal & biomass | Stoke holes | 1 | 1.36 | NA | |
| Stockwell 2016 | CH4 | g/kg fuel | FDZ | Coal, bagasse | Coal & biomass | Stoke holes | 1 | 4.59 | NA | |
| Stockwell 2016 | NH3 | g/kg fuel | FDZ | Coal, bagasse | Coal & biomass | Stoke holes | 1 | BDL | NA | |
| Weyant 2014 | CO | g/kg fuel | BTK | Coal and wood | Coal & biomass | Stack | 3 | 36.5 | 2.3 | |
| Weyant 2014 | CO | g/kg fuel | BTK | Coal and wood | Coal & biomass | Stack | 3 | 53.8 | 5.2 | |
| Weyant 2014 | CO | g/kg fuel | BTK | Coal and wood | Coal & biomass | Stack | 3 | 26.4 | 1.8 | |
| Weyant 2014 | CO | g/kg fuel | NDZ | Coal | Coal | Stack | 3 | 15.0 | 1.0 | |
| Weyant 2014 | CO | g/kg fuel | NDZ | Coal | Coal | Stack | 3 | 6.9 | 1.3 | |
| Weyant 2014 | CO | g/kg fuel | NDZ | Coal | Coal | Stack | 3 | 10.7 | 2.0 | |
| Weyant 2014 | CO | g/kg fuel | FDZ | Coal | Coal | Stack | 3 | 19.7 | 4.8 | |
| Weyant 2014 | CO | g/kg fuel | FDZ | Coal | Coal | Stack | 3 | 32.5 | 4.7 | |
| Weyant 2014 | CO | g/kg fuel | FDZ | Coal | Coal | Stack | 3 | 21.7 | 1.7 | |
| Weyant 2014 | CO | g/kg fuel | VSBK | Coal | Coal | Stack | 3 | 67.4 | 6.7 | India |
| Weyant 2014 | CO | g/kg fuel | VSBK | Coal | Coal | Stack | 3 | 21.6 | 2.7 | Vietnam |
| Weyant 2014 | CO | g/kg fuel | DDK | Wood | Biomass | Stack | 1 | 78.6 | NA | |

| | | | | | | | | | | |
|---|---|---|---|---|---|---|---|---|---|---|
| Weyant 2014 | CO | g/kg fuel | TK | Coal | Coal | Stack | 3 | 29.6 | 4.1 | |
| Weyant 2014 | $PM_{2.5}$ | g/kg fuel | BTK | Coal and wood | Coal & biomass | Stack | 3 | 4.4 | 0.5 | |
| Weyant 2014 | $PM_{2.5}$ | g/kg fuel | BTK | Coal and wood | Coal & biomass | Stack | 3 | 3.7 | 2.1 | |
| Weyant 2014 | $PM_{2.5}$ | g/kg fuel | BTK | Coal and wood | Coal & biomass | Stack | 3 | 1.7 | 2.4 | |
| Weyant 2014 | $PM_{2.5}$ | g/kg fuel | NDZ | Coal | Coal | Stack | 3 | 2.7 | 1.1 | |
| Weyant 2014 | $PM_{2.5}$ | g/kg fuel | NDZ | Coal | Coal | Stack | 3 | 0.5 | 0.2 | |
| Weyant 2014 | $PM_{2.5}$ | g/kg fuel | NDZ | Coal | Coal | Stack | 3 | 3.8 | 0.7 | |
| Weyant 2014 | $PM_{2.5}$ | g/kg fuel | FDZ | Coal | Coal | Stack | 3 | 1.2 | 0.6 | |
| Weyant 2014 | $PM_{2.5}$ | g/kg fuel | FDZ | Coal | Coal | Stack | 3 | 1.0 | 0.2 | |
| Weyant 2014 | $PM_{2.5}$ | g/kg fuel | FDZ | Coal | Coal | Stack | 3 | 0.6 | 0.3 | |
| Weyant 2014 | $PM_{2.5}$ | g/kg fuel | VSBK | Coal | Coal | Stack | 3 | 1.3 | 0.8 | India |
| Weyant 2014 | $PM_{2.5}$ | g/kg fuel | VSBK | Coal | Coal | Stack | 3 | 1.3 | 0.7 | Vietnam |
| Weyant 2014 | $PM_{2.5}$ | g/kg fuel | DDK | Wood | Biomass | Stack | 3 | 3.0 | 0.5 | |
| Weyant 2014 | $PM_{2.5}$ | g/kg fuel | TK | Coal | Coal | Stack | 3 | 1.6 | 1.8 | |
| Weyant 2014 | EC | g/kg fuel | BTK | Coal and wood | Coal & biomass | Stack | 3 | 3.7 | 1.0 | |
| Weyant 2014 | EC | g/kg fuel | BTK | Coal and wood | Coal & biomass | Stack | 3 | 2.7 | 1.3 | |
| Weyant 2014 | EC | g/kg fuel | BTK | Coal and wood | Coal & biomass | Stack | 3 | 1.8 | 0.6 | |
| Weyant 2014 | EC | g/kg fuel | NDZ | Coal | Coal | Stack | 3 | 0.4 | 0.4 | |
| Weyant 2014 | EC | g/kg fuel | NDZ | Coal | Coal | Stack | 3 | 0.2 | 0.2 | |
| Weyant 2014 | EC | g/kg fuel | NDZ | Coal | Coal | Stack | 3 | 0.2 | 0.03 | |
| Weyant 2014 | EC | g/kg fuel | FDZ | Coal | Coal | Stack | 3 | 0.5 | 0.2 | |
| Weyant 2014 | EC | g/kg fuel | FDZ | Coal | Coal | Stack | 3 | 0.1 | 0.05 | |
| Weyant 2014 | EC | g/kg fuel | FDZ | Coal | Coal | Stack | 3 | 0.07 | 0.07 | |
| Weyant 2014 | EC | g/kg fuel | VSBK | Coal | Coal | Stack | 3 | 0.06 | 0.04 | India |
| Weyant 2014 | EC | g/kg fuel | VSBK | Coal | Coal | Stack | 3 | 0.01 | 0.006 | Vietnam |
| Weyant 2014 | EC | g/kg fuel | DDK | Wood | Biomass | Stack | 3 | 1.1 | 0.4 | |

| | | | | | | | | | | |
|---|---|---|---|---|---|---|---|---|---|---|
| Weyant 2014 | EC | g/kg fuel | TK | Coal | Coal | Stack | 3 | 0.01 | 0.01 | |
| Weyant 2014 | CO | g/kg brick | BTK | Coal and wood | Coal & biomass | Stack | 3 | 2.7 | 0.2 | |
| Weyant 2014 | CO | g/kg brick | BTK | Coal and wood | Coal & biomass | Stack | 3 | 2.1 | 0.2 | |
| Weyant 2014 | CO | g/kg brick | BTK | Coal and wood | Coal & biomass | Stack | 3 | 1.3 | 0.1 | |
| Weyant 2014 | CO | g/kg brick | NDZ | Coal | Coal | Stack | 3 | 0.9 | 0.1 | |
| Weyant 2014 | CO | g/kg brick | NDZ | Coal | Coal | Stack | 3 | 0.4 | 0.1 | |
| Weyant 2014 | CO | g/kg brick | NDZ | Coal | Coal | Stack | 3 | 0.5 | 0.1 | |
| Weyant 2014 | CO | g/kg brick | FDZ | Coal | Coal | Stack | 3 | 0.8 | 0.2 | |
| Weyant 2014 | CO | g/kg brick | FDZ | Coal | Coal | Stack | 3 | 1.8 | 0.3 | |
| Weyant 2014 | CO | g/kg brick | FDZ | Coal | Coal | Stack | 3 | 1.1 | 0.1 | |
| Weyant 2014 | CO | g/kg brick | VSBK | Coal | Coal | Stack | 3 | 2.8 | 0.3 | India |
| Weyant 2014 | CO | g/kg brick | VSBK | Coal | Coal | Stack | 3 | 1.6 | 0.2 | Vietnam |
| Weyant 2014 | CO | g/kg brick | DDK | Wood | Biomass | Stack | 1 | 13.2 | NA | |
| Weyant 2014 | CO | g/kg brick | TK | Coal | Coal | Stack | 3 | 4.5 | 0.6 | |
| Weyant 2014 | $PM_{2.5}$ | g/kg brick | BTK | Coal and wood | Coal & biomass | Stack | 3 | 0.33 | 0.04 | |
| Weyant 2014 | $PM_{2.5}$ | g/kg brick | BTK | Coal and wood | Coal & biomass | Stack | 3 | 0.14 | 0.08 | |
| Weyant 2014 | $PM_{2.5}$ | g/kg brick | BTK | Coal and wood | Coal & biomass | Stack | 3 | 0.08 | 0.12 | |
| Weyant 2014 | $PM_{2.5}$ | g/kg brick | NDZ | Coal | Coal | Stack | 3 | 0.16 | 0.07 | |
| Weyant 2014 | $PM_{2.5}$ | g/kg brick | NDZ | Coal | Coal | Stack | 3 | 0.03 | 0.01 | |
| Weyant 2014 | $PM_{2.5}$ | g/kg brick | NDZ | Coal | Coal | Stack | 3 | 0.19 | 0.03 | |
| Weyant 2014 | $PM_{2.5}$ | g/kg brick | FDZ | Coal | Coal | Stack | 3 | 0.05 | 0.02 | |
| Weyant 2014 | $PM_{2.5}$ | g/kg brick | FDZ | Coal | Coal | Stack | 3 | 0.06 | 0.01 | |
| Weyant 2014 | $PM_{2.5}$ | g/kg brick | FDZ | Coal | Coal | Stack | 3 | 0.03 | 0.02 | |
| Weyant 2014 | $PM_{2.5}$ | g/kg brick | VSBK | Coal | Coal | Stack | 3 | 0.05 | 0.03 | India |
| Weyant 2014 | $PM_{2.5}$ | g/kg brick | VSBK | Coal | Coal | Stack | 3 | 0.09 | 0.05 | Vietnam |
| Weyant 2014 | $PM_{2.5}$ | g/kg brick | DDK | Wood | Biomass | Stack | 3 | 0.50 | 0.08 | |

| | | | | | | | | | | |
|---|---|---|---|---|---|---|---|---|---|---|
| Weyant 2014 | $PM_{2.5}$ | g/kg brick | TK | Coal | Coal | Stack | 3 | 0.24 | 0.27 | |
| Weyant 2014 | EC | g/kg brick | BTK | Coal and wood | Coal & biomass | Stack | 3 | 0.27 | 0.07 | |
| Weyant 2014 | EC | g/kg brick | BTK | Coal and wood | Coal & biomass | Stack | 3 | 0.11 | 0.05 | |
| Weyant 2014 | EC | g/kg brick | BTK | Coal and wood | Coal & biomass | Stack | 3 | 0.09 | 0.03 | |
| Weyant 2014 | EC | g/kg brick | NDZ | Coal | Coal | Stack | 3 | 0.03 | 0.02 | |
| Weyant 2014 | EC | g/kg brick | NDZ | Coal | Coal | Stack | 3 | 0.01 | 0.01 | |
| Weyant 2014 | EC | g/kg brick | NDZ | Coal | Coal | Stack | 3 | 0.01 | 0.002 | |
| Weyant 2014 | EC | g/kg brick | FDZ | Coal | Coal | Stack | 3 | 0.02 | 0.01 | |
| Weyant 2014 | EC | g/kg brick | FDZ | Coal | Coal | Stack | 3 | 0.01 | 0.003 | |
| Weyant 2014 | EC | g/kg brick | FDZ | Coal | Coal | Stack | 3 | 0.004 | 0.004 | |
| Weyant 2014 | EC | g/kg brick | VSBK | Coal | Coal | Stack | 3 | 0.002 | 0.002 | India |
| Weyant 2014 | EC | g/kg brick | VSBK | Coal | Coal | Stack | 3 | 0.001 | 0.0004 | Vietnam |
| Weyant 2014 | EC | g/kg brick | DDK | Wood | Biomass | Stack | 3 | 0.19 | 0.07 | |
| Weyant 2014 | EC | g/kg brick | TK | Coal | Coal | Stack | 3 | 0.001 | 0.001 | |
| Ying 2021 | PM | g/brick | TK | Coal, sewage sludge | Coal & sewage sludge | Flue gas | 8 | 0.479 | NA | Cleaning treatment system |
| Ying 2021 | HCl | g/brick | TK | Coal, sewage sludge | Coal & sewage sludge | Flue gas | 8 | 0.607 | NA | Cleaning treatment system |
| Ying 2021 | $SO_2$ | g/brick | TK | Coal, sewage sludge | Coal & sewage sludge | Flue gas | 8 | 0.576 | NA | Cleaning treatment system |
| Ying 2021 | $NO_x$ | g/brick | TK | Coal, sewage sludge | Coal & sewage sludge | Flue gas | 8 | 1.170 | NA | Cleaning treatment system |
| Ying 2021 | $H_2S$ | g/brick | TK | Coal, sewage sludge | Coal & sewage sludge | Flue gas | 8 | 0.000 | NA | Cleaning treatment system |
| Ying 2021 | $NH_3$ | g/brick | TK | Coal, sewage sludge | Coal & sewage sludge | Flue gas | 8 | 0.208 | NA | Cleaning treatment system |
| Zavala 2018 | $PM_{2.5}$ | g/kg fuel | MK2 | Wood | Biomass | Flue gas | NA | 1.94 | 0.6 | SP technique |
| Zavala 2018 | BC | g/kg fuel | MK2 | Wood | Biomass | Flue gas | NA | 0.15 | 0.1 | SP technique |

| | | | | | | | | | | |
|---|---|---|---|---|---|---|---|---|---|---|
| Zavala 2018 | OC | g/kg fuel | MK2 | Wood | Biomass | Flue gas | NA | 0.03 | 0.03 | SP technique |
| Zavala 2018 | $SO_2$ | g/kg fuel | MK2 | Wood | Biomass | Flue gas | NA | 1.0 | 1.4 | SP technique |
| Zavala 2018 | CO | g/kg fuel | MK2 | Wood | Biomass | Flue gas | NA | 44.4 | 17.7 | SP technique |
| Zavala 2018 | $CO_2$ | g/kg fuel | MK2 | Wood | Biomass | Flue gas | NA | 1583 | 28 | SP technique |
| Zavala 2018 | NO | g/kg fuel | MK2 | Wood | Biomass | Flue gas | NA | 1.02 | 0.9 | SP technique |
| Zavala 2018 | $NO_2$ | g/kg fuel | MK2 | Wood | Biomass | Flue gas | NA | 1.7 | 1.8 | SP technique |
| Zavala 2018 | $PM_{2.5}$ | g/kg fuel | TCK | Wood | Biomass | Fugitive emissions | NA | 4.62 | 4.3 | SP technique |
| Zavala 2018 | BC | g/kg fuel | TCK | Wood | Biomass | Fugitive emissions | NA | 0.28 | 0.2 | SP technique |
| Zavala 2018 | OC | g/kg fuel | TCK | Wood | Biomass | Fugitive emissions | NA | 0.3 | 0.7 | SP technique |
| Zavala 2018 | CO | g/kg fuel | TCK | Wood | Biomass | Fugitive emissions | NA | 50.5 | 16.7 | SP technique |
| Zavala 2018 | $CO_2$ | g/kg fuel | TCK | Wood | Biomass | Fugitive emissions | NA | 1527 | 28 | SP technique |
| Zavala 2018 | $PM_{2.5}$ | g/kg fuel | TFK | Wood, diesel, sawdust | Biomass | Fugitive emissions | NA | 1.32 | 1.3 | SP technique |
| Zavala 2018 | BC | g/kg fuel | TFK | Wood, diesel, sawdust | Biomass | Fugitive emissions | NA | 0.54 | 0.8 | SP technique |
| Zavala 2018 | OC | g/kg fuel | TFK | Wood, diesel, sawdust | Biomass | Fugitive emissions | NA | 0.14 | 0.1 | SP technique |
| Zavala 2018 | $SO_2$ | g/kg fuel | TFK | Wood, diesel, sawdust | Biomass | Fugitive emissions | NA | 0.13 | 0.1 | SP technique |
| Zavala 2018 | CO | g/kg fuel | TFK | Wood, diesel, sawdust | Biomass | Fugitive emissions | NA | 105.2 | 24.3 | SP technique |
| Zavala 2018 | $CO_2$ | g/kg fuel | TFK | Wood, diesel, sawdust | Biomass | Fugitive emissions | NA | 1668 | 40 | SP technique |
| Zavala 2018 | NO | g/kg fuel | TFK | Wood, diesel, sawdust | Biomass | Fugitive emissions | NA | 0.76 | 0.3 | SP technique |
| Zavala 2018 | $NO_2$ | g/kg fuel | TFK | Wood, diesel, sawdust | Biomass | Fugitive emissions | NA | 1.01 | 0.6 | SP technique |
| Zavala 2018 | NO | g/kg fuel | MK2 | Wood | Biomass | Flue gas | NA | 1.02 | 0.9 | TR technique |
| Zavala 2018 | $NO_2$ | g/kg fuel | MK2 | Wood | Biomass | Flue gas | NA | 1.7 | 1.8 | TR technique |
| Zavala 2018 | $SO_2$ | g/kg fuel | MK2 | Wood | Biomass | Flue gas | NA | 1.0 | 1.4 | TR technique |
| Zavala 2018 | $PM_{2.5}$ | g/kg fuel | MK2 | Wood | Biomass | Flue gas | NA | 1.66 | 0.8 | TR technique |
| Zavala 2018 | BC | g/kg fuel | MK2 | Wood | Biomass | Flue gas | NA | 0.67 | 0.5 | TR technique |

| | | | | | | | | | | |
|---|---|---|---|---|---|---|---|---|---|---|
| Zavala 2018 | OC | g/kg fuel | MK2 | Wood | Biomass | Flue gas | NA | 0.52 | 0.6 | TR technique |
| Zavala 2018 | CO | g/kg fuel | MK2 | Wood | Biomass | Flue gas | NA | 65.4 | 526 | TR technique |
| Zavala 2018 | $CO_2$ | g/kg fuel | MK2 | Wood | Biomass | Flue gas | NA | 1595 | 58 | TR technique |
| Zavala 2018 | Ethane | g/kg fuel | MK2 | Wood | Biomass | Flue gas | NA | 0.15 | 0.2 | TR technique |
| Zavala 2018 | $CH_3OH$ | g/kg fuel | MK2 | Wood | Biomass | Flue gas | NA | 1.99 | 2.0 | TR technique |
| Zavala 2018 | Acetonitrile | g/kg fuel | MK2 | Wood | Biomass | Flue gas | NA | 0.24 | 0.2 | TR technique |
| Zavala 2018 | Acetaldehyde | g/kg fuel | MK2 | Wood | Biomass | Flue gas | NA | 1.13 | 1.2 | TR technique |
| Zavala 2018 | Acetone | g/kg fuel | MK2 | Wood | Biomass | Flue gas | NA | 1.28 | 1.5 | TR technique |
| Zavala 2018 | HAc | g/kg fuel | MK2 | Wood | Biomass | Flue gas | NA | 2.64 | 3.1 | TR technique |
| Zavala 2018 | Benzene | g/kg fuel | MK2 | Wood | Biomass | Flue gas | NA | 0.84 | 0.9 | TR technique |
| Zavala 2018 | Toluene | g/kg fuel | MK2 | Wood | Biomass | Flue gas | NA | 0.93 | 0.8 | TR technique |
| Zavala 2018 | C2Benzenes | g/kg fuel | MK2 | Wood | Biomass | Flue gas | NA | 1.01 | 1.1 | TR technique |
| Zavala 2018 | C3Benzenes | g/kg fuel | MK2 | Wood | Biomass | Flue gas | NA | 0.86 | 1.0 | TR technique |
| Zavala 2018 | NO | g/kg fuel | TCK | Wood | Biomass | Fugitive emissions | NA | 1.05 | 2.1 | TR technique |
| Zavala 2018 | $NO_2$ | g/kg fuel | TCK | Wood | Biomass | Fugitive emissions | NA | 0.93 | 1.4 | TR technique |
| Zavala 2018 | $SO_2$ | g/kg fuel | TCK | Wood | Biomass | Fugitive emissions | NA | 0.27 | 0.3 | TR technique |
| Zavala 2018 | $PM_{2.5}$ | g/kg fuel | TCK | Wood | Biomass | Fugitive emissions | NA | 2.28 | 1.8 | TR technique |
| Zavala 2018 | BC | g/kg fuel | TCK | Wood | Biomass | Fugitive emissions | NA | 0.73 | 0.6 | TR technique |
| Zavala 2018 | OC | g/kg fuel | TCK | Wood | Biomass | Fugitive emissions | NA | 1.18 | 1.7 | TR technique |
| Zavala 2018 | $CO_2$ | g/kg fuel | TCK | Wood | Biomass | Fugitive emissions | NA | 1597 | 54 | TR technique |
| Zavala 2018 | CO | g/kg fuel | TCK | Wood | Biomass | Fugitive emissions | NA | 65.3 | 43 | TR technique |
| Zavala 2018 | Ethane | g/kg fuel | TCK | Wood | Biomass | Fugitive emissions | NA | 0.21 | 0.22 | TR technique |
| Zavala 2018 | $CH_3OH$ | g/kg fuel | TCK | Wood | Biomass | Fugitive emissions | NA | 1.19 | 2.3 | TR technique |
| Zavala 2018 | Acetonitrile | g/kg fuel | TCK | Wood | Biomass | Fugitive emissions | NA | 0.15 | 0.1 | TR technique |

| | | | | | | | | | | |
|---|---|---|---|---|---|---|---|---|---|---|
| Zavala 2018 | Acetaldehyde | g/kg fuel | TCK | Wood | Biomass | Fugitive emissions | NA | 0.54 | 0.4 | TR technique |
| Zavala 2018 | Acetone | g/kg fuel | TCK | Wood | Biomass | Fugitive emissions | NA | 0.61 | 1.9 | TR technique |
| Zavala 2018 | HAc | g/kg fuel | TCK | Wood | Biomass | Fugitive emissions | NA | 0.89 | 2.6 | TR technique |
| Zavala 2018 | Benzene | g/kg fuel | TCK | Wood | Biomass | Fugitive emissions | NA | 0.66 | 0.7 | TR technique |
| Zavala 2018 | Toluene | g/kg fuel | TCK | Wood | Biomass | Fugitive emissions | NA | 0.42 | 0.9 | TR technique |
| Zavala 2018 | C2Benzenes | g/kg fuel | TCK | Wood | Biomass | Fugitive emissions | NA | 0.54 | 1.5 | TR technique |
| Zavala 2018 | C3Benzenes | g/kg fuel | TCK | Wood | Biomass | Fugitive emissions | NA | 0.45 | 1.2 | TR technique |
| Zavala 2018 | NO | g/kg fuel | TFK | Wood, diesel, sawdust | Biomass | Fugitive emissions | NA | 0.76 | 0.3 | TR technique |
| Zavala 2018 | $NO_2$ | g/kg fuel | TFK | Wood, diesel, sawdust | Biomass | Fugitive emissions | NA | 1.01 | 0.6 | TR technique |
| Zavala 2018 | $SO_2$ | g/kg fuel | TFK | Wood, diesel, sawdust | Biomass | Fugitive emissions | NA | 0.13 | 0.1 | TR technique |
| Zavala 2018 | $PM_{2.5}$ | g/kg fuel | TFK | Wood, diesel, sawdust | Biomass | Fugitive emissions | NA | 1.26 | 2.2 | TR technique |
| Zavala 2018 | BC | g/kg fuel | TFK | Wood, diesel, sawdust | Biomass | Fugitive emissions | NA | 1.03 | 2.2 | TR technique |
| Zavala 2018 | OC | g/kg fuel | TFK | Wood, diesel, sawdust | Biomass | Fugitive emissions | NA | 0.18 | 0.2 | TR technique |
| Zavala 2018 | CO | g/kg fuel | TFK | Wood, diesel, sawdust | Biomass | Fugitive emissions | NA | 105.3 | 36 | TR technique |
| Zavala 2018 | $CO_2$ | g/kg fuel | TFK | Wood, diesel, sawdust | Biomass | Fugitive emissions | NA | 1658 | 43 | TR technique |
| Zavala 2018 | Ethane | g/kg fuel | TFK | Wood, diesel, sawdust | Biomass | Fugitive emissions | NA | 0.44 | 0.1 | TR technique |
| Zavala 2018 | $CH_3OH$ | g/kg fuel | TFK | Wood, diesel, sawdust | Biomass | Fugitive emissions | NA | 3.25 | 1.2 | TR technique |
| Zavala 2018 | Acetonitrile | g/kg fuel | TFK | Wood, diesel, sawdust | Biomass | Fugitive emissions | NA | 0.46 | 0.2 | TR technique |
| Zavala 2018 | Acetaldehyde | g/kg fuel | TFK | Wood, diesel, sawdust | Biomass | Fugitive emissions | NA | 2.18 | 0.5 | TR technique |
| Zavala 2018 | Acetone | g/kg fuel | TFK | Wood, diesel, sawdust | Biomass | Fugitive emissions | NA | 0.91 | 0.3 | TR technique |
| Zavala 2018 | HAc | g/kg fuel | TFK | Wood, diesel, sawdust | Biomass | Fugitive emissions | NA | 1.04 | 0.8 | TR technique |
| Zavala 2018 | Benzene | g/kg fuel | TFK | Wood, diesel, sawdust | Biomass | Fugitive emissions | NA | 0.5 | 0.3 | TR technique |

| | | | | | | | | | | |
|---|---|---|---|---|---|---|---|---|---|---|
| Zavala 2018 | Toluene | g/kg fuel | TFK | Wood, diesel, sawdust | Biomass | Fugitive emissions | NA | 0.28 | 0.2 | TR technique |
| Zavala 2018 | C2Benzenes | g/kg fuel | TFK | Wood, diesel, sawdust | Biomass | Fugitive emissions | NA | 0.19 | 0.1 | TR technique |
| Zavala 2018 | C3Benzenes | g/kg fuel | TFK | Wood, diesel, sawdust | Biomass | Fugitive emissions | NA | 0.13 | 0.1 | TR technique |

**Table S6. Concentrations of brick kiln pollutants reported in studies.**

| Study | Pollutant | Units | Kiln type | Fuels used | Fuel category | Measurement location | Sample size | Mean | SD | Notes |
|---|---|---|---|---|---|---|---|---|---|---|
| Akinshipe 2018 | CO | mg/m$^3$ | CK | Coal | Coal | Stack | 9 | 395 | 607 | Data from Table B excluding Batches 2, 7 and 13. Recalculated pooled SD. |
| Akinshipe 2018 | $NO_x$ | mg/m$^3$ | CK | Coal | Coal | Stack | 9 | 2.7 | 3.2 | |
| Akinshipe 2018 | NO | mg/m$^3$ | CK | Coal | Coal | Stack | 9 | 2.7 | 3.2 | |
| Akinshipe 2018 | $NO_2$ | mg/m$^3$ | CK | Coal | Coal | Stack | 9 | 0 | 0 | |
| Akinshipe 2018 | $CO_2$ | mg/m$^3$ | CK | Coal | Coal | Stack | 9 | 7911 | 12305 | |
| Akinshipe 2018 | $SO_2$ | mg/m$^3$ | CK | Coal | Coal | Stack | 8 | 29.4 | 50.6 | Data from Table B excluding Batches 2, 6, 7 and 13. Recalculated pooled SD. |
| Akinshipe 2018 | $PM_{10}$ | mg/m$^3$ | CK | Coal | Coal | Stack | 9 | 19.5 | 20.3 | Computed from Table B excluding Batches 2, 7 and 13. Recalculated pooled SD. |
| Bashir 2023 | CO | ppm | BTK | Coal | Coal | Stack | 1 | 2349 | NA | |
| Bashir 2023 | $CO_2$ | ppm | BTK | Coal | Coal | Stack | 1 | 85500 | NA | |
| Bashir 2023 | $SO_2$ | ppm | BTK | Coal | Coal | Stack | 1 | 648 | NA | |
| Bashir 2023 | $PM_{2.5}$ | mg/m$^3$ | BTK | Coal | Coal | Stack | 1 | 509.2 | NA | |
| Bashir 2023 | BC | mg/m$^3$ | BTK | Coal | Coal | Stack | 1 | 48.7 | NA | |
| Bashir 2023 | CO | ppm | FDZ | Coal | Coal | Stack | 1 | 718 | NA | |
| Bashir 2023 | $CO_2$ | ppm | FDZ | Coal | Coal | Stack | 1 | 44420 | NA | |
| Bashir 2023 | $SO_2$ | ppm | FDZ | Coal | Coal | Stack | 1 | 27 | NA | |
| Bashir 2023 | $PM_{2.5}$ | mg/m$^3$ | FDZ | Coal | Coal | Stack | 1 | 172.3 | NA | |
| Bashir 2023 | BC | mg/m$^3$ | FDZ | Coal | Coal | Stack | 1 | 3.23 | NA | |
| Beard 2022 | CO | ppm | BTK | NS | NS | Site at brick kiln | 32 | 4.69 | 1.81 | Arithmetic means and standard deviations not reported in paper but provided to us by the authors |
| Beard 2022 | $CO_2$ | ppm | BTK | NS | NS | Site at brick kiln | 32 | 3329 | 2534 | |
| Beard 2022 | silica | μg/m$^3$ | BTK | NS | NS | Site at brick kiln | 31 | 8.75 | 8.16 | |

| | | | | | | | | | | |
|---|---|---|---|---|---|---|---|---|---|---|
| Beard 2022 | $NO_2$ | μg/$m^3$ | BTK | NS | NS | Site at brick kiln | 32 | ND | NA | |
| Beard 2022 | $SO_2$ | μg/$m^3$ | BTK | NS | NS | Site at brick kiln | 32 | ND | NA | |
| Berumen-Rodriguez 2021 | $PM_{10}$ | μg/$m^3$ | NS | NS | NS | BKS | 6 | 1804.3 | NA | Geometric mean |
| Berumen-Rodriguez 2021 | $PM_{2.5}$ | μg/$m^3$ | NS | NS | NS | BKS | 7 | 276.1 | NA | Geometric mean |
| Bruce 2007 | PM | g/$m^3$ | TFK | Wood | Biomass | Stack | 3 | 0.176 | 0.23 | |
| Bruce 2007 | PM | g/$m^3$ | MK | Wood | Biomass | Stack | 6 | 0.137 | 0.25 | |
| Bruce 2007 | PM | g/$m^3$ | TFK | Sawdust | Biomass | Stack | 5 | 0.7 | NA | |
| Bruce 2007 | PM | g/$m^3$ | MK | Sawdust | Biomass | Stack | 5 | 0.51 | 0.12 | |
| Bruce 2007 | Fl | μg/g | MK active | Motor oil | Oil | Stack | 1 | 1105 | NA | |
| Bruce 2007 | Fl | μg/g | MK filter | Motor oil | Oil | Stack | 1 | 299 | NA | |
| Bruce 2007 | Phe | μg/g | MK active | Motor oil | Oil | Stack | 1 | 1235 | NA | |
| Bruce 2007 | Phe | μg/g | MK filter | Motor oil | Oil | Stack | 1 | 182 | NA | |
| Bruce 2007 | Ant | μg/g | MK active | Motor oil | Oil | Stack | 1 | 242 | NA | |
| Bruce 2007 | Ant | μg/g | MK filter | Motor oil | Oil | Stack | 1 | 195 | NA | |
| Bruce 2007 | Fth | μg/g | MK active | Motor oil | Oil | Stack | 1 | 1173 | NA | |
| Bruce 2007 | Fth | μg/g | MK filter | Motor oil | Oil | Stack | 1 | 377 | NA | |
| Bruce 2007 | Py | μg/g | MK active | Motor oil | Oil | Stack | 1 | 2222 | NA | |
| Bruce 2007 | Py | μg/g | MK filter | Motor oil | Oil | Stack | 1 | 337 | NA | |
| Bruce 2007 | B[b]A | μg/g | MK active | Motor oil | Oil | Stack | 1 | 714 | NA | |
| Bruce 2007 | B[b]A | μg/g | MK filter | Motor oil | Oil | Stack | 1 | 581 | NA | |
| Bruce 2007 | Chr | μg/g | MK active | Motor oil | Oil | Stack | 1 | 403 | NA | |
| Bruce 2007 | Chr | μg/g | MK filter | Motor oil | Oil | Stack | 1 | 333 | NA | |
| Bruce 2007 | B[b]F | μg/g | MK active | Motor oil | Oil | Stack | 1 | 4078 | NA | |
| Bruce 2007 | B[b]F | μg/g | MK filter | Motor oil | Oil | Stack | 1 | 2581 | NA | |

| | | | | | | | | | |
|---|---|---|---|---|---|---|---|---|---|
| Bruce 2007 | B[k]F | µg/g | MK active | Motor oil | Oil | Stack | 1 | 1024 | NA |
| Bruce 2007 | B[k]F | µg/g | MK filter | Motor oil | Oil | Stack | 1 | 735 | NA |
| Bruce 2007 | B[a]P | µg/g | MK active | Motor oil | Oil | Stack | 1 | 990 | NA |
| Bruce 2007 | B[a]P | µg/g | MK filter | Motor oil | Oil | Stack | 1 | 581 | NA |
| Bruce 2007 | D[ah]A | µg/g | MK active | Motor oil | Oil | Stack | 1 | 518 | NA |
| Bruce 2007 | D[ah]A | µg/g | MK filter | Motor oil | Oil | Stack | 1 | 300 | NA |
| Bruce 2007 | B[ghi]P | µg/g | MK active | Motor oil | Oil | Stack | 1 | 425 | NA |
| Bruce 2007 | B[ghi]P | µg/g | MK filter | Motor oil | Oil | Stack | 1 | 273 | NA |
| Bruce 2007 | Ind | µg/g | MK active | Motor oil | Oil | Stack | 1 | 953 | NA |
| Bruce 2007 | Ind | µg/g | MK filter | Motor oil | Oil | Stack | 1 | 368 | NA |
| Bruce 2007 | Pb | µg/m$^3$ | MK active | Motor oil | Oil | Stack | 1 | 2.41 | NA |
| Bruce 2007 | Pb | µg/m$^3$ | MK filter | Motor oil | Oil | Stack | 1 | 1.43 | NA |
| Bruce 2007 | Cd | µg/m$^3$ | MK active | Motor oil | Oil | Stack | 1 | ND | NA |
| Bruce 2007 | Cd | µg/m$^3$ | MK filter | Motor oil | Oil | Stack | 1 | ND | NA |
| Bruce 2007 | Ba | µg/m$^3$ | MK active | Motor oil | Oil | Stack | 1 | 28.85 | NA |
| Bruce 2007 | Ba | µg/m$^3$ | MK filter | Motor oil | Oil | Stack | 1 | 14.36 | NA |
| Haque 2018 | $PM_{2.5}$ | mg/m$^3$ | BTK | Coal and biomass | Coal & biomass | Stack | 10 | 141 | 86 |
| Haque 2018 | $PM_{2.5}$ | mg/m$^3$ | ZZK | Coal | Coal | Stack | 6 | 128 | 72 |
| Haque 2018 | $PM_{2.5}$ | mg/m$^3$ | HK | Coal | Coal | Stack | 2 | 109 | 53 |
| Haque 2018 | CO | mg/m$^3$ | BTK | Coal and biomass | Coal & biomass | Stack | 10 | 264 | 75 |
| Haque 2018 | CO | mg/m$^3$ | ZZK | Coal | Coal | Stack | 6 | 177 | 81 |
| Haque 2018 | CO | mg/m$^3$ | HK | Coal | Coal | Stack | 2 | 74 | 21 |
| Haque 2018 | $SO_2$ | mg/m$^3$ | BTK | Coal and biomass | Coal & biomass | Stack | 10 | 578 | 354 |
| Haque 2018 | $SO_2$ | mg/m$^3$ | ZZK | Coal | Coal | Stack | 6 | 332 | 196 |

| | | | | | | | | | | |
|---|---|---|---|---|---|---|---|---|---|---|
| Haque 2018 | $SO_2$ | mg/$m^3$ | HK | Coal | Coal | Stack | 2 | 316 | 219 | |
| Haque 2018 | $CO_2$ | mg/$m^3$ | BTK | Coal and biomass | Coal & biomass | Stack | 10 | 5254 | 2021 | |
| Haque 2018 | $CO_2$ | mg/$m^3$ | ZZK | Coal | Coal | Stack | 6 | 6995 | 2667 | |
| Haque 2018 | $CO_2$ | mg/$m^3$ | HK | Coal | Coal | Stack | 2 | 2350 | 758 | |
| Haque 2018 | VOCs | mg/$m^3$ | BTK | Coal and biomass | Coal & biomass | Stack | 10 | 23204 | 2560 | |
| Haque 2018 | VOCs | mg/$m^3$ | ZZK | Coal | Coal | Stack | 6 | 25266 | 3563 | |
| Haque 2018 | VOCs | mg/$m^3$ | HK | Coal | Coal | Stack | 2 | 22939 | 2760 | |
| Haque 2018 | $NO_x$ | mg/$m^3$ | BTK | Coal and biomass | Coal & biomass | Stack | 10 | 1.6 | 0.75 | |
| Haque 2018 | $NO_x$ | mg/$m^3$ | ZZK | Coal | Coal | Stack | 6 | 1.2 | 0.58 | |
| Haque 2018 | $NO_x$ | mg/$m^3$ | HK | Coal | Coal | Stack | 2 | 0.74 | 0.63 | |
| Iqbal 2013 | B[a]p | mg/Kg | NS | NS | NS | Soot from stack | 1 | ND | NA | Brick kiln Kohat Road |
| Iqbal 2013 | Chr | mg/Kg | NS | NS | NS | Soot from stack | 1 | 71.135 | NA | Brick kiln Kohat Road |
| Iqbal 2013 | Ant | mg/Kg | NS | NS | NS | Soot from stack | 1 | ND | NA | Brick kiln Kohat Road |
| Iqbal 2013 | Fl | mg/Kg | NS | NS | NS | Soot from stack | 1 | 33.392 | NA | Brick kiln Kohat Road |
| Iqbal 2013 | Nap | mg/Kg | NS | NS | NS | Soot from stack | 1 | 70.509 | NA | Brick kiln Kohat Road |
| Iqbal 2013 | Phe | mg/Kg | NS | NS | NS | Soot from stack | 1 | 6.846 | NA | Brick kiln Kohat Road |
| Iqbal 2013 | Acy | mg/Kg | NS | NS | NS | Soot from stack | 1 | 116.952 | NA | Brick kiln Kohat Road |
| Iqbal 2013 | Fth | mg/Kg | NS | NS | NS | Soot from stack | 1 | 111.347 | NA | Brick kiln Kohat Road |
| Iqbal 2013 | B[a]p | mg/Kg | NS | NS | NS | Soot from stack | 1 | ND | NA | Brick kiln 33 |
| Iqbal 2013 | Chr | mg/Kg | NS | NS | NS | Soot from stack | 1 | ND | NA | Brick kiln 33 |
| Iqbal 2013 | Ant | mg/Kg | NS | NS | NS | Soot from stack | 1 | ND | NA | Brick kiln 33 |
| Iqbal 2013 | Fl | mg/Kg | NS | NS | NS | Soot from stack | 1 | ND | NA | Brick kiln 33 |
| Iqbal 2013 | Nap | mg/Kg | NS | NS | NS | Soot from stack | 1 | ND | NA | Brick kiln 33 |

| | | | | | | | | | | |
|---|---|---|---|---|---|---|---|---|---|---|
| Iqbal 2013 | Phe | mg/Kg | NS | NS | NS | Soot from stack | 1 | 366.39 | NA | Brick kiln 33 |
| Iqbal 2013 | Acy | mg/Kg | NS | NS | NS | Soot from stack | 1 | 4078.8 | NA | Brick kiln 33 |
| Iqbal 2013 | Fth | mg/Kg | NS | NS | NS | Soot from stack | 1 | 3.934 | NA | Brick kiln 33 |
| Iqbal 2013 | B[a]p | mg/Kg | NS | NS | NS | Soot from stack | 1 | ND | NA | Brick kiln PM |
| Iqbal 2013 | Chr | mg/Kg | NS | NS | NS | Soot from stack | 1 | ND | NA | Brick kiln PM |
| Iqbal 2013 | Ant | mg/Kg | NS | NS | NS | Soot from stack | 1 | 8.923 | NA | Brick kiln PM |
| Iqbal 2013 | Fl | mg/Kg | NS | NS | NS | Soot from stack | 1 | 1.1492 | NA | Brick kiln PM |
| Iqbal 2013 | Nap | mg/Kg | NS | NS | NS | Soot from stack | 1 | 3.5614 | NA | Brick kiln PM |
| Iqbal 2013 | Phe | mg/Kg | NS | NS | NS | Soot from stack | 1 | 0.2047 | NA | Brick kiln PM |
| Iqbal 2013 | Acy | mg/Kg | NS | NS | NS | Soot from stack | 1 | 0.7653 | NA | Brick kiln PM |
| Iqbal 2013 | Fth | mg/Kg | NS | NS | NS | Soot from stack | 1 | 0.054 | NA | Brick kiln PM |
| Iqbal 2013 | B[a]p | mg/Kg | NS | NS | NS | Soot from stack | 1 | ND | NA | TGR Brick Kiln |
| Iqbal 2013 | Chr | mg/Kg | NS | NS | NS | Soot from stack | 1 | 3.0641 | NA | TGR Brick Kiln |
| Iqbal 2013 | Ant | mg/Kg | NS | NS | NS | Soot from stack | 1 | 13.527 | NA | TGR Brick Kiln |
| Iqbal 2013 | Fl | mg/Kg | NS | NS | NS | Soot from stack | 1 | 3.299 | NA | TGR Brick Kiln |
| Iqbal 2013 | Nap | mg/Kg | NS | NS | NS | Soot from stack | 1 | 4.094 | NA | TGR Brick Kiln |
| Iqbal 2013 | Phe | mg/Kg | NS | NS | NS | Soot from stack | 1 | 0.164 | NA | TGR Brick Kiln |
| Iqbal 2013 | Acy | mg/Kg | NS | NS | NS | Soot from stack | 1 | 0.613 | NA | TGR Brick Kiln |
| Iqbal 2013 | Fth | mg/Kg | NS | NS | NS | Soot from stack | 1 | 18.055 | NA | TGR Brick Kiln |
| Iqbal 2013 | B[a]p | mg/Kg | NS | NS | NS | Soot from stack | 1 | ND | NA | Brick Kiln Achini Bala |
| Iqbal 2013 | Chr | mg/kg | NS | NS | NS | Soot from stack | 1 | 25.7202 | NA | Brick Kiln Achini Bala |
| Iqbal 2013 | Ant | mg/kg | NS | NS | NS | Soot from stack | 1 | 89.105 | NA | Brick Kiln Achini Bala |

| | | | | | | | | | | |
|---|---|---|---|---|---|---|---|---|---|---|
| Iqbal 2013 | Fl | mg/kg | NS | NS | NS | Soot from stack | 1 | 37.1615 | NA | Brick Kiln Achini Bala |
| Iqbal 2013 | Nap | mg/kg | NS | NS | NS | Soot from stack | 1 | 55.696 | NA | Brick Kiln Achini Bala |
| Iqbal 2013 | Phe | mg/kg | NS | NS | NS | Soot from stack | 1 | 0.866 | NA | Brick Kiln Achini Bala |
| Iqbal 2013 | Acy | mg/kg | NS | NS | NS | Soot from stack | 1 | 27.619 | NA | Brick Kiln Achini Bala |
| Iqbal 2013 | Fth | mg/kg | NS | NS | NS | Soot from stack | 1 | 32.794 | NA | Brick Kiln Achini Bala |
| Jan 2014 | BaP | mg/kg | NS | NS | NS | Soil dust at kiln | NA | 0.0026 | NA | |
| Jan 2014 | Chr | mg/kg | NS | NS | NS | Soil dust at kiln | NA | 0.0646 | NA | |
| Jan 2014 | Ant | mg/kg | NS | NS | NS | Soil dust at kiln | NA | 0.0044 | NA | |
| Jan 2014 | Fl | mg/kg | NS | NS | NS | Soil dust at kiln | NA | ND | NA | |
| Jan 2014 | Nap | mg/kg | NS | NS | NS | Soil dust at kiln | NA | 0.2167 | NA | |
| Jan 2014 | Phe | mg/kg | NS | NS | NS | Soil dust at kiln | NA | ND | NA | |
| Jan 2014 | Ace | mg/kg | NS | NS | NS | Soil dust at kiln | NA | 0.1127 | NA | |
| Jan 2014 | Fth | mg/kg | NS | NS | NS | Soil dust at kiln | NA | ND | NA | |
| Kamal 2014a | Nap | ng/g | NS | NS | NS | Soil dust at kiln | 26 | 132 | 126 | |
| Kamal 2014a | Ace | ng/g | NS | NS | NS | Soil dust at kiln | 26 | 35.5 | 25.9 | |
| Kamal 2014a | Acy | ng/g | NS | NS | NS | Soil dust at kiln | 26 | 38.3 | 14.4 | |
| Kamal 2014a | Ant | ng/g | NS | NS | NS | Soil dust at kiln | 26 | 95.2 | 79.5 | |
| Kamal 2014a | Fl | ng/g | NS | NS | NS | Soil dust at kiln | 26 | 47.5 | 29.4 | |
| Kamal 2014a | Phe | ng/g | NS | NS | NS | Soil dust at kiln | 26 | 301 | 237 | |
| Kamal 2014a | B[a]A | ng/g | NS | NS | NS | Soil dust at kiln | 26 | 40.4 | 21.5 | |
| Kamal 2014a | Chr | ng/g | NS | NS | NS | Soil dust at kiln | 26 | 170 | 97.2 | |
| Kamal 2014a | Fth | ng/g | NS | NS | NS | Soil dust at kiln | 26 | 239 | 151 | |

| | | | | | | | | | | |
|---|---|---|---|---|---|---|---|---|---|---|
| Kamal 2014a | Py | ng/g | NS | NS | NS | Soil dust at kiln | 26 | 179 | 138 | |
| Kamal 2014a | B[a]P | ng/g | NS | NS | NS | Soil dust at kiln | 26 | 102 | 90.6 | |
| Kamal 2014a | B[b+k]F | ng/g | NS | NS | NS | Soil dust at kiln | 26 | 117 | 99.8 | |
| Kamal 2014a | D[a,h]A | ng/g | NS | NS | NS | Soil dust at kiln | 26 | 61.4 | 40 | |
| Kamal 2014a | B[e]P | ng/g | NS | NS | NS | Soil dust at kiln | 26 | 70.9 | 64.8 | |
| Kamal 2014a | Ind | ng/g | NS | NS | NS | Soil dust at kiln | 26 | 46.1 | 27.2 | |
| Kamal 2014a | B[ghi]P | ng/g | NS | NS | NS | Soil dust at kiln | 26 | 47.9 | 26.6 | |
| Kamal 2014a | Cor | ng/g | NS | NS | NS | Soil dust at kiln | 26 | 9.1 | 3.91 | |
| Kamal 2014a | Total PAHs | ng/g | NS | NS | NS | Soil dust at kiln | 26 | 1528 | 1416 | |
| Le 2010 | CO | $mg/m^3$ | TIK | Coal briquettes | Coal | Stack | 2-6 (not specified) | 4 | 1.4 | Day 1 |
| Le 2010 | CO | $mg/m^3$ | TIK | Coal briquettes | Coal | Stack | 2-6 (not specified) | 1867 | 18 | Day 2 |
| Le 2010 | CO | $mg/m^3$ | TIK | Coal briquettes | Coal | Stack | 2-6 (not specified) | 1770 | 36 | Day 3 |
| Le 2010 | CO | $mg/m^3$ | TIK | Coal briquettes | Coal | Stack | 2-6 (not specified) | 1277 | 30 | Day 4 |
| Le 2010 | CO | $mg/m^3$ | TIK | Coal briquettes | Coal | Stack | 2-6 (not specified) | 1712 | 355 | Day 5 |
| Le 2010 | CO | $mg/m^3$ | TIK | Coal briquettes | Coal | Stack | 2-6 (not specified) | 1212 | 139 | Day 6 |
| Le 2010 | CO | $mg/m^3$ | TIK | Coal briquettes | Coal | Stack | 2-6 (not specified) | 339 | 32 | Day 7 |
| Le 2010 | $SO_2$ | $mg/m^3$ | TIK | Coal briquettes | Coal | Stack | 2-6 (not specified) | 2.9 | 0 | Day 1 |
| Le 2010 | $SO_2$ | $mg/m^3$ | TIK | Coal briquettes | Coal | Stack | 2-6 (not specified) | 23 | 4.2 | Day 2 |
| Le 2010 | $SO_2$ | $mg/m^3$ | TIK | Coal briquettes | Coal | Stack | 2-6 (not specified) | 11 | 0 | Day 3 |
| Le 2010 | $SO_2$ | $mg/m^3$ | TIK | Coal briquettes | Coal | Stack | 2-6 (not specified) | 6 | 0 | Day 4 |
| Le 2010 | $SO_2$ | $mg/m^3$ | TIK | Coal briquettes | Coal | Stack | 2-6 (not specified) | 150 | 3 | Day 5 |
| Le 2010 | $SO_2$ | $mg/m^3$ | TIK | Coal briquettes | Coal | Stack | 2-6 (not specified) | 199 | 17 | Day 6 |

| | | | | | | | | | | |
|---|---|---|---|---|---|---|---|---|---|---|
| Le 2010 | $SO_2$ | $mg/m^3$ | TIK | Coal briquettes | Coal | Stack | 2-6 (not specified) | 179 | 6.1 | Day 7 |
| Le 2010 | PM | $mg/m^3$ | TIK | Coal briquettes | Coal | Stack | 2-6 (not specified) | 13 | NA | Day 1 |
| Le 2010 | PM | $mg/m^3$ | TIK | Coal briquettes | Coal | Stack | 2-6 (not specified) | 36 | NA | Day 2 |
| Le 2010 | PM | $mg/m^3$ | TIK | Coal briquettes | Coal | Stack | 2-6 (not specified) | 108 | NA | Day 3 |
| Le 2010 | PM | $mg/m^3$ | TIK | Coal briquettes | Coal | Stack | 2-6 (not specified) | 105 | NA | Day 4 |
| Le 2010 | PM | $mg/m^3$ | TIK | Coal briquettes | Coal | Stack | 2-6 (not specified) | 157 | NA | Day 5 |
| Le 2010 | PM | $mg/m^3$ | TIK | Coal briquettes | Coal | Stack | 2-6 (not specified) | 367 | NA | Day 6 |
| Le 2010 | PM | $mg/m^3$ | TIK | Coal briquettes | Coal | Stack | 2-6 (not specified) | 17 | NA | Day 7 |
| Mondal 2017 | Cd | mg/kg | NS | NS | NS | Bottom ash | 20 | 9.8 | NA | West Bengal |
| Mondal 2017 | Cr | mg/kg | NS | NS | NS | Bottom ash | 20 | 56.8 | NA | West Bengal |
| Mondal 2017 | Pb | mg/kg | NS | NS | NS | Bottom ash | 20 | 240.6 | NA | West Bengal |
| Mondal 2017 | Mn | mg/kg | NS | NS | NS | Bottom ash | 20 | 540.9 | NA | West Bengal |
| Mondal 2017 | Cu | mg/kg | NS | NS | NS | Bottom ash | 20 | 93.2 | NA | West Bengal |
| Mondal 2017 | Zn | mg/kg | NS | NS | NS | Bottom ash | 20 | 253.4 | NA | West Bengal |
| Mondal 2017 | Cd | mg/kg | NS | NS | NS | Bottom ash | 12 | 6.5 | NA | Assam |
| Mondal 2017 | Cr | mg/kg | NS | NS | NS | Bottom ash | 12 | 89.9 | NA | Assam |
| Mondal 2017 | Pb | mg/kg | NS | NS | NS | Bottom ash | 12 | 234.6 | NA | Assam |
| Mondal 2017 | Mn | mg/kg | NS | NS | NS | Bottom ash | 12 | 206.2 | NA | Assam |
| Mondal 2017 | Cu | mg/kg | NS | NS | NS | Bottom ash | 12 | 73.0 | NA | Assam |
| Mondal 2017 | Zn | mg/kg | NS | NS | NS | Bottom ash | 12 | 251.6 | NA | Assam |
| Nepal 2019 | $CO_2$ | $mg/m^3$ | BTK | Coal, rice husk, briquette | Coal & biomass | Stack | 4 | 46175 | 6947 | |
| Nepal 2019 | $SO_2$ | $mg/m^3$ | BTK | Coal, rice husk, briquette | Coal & biomass | Stack | 4 | 735 | 728 | |
| Nepal 2019 | $PM_{2.5}$ | $mg/m^3$ | BTK | Coal, rice husk, briquette | Coal & biomass | Stack | 4 | 303 | 152 | |
| Nepal 2019 | BC | $mg/m^3$ | BTK | Coal, rice husk, briquette | Coal & biomass | Stack | 4 | 55.5 | 16.4 | |

| | | | | | | | | | | |
|---|---|---|---|---|---|---|---|---|---|---|
| Nepal 2019 | $CO_2$ | mg/m$^3$ | FDZ | Coal, rice husk, sawdust | Coal & biomass | Stack | 3 | 35639 | 3337 | |
| Nepal 2019 | $SO_2$ | mg/m$^3$ | FDZ | Coal, rice husk, sawdust | Coal & biomass | Stack | 3 | 371 | 299 | |
| Nepal 2019 | $PM_{2.5}$ | mg/m$^3$ | FDZ | Coal, rice husk, sawdust | Coal & biomass | Stack | 3 | 148 | 61 | |
| Nepal 2019 | BC | mg/m$^3$ | FDZ | Coal, rice husk, sawdust | Coal & biomass | Stack | 3 | 21.7 | 15.5 | |
| Ortinez-Alvarez 2018 | $PM_{2.5}$ | µg/m$^3$ | TFK | Pine wood | Biomass | Flue gas | 9 | 8.39 | NA | sampling from crane system on top of kiln domes |
| Ortinez-Alvarez 2018 | EC | µg/m$^3$ | TFK | Pine wood | Biomass | Flue gas | 9 | 0.67 | NA | sampling from crane system on top of kiln domes |
| Ortinez-Alvarez 2018 | OC | µg/m$^3$ | TFK | Pine wood | Biomass | Flue gas | 9 | 4.57 | NA | sampling from crane system on top of kiln domes |
| Ortinez-Alvarez 2018 | $PM_{2.5}$ | µg/m$^3$ | MK2 | Pine wood | Biomass | Flue gas | 9 | 4.07 | NA | sampling from crane system on top of kiln domes |
| Ortinez-Alvarez 2018 | EC | µg/m$^3$ | MK2 | Pine wood | Biomass | Flue gas | 9 | 0.28 | NA | sampling from crane system on top of kiln domes |
| Ortinez-Alvarez 2018 | OC | µg/m$^3$ | MK2 | Pine wood | Biomass | Flue gas | 9 | 2.24 | NA | sampling from crane system on top of kiln domes |
| Pangtey 2004 | $SO_2$ | ppm | BTK | Coal, firewood | Coal & biomass | Site at brick kiln | 30 | 0.35 | NA | |
| Pangtey 2004 | $NO_x$ | ppm | BTK | Coal, firewood | Coal & biomass | Site at brick kiln | 30 | 0.23 | NA | |
| Pangtey 2004 | CO | ppm | BTK | Coal, firewood | Coal & biomass | Site at brick kiln | 30 | 3.44 | NA | |
| Pangtey 2004 | TSP | ppm | BTK | Coal, firewood | Coal & biomass | Site at brick kiln | 30 | 93.3 | NA | |
| Pangtey 2004 | RSP | ppm | BTK | Coal, firewood | Coal & biomass | Site at brick kiln | 30 | 4.66 | NA | |
| Pangtey 2004 | Cd | µg/g | BTK | Coal, fire wood | Coal & biomass | Soil dust at kiln | 11 | 1.05 | NA | |

| | | | | | | | | | | |
|---|---|---|---|---|---|---|---|---|---|---|
| Pangtey 2004 | Cr | µg/g | BTK | Coal, fire wood | Coal & biomass | Soil dust at kiln | 11 | 6.19 | NA | |
| Pangtey 2004 | Cu | µg/g | BTK | Coal, fire wood | Coal & biomass | Soil dust at kiln | 11 | 8.82 | NA | |
| Pangtey 2004 | Fe | µg/g | BTK | Coal, fire wood | Coal & biomass | Soil dust at kiln | 11 | 4494.6 | NA | |
| Pangtey 2004 | Mn | µg/g | BTK | Coal, fire wood | Coal & biomass | Soil dust at kiln | 11 | 230.03 | NA | |
| Pangtey 2004 | Ni | µg/g | BTK | Coal, fire wood | Coal & biomass | Soil dust at kiln | 11 | 14.29 | NA | |
| Pangtey 2004 | Pb | µg/g | BTK | Coal, fire wood | Coal & biomass | Soil dust at kiln | 11 | 30.04 | NA | |
| Pangtey 2004 | Zn | µg/g | BTK | Coal, fire wood | Coal & biomass | Soil dust at kiln | 11 | 28.66 | NA | |
| Ravankhah 2017 | Cd | mg/kg | traditional | Coal, wood, furnace oil | Coal & biomass | Soil dust | 12 | 0.79 | 0.20 | 0-500 m from kiln |
| Ravankhah 2017 | Pb | mg/kg | traditional | Coal, wood, furnace oil | Coal & biomass | Soil dust | 12 | 11.78 | 2.44 | 0-500 m from kiln |
| Ravankhah 2017 | Ni | mg/kg | traditional | Coal, wood, furnace oil | Coal & biomass | Soil dust | 12 | 20.14 | 7.02 | 0-500 m from kiln |
| Ravankhah 2017 | Zn | mg/kg | traditional | Coal, wood, furnace oil | Coal & biomass | Soil dust | 12 | 35.41 | 6.30 | 0-500 m from kiln |
| Ravankhah 2017 | Cu | mg/kg | traditional | Coal, wood, furnace oil | Coal & biomass | Soil dust | 12 | 6.75 | 4.10 | 0-500 m from kiln |
| Raza 2014 | $PM_{2.5}$ | $mg/m^3$ | NS | NS | NS | Site at brick kiln | NA | 0.301 | NA | modulation and loading |
| Raza 2014 | $PM_{2.5}$ | $mg/m^3$ | NS | NS | NS | Site at brick kiln | NA | 0.307 | NA | burning |
| Raza 2014 | $PM_{2.5}$ | $mg/m^3$ | NS | NS | NS | Site at brick kiln | NA | 0.628 | NA | unloading |
| Raza 2014 | $PM_{10}$ | $mg/m^3$ | NS | NS | NS | Site at brick kiln | NA | 0.888 | NA | modulation and loading |
| Raza 2014 | $PM_{10}$ | $mg/m^3$ | NS | NS | NS | Site at brick kiln | NA | 1.83 | NA | burning |
| Raza 2014 | $PM_{10}$ | $mg/m^3$ | NS | NS | NS | Site at brick kiln | NA | 0.861 | NA | unloading |
| Raza 2021 | $PM_{2.5}$ | $\mu g/m^3$ | BTK | Anthracite | Coal | Site at brick kiln | 3 | 48.58 | 26.05 | modulation area |
| Raza 2021 | $PM_{10}$ | $\mu g/m^3$ | BTK | Anthracite | Coal | Site at brick kiln | 3 | 656.36 | 26.05 | modulation area |
| Raza 2021 | $SO_2$ | ppm | BTK | Anthracite | Coal | Site at brick kiln | 3 | 0 | 0 | modulation area |
| Raza 2021 | $NO_2$ | ppm | BTK | Anthracite | Coal | Site at brick kiln | 3 | 0.0591 | 0.013 | modulation area |

| | | | | | | | | | | |
|---|---|---|---|---|---|---|---|---|---|---|
| Raza 2021 | VOCs | ppm | BTK | Anthracite | Coal | Site at brick kiln | 3 | 1303.08 | 388.41 | modulation area |
| Raza 2021 | $PM_{2.5}$ | μg/m$^3$ | BTK | Anthracite | Coal | Site at brick kiln | 3 | 100.34 | 27.22 | kiln area |
| Raza 2021 | $PM_{10}$ | μg/m$^3$ | BTK | Anthracite | Coal | Site at brick kiln | 3 | 1327.55 | 388.98 | kiln area |
| Raza 2021 | $SO_2$ | ppm | BTK | Anthracite | Coal | Site at brick kiln | 3 | 0.0652 | 0.018 | kiln area |
| Raza 2021 | $NO_2$ | ppm | BTK | Anthracite | Coal | Site at brick kiln | 3 | 0.07 | 0.007 | kiln area |
| Raza 2021 | VOCs | ppm | BTK | Anthracite | Coal | Site at brick kiln | 3 | 1106.04 | 338.12 | kiln area |
| Saikia 2018 | $PM_{2.5}$ | μg/m$^3$ | NS | Coal | Coal | Site at brick kiln | NA | 27.4 | NA | |
| Saikia 2018 | $PM_{10}$ | μg/m$^3$ | NS | Coal | Coal | Site at brick kiln | NA | 21.7 | NA | |
| Saikia 2018 | $SO_2$ | μg/m$^3$ | NS | Coal | Coal | Site at brick kiln | NA | 23.5 | NA | |
| Saikia 2018 | $NO_2$ | μg/m$^3$ | NS | Coal | Coal | Site at brick kiln | NA | 10.9 | NA | |
| Saikia 2018 | $PM_{2.5}$ | μg/m$^3$ | NS | Coal | Coal | Site at brick kiln | 8 | 27.4 | NA | |
| Saikia 2018 | $SO_2$ | μg/m$^3$ | NS | Coal | Coal | Site at brick kiln | 8 | 23.5 | NA | |
| Saikia 2018 | $NO_2$ | μg/m$^3$ | NS | Coal | Coal | Site at brick kiln | 8 | 11.1 | NA | |
| Saikia 2018 | $NH_3$ | μg/m$^3$ | NS | Coal | Coal | Site at brick kiln | 8 | 28.1 | NA | |
| Saikia 2018 | Fl | ng/m$^3$ | NS | Coal | Coal | Site at brick kiln | 8 | 56.3 | NA | PAHs in $PM_{2.5}$ |
| Saikia 2018 | Nap | ng/m$^3$ | NS | Coal | Coal | Site at brick kiln | 8 | 18.7 | NA | PAHs in $PM_{2.5}$ |
| Saikia 2018 | Ace | ng/m$^3$ | NS | Coal | Coal | Site at brick kiln | 8 | 18.2 | NA | PAHs in $PM_{2.5}$ |
| Saikia 2018 | Acy | ng/m$^3$ | NS | Coal | Coal | Site at brick kiln | 8 | 17 | NA | PAHs in $PM_{2.5}$ |
| Sanjel 2016 | $PM_{10}$ | mg/m$^3$ | NS | NS | NS | Site at brick kiln | 80 | 4.958 | NA | |
| Sanjel 2016 | $PM_{2.5}$ | mg/m$^3$ | BTK | NS | NS | Site at brick kiln | 80 | 3.965 | NA | |
| Sughis 2014 | $PM_{2.5}$ | μg/m$^3$ | NS | NS | NS | Site at brick kiln | NA | 123.8 | 25.8 | |
| Sughis 2014 | $PM_{10}$ | μg/m$^3$ | NS | NS | NS | Site at brick kiln | NA | 814 | 134 | |

| | | | | | | | | | |
|---|---|---|---|---|---|---|---|---|---|
| Tabinda 2019 | CO | ppm | BTK | High-quality coal | Coal | Flue gas | 2 | 1 | 0.003 |
| Tabinda 2019 | CO | ppm | BTK | High-quality coal | Coal | Stack | 2 | 108 | 0.9 |
| Tabinda 2019 | $NO_x$ | ppm | BTK | High-quality coal | Coal | Flue gas | 2 | 0 | 0 |
| Tabinda 2019 | $NO_x$ | ppm | BTK | High-quality coal | Coal | Stack | 2 | 0 | 0 |
| Tabinda 2019 | $NO_2$ | ppm | BTK | High-quality coal | Coal | Flue gas | 2 | 0 | 0 |
| Tabinda 2019 | $NO_2$ | ppm | BTK | High-quality coal | Coal | Stack | 2 | 0 | 0 |
| Tabinda 2019 | NO | ppm | BTK | High-quality coal | Coal | Flue gas | 2 | 0 | 0 |
| Tabinda 2019 | NO | ppm | BTK | High-quality coal | Coal | Stack | 2 | 0 | 0 |
| Tabinda 2019 | $CO_2$ | ppm | BTK | High-quality coal | Coal | Flue gas | 2 | 0 | 0 |
| Tabinda 2019 | $CO_2$ | ppm | BTK | High-quality coal | Coal | Stack | 2 | 4.54 | 0.03 |
| Tabinda 2019 | $SO_2$ | ppm | BTK | High-quality coal | Coal | Flue gas | 2 | 0 | 0 |
| Tabinda 2019 | $SO_2$ | ppm | BTK | High-quality coal | Coal | Stack | 2 | 6 | 0.07 |
| Tabinda 2019 | CO | ppm | BTK | Low-quality coal & wood | Coal & biomass | Flue gas | 2 | 0 | 0 |
| Tabinda 2019 | CO | ppm | BTK | Low-quality coal & wood | Coal & biomass | Stack | 2 | 82 | 0.98 |
| Tabinda 2019 | $NO_x$ | ppm | BTK | Low-quality coal & wood | Coal & biomass | Flue gas | 2 | 0.1 | 0 |
| Tabinda 2019 | $NO_x$ | ppm | BTK | Low-quality coal & wood | Coal & biomass | Stack | 2 | 19 | 0.43 |
| Tabinda 2019 | $NO_2$ | ppm | BTK | Low-quality coal & wood | Coal & biomass | Flue gas | 2 | 0.1 | 0 |
| Tabinda 2019 | $NO_2$ | ppm | BTK | Low-quality coal & wood | Coal & biomass | Stack | 2 | 0 | 0 |
| Tabinda 2019 | NO | ppm | BTK | Low-quality coal & wood | Coal & biomass | Flue gas | 2 | 0 | 0 |
| Tabinda 2019 | NO | ppm | BTK | Low-quality coal & wood | Coal & biomass | Stack | 2 | 19 | 0.53 |
| Tabinda 2019 | $CO_2$ | ppm | BTK | Low-quality coal & wood | Coal & biomass | Flue gas | 2 | 0 | 0 |
| Tabinda 2019 | $CO_2$ | ppm | BTK | Low-quality coal & wood | Coal & biomass | Stack | 2 | 6.26 | 0.08 |

| | | | | | | | | | | |
|---|---|---|---|---|---|---|---|---|---|---|
| Tabinda 2019 | $SO_2$ | ppm | BTK | Low-quality coal & wood | Coal & biomass | Flue gas | 2 | 0 | 0 | |
| Tabinda 2019 | $SO_2$ | ppm | BTK | Low-quality coal & wood | Coal & biomass | Stack | 2 | 1955 | 3.76 | |
| Tandon 2017 | $SO_X$ | $\mu g/m^3$ | NS | NS | NS | Site at brick kiln | NA | 91.76 | 12.32 | |
| Tandon 2017 | $NO_X$ | $\mu g/m^3$ | NS | NS | NS | Site at brick kiln | NA | 96 | 40 | |
| Tandon 2017 | TSP | $\mu g/m^3$ | NS | NS | NS | Site at brick kiln | NA | 133 | 18.76 | |
| Tandon 2017 | CO | $\mu g/m^3$ | NS | NS | NS | Site at brick kiln | NA | 2.02 | 0.82 | |
| Tandon 2017 | $CO_2$ | $\mu g/m^3$ | NS | NS | NS | Site at brick kiln | NA | 0.89 | 0.32 | |
| Thygerson 2019 | $PM_{2.5}$ | $\mu g/m^3$ | NS | NS | NS | Site at brick kiln | 4 | 184.65 | NA | sampling from houses at kiln site |
| Ubaque 2010 | PM | $mg/m^3$ | HK | Coal & solid waste | Coal & solid waste | Stack | 3 | 27.29 | NA | MP 1 |
| Ubaque 2010 | PM | $mg/m^3$ | HK | Coal & solid waste | Coal & solid waste | Stack | 3 | 40.07 | NA | MP 2 |
| Ubaque 2010 | PM | $mg/m^3$ | HK | Coal & solid waste | Coal & solid waste | Stack | 3 | 17.38 | NA | MP 3 |
| Ubaque 2010 | PM | $mg/m^3$ | HK | Coal & solid waste | Coal & solid waste | Stack | 3 | 24.2 | NA | MP 4 |
| Ubaque 2010 | PM | $mg/m^3$ | HK | Coal & solid waste | Coal & solid waste | Stack | 3 | 18.86 | NA | MP 5 |
| Ubaque 2010 | PM | $mg/m^3$ | HK | Coal & solid waste | Coal & solid waste | Stack | 3 | 39.62 | NA | MP 6 |
| Ubaque 2010 | $SO_2$ | $mg/m^3$ | HK | Coal & solid waste | Coal & solid waste | Stack | 3 | 78 | NA | MP 1 |
| Ubaque 2010 | $SO_2$ | $mg/m^3$ | HK | Coal & solid waste | Coal & solid waste | Stack | 3 | 37.75 | NA | MP 2 |
| Ubaque 2010 | $SO_2$ | $mg/m^3$ | HK | Coal & solid waste | Coal & solid waste | Stack | 3 | 12.08 | NA | MP 3 |

| | | | | | | | | | | |
|---|---|---|---|---|---|---|---|---|---|---|
| Ubaque 2010 | $SO_2$ | $mg/m^3$ | HK | Coal & solid waste | Coal & solid waste | Stack | 3 | 97.54 | NA | MP 4 |
| Ubaque 2010 | $SO_2$ | $mg/m^3$ | HK | Coal & solid waste | Coal & solid waste | Stack | 3 | 28.92 | NA | MP 5 |
| Ubaque 2010 | $SO_2$ | $mg/m^3$ | HK | Coal & solid waste | Coal & solid waste | Stack | 3 | 25.71 | NA | MP 6 |
| Ubaque 2010 | $NO_x$ | $mg/m^3$ | HK | Coal & solid waste | Coal & solid waste | Stack | 3 | 15.72 | NA | MP 1 |
| Ubaque 2010 | $NO_x$ | $mg/m^3$ | HK | Coal & solid waste | Coal & solid waste | Stack | 3 | 13.32 | NA | MP 2 |
| Ubaque 2010 | $NO_x$ | $mg/m^3$ | HK | Coal & solid waste | Coal & solid waste | Stack | 3 | 12.84 | NA | MP 3 |
| Ubaque 2010 | $NO_x$ | $mg/m^3$ | HK | Coal & solid waste | Coal & solid waste | Stack | 3 | 16.13 | NA | MP 4 |
| Ubaque 2010 | $NO_x$ | $mg/m^3$ | HK | Coal & solid waste | Coal & solid waste | Stack | 3 | 21.69 | NA | MP 5 |
| Ubaque 2010 | $NO_x$ | $mg/m^3$ | HK | Coal & solid waste | Coal & solid waste | Stack | 3 | 18.76 | NA | MP 6 |
| Ubaque 2010 | $CH_4$ | $mg/m^3$ | HK | Coal & solid waste | Coal & solid waste | Stack | 3 | 9.27 | NA | MP 1 |
| Ubaque 2010 | $CH_4$ | $mg/m^3$ | HK | Coal & solid waste | Coal & solid waste | Stack | 3 | 12.75 | NA | MP 2 |
| Ubaque 2010 | $CH_4$ | $mg/m^3$ | HK | Coal & solid waste | Coal & solid waste | Stack | 3 | 21.43 | NA | MP 3 |
| Ubaque 2010 | $CH_4$ | $mg/m^3$ | HK | Coal & solid waste | Coal & solid waste | Stack | 3 | 18.7 | NA | MP 4 |
| Ubaque 2010 | $CH_4$ | $mg/m^3$ | HK | Coal & solid waste | Coal & solid waste | Stack | 3 | 22.17 | NA | MP 5 |

| | | | | | | | | | | |
|---|---|---|---|---|---|---|---|---|---|---|
| Ubaque 2010 | $CH_4$ | mg/m$^3$ | HK | Coal & solid waste | Coal & solid waste | Stack | 3 | 23.89 | NA | MP 6 |
| Ubaque 2010 | Heavy metals | mg/m$^3$ | HK | Coal & solid waste | Coal & solid waste | Stack | 3 | 0.0984 | NA | MP 1 |
| Ubaque 2010 | Heavy metals | mg/m$^3$ | HK | Coal & solid waste | Coal & solid waste | Stack | 3 | 0.1055 | NA | MP 2 |
| Ubaque 2010 | Heavy metals | mg/m$^3$ | HK | Coal & solid waste | Coal & solid waste | Stack | 3 | 0.1111 | NA | MP 3 |
| Ubaque 2010 | Heavy metals | mg/m$^3$ | HK | Coal & solid waste | Coal & solid waste | Stack | 3 | 0.0826 | NA | MP 4 |
| Ubaque 2010 | Heavy metals | mg/m$^3$ | HK | Coal & solid waste | Coal & solid waste | Stack | 3 | 0.0697 | NA | MP 5 |
| Ubaque 2010 | Heavy metals | mg/m$^3$ | HK | Coal & solid waste | Coal & solid waste | Stack | 3 | 0.1007 | NA | MP 6 |
| Ubaque 2010 | Sb | mg/m$^3$ | HK | Coal & solid waste | Coal & solid waste | Stack | 3 | 0.0010 | NA | MP 1 |
| Ubaque 2010 | Sb | mg/m$^3$ | HK | Coal & solid waste | Coal & solid waste | Stack | 3 | 0.0008 | NA | MP 2 |
| Ubaque 2010 | Sb | mg/m$^3$ | HK | Coal & solid waste | Coal & solid waste | Stack | 3 | 0.0018 | NA | MP 3 |
| Ubaque 2010 | Sb | mg/m$^3$ | HK | Coal & solid waste | Coal & solid waste | Stack | 3 | 0.0011 | NA | MP 4 |
| Ubaque 2010 | Sb | mg/m$^3$ | HK | Coal & solid waste | Coal & solid waste | Stack | 3 | 0.0008 | NA | MP 5 |
| Ubaque 2010 | Sb | mg/m$^3$ | HK | Coal & solid waste | Coal & solid waste | Stack | 3 | 0.0008 | NA | MP 6 |
| Ubaque 2010 | As | mg/m$^3$ | HK | Coal & solid waste | Coal & solid waste | Stack | 3 | 0.0004 | NA | MP 1 |

| | | | | | | | | | | |
|---|---|---|---|---|---|---|---|---|---|---|
| Ubaque 2010 | As | mg/m³ | HK | Coal & solid waste | Coal & solid waste | Stack | 3 | 0.0003 | NA | MP 2 |
| Ubaque 2010 | As | mg/m³ | HK | Coal & solid waste | Coal & solid waste | Stack | 3 | 0.0012 | NA | MP 3 |
| Ubaque 2010 | As | mg/m³ | HK | Coal & solid waste | Coal & solid waste | Stack | 3 | 0.0022 | NA | MP 4 |
| Ubaque 2010 | As | mg/m³ | HK | Coal & solid waste | Coal & solid waste | Stack | 3 | 0.0002 | NA | MP 5 |
| Ubaque 2010 | As | mg/m³ | HK | Coal & solid waste | Coal & solid waste | Stack | 3 | 0.0001 | NA | MP 6 |
| Ubaque 2010 | Cd | mg/m³ | HK | Coal & solid waste | Coal & solid waste | Stack | 3 | 0.0010 | NA | MP 1 |
| Ubaque 2010 | Cd | mg/m³ | HK | Coal & solid waste | Coal & solid waste | Stack | 3 | 0.0008 | NA | MP 2 |
| Ubaque 2010 | Cd | mg/m³ | HK | Coal & solid waste | Coal & solid waste | Stack | 3 | 0.0017 | NA | MP 3 |
| Ubaque 2010 | Cd | mg/m³ | HK | Coal & solid waste | Coal & solid waste | Stack | 3 | 0.0005 | NA | MP 4 |
| Ubaque 2010 | Cd | mg/m³ | HK | Coal & solid waste | Coal & solid waste | Stack | 3 | 0.0008 | NA | MP 5 |
| Ubaque 2010 | Cd | mg/m³ | HK | Coal & solid waste | Coal & solid waste | Stack | 3 | 0.0005 | NA | MP 6 |
| Ubaque 2010 | Co | mg/m³ | HK | Coal & solid waste | Coal & solid waste | Stack | 3 | 0.054 | NA | MP 1 |
| Ubaque 2010 | Co | mg/m³ | HK | Coal & solid waste | Coal & solid waste | Stack | 3 | 0.045 | NA | MP 2 |
| Ubaque 2010 | Co | mg/m³ | HK | Coal & solid waste | Coal & solid waste | Stack | 3 | 0.040 | NA | MP 3 |

| | | | | | | | | | | |
|---|---|---|---|---|---|---|---|---|---|---|
| Ubaque 2010 | Co | mg/m$^3$ | HK | Coal & solid waste | Coal & solid waste | Stack | 3 | 0.028 | NA | MP 4 |
| Ubaque 2010 | Co | mg/m$^3$ | HK | Coal & solid waste | Coal & solid waste | Stack | 3 | 0.009 | NA | MP 5 |
| Ubaque 2010 | Co | mg/m$^3$ | HK | Coal & solid waste | Coal & solid waste | Stack | 3 | 0.015 | NA | MP 6 |
| Ubaque 2010 | Cu | mg/m$^3$ | HK | Coal & solid waste | Coal & solid waste | Stack | 3 | 0.004 | NA | MP 1 |
| Ubaque 2010 | Cu | mg/m$^3$ | HK | Coal & solid waste | Coal & solid waste | Stack | 3 | 0.003 | NA | MP 2 |
| Ubaque 2010 | Cu | mg/m$^3$ | HK | Coal & solid waste | Coal & solid waste | Stack | 3 | 0.010 | NA | MP 3 |
| Ubaque 2010 | Cu | mg/m$^3$ | HK | Coal & solid waste | Coal & solid waste | Stack | 3 | 0.013 | NA | MP 4 |
| Ubaque 2010 | Cu | mg/m$^3$ | HK | Coal & solid waste | Coal & solid waste | Stack | 3 | 0.009 | NA | MP 5 |
| Ubaque 2010 | Cu | mg/m$^3$ | HK | Coal & solid waste | Coal & solid waste | Stack | 3 | 0.003 | NA | MP 6 |
| Ubaque 2010 | Cr | mg/m$^3$ | HK | Coal & solid waste | Coal & solid waste | Stack | 3 | 0.007 | NA | MP 1 |
| Ubaque 2010 | Cr | mg/m$^3$ | HK | Coal & solid waste | Coal & solid waste | Stack | 3 | 0.005 | NA | MP 2 |
| Ubaque 2010 | Cr | mg/m$^3$ | HK | Coal & solid waste | Coal & solid waste | Stack | 3 | 0.001 | NA | MP 3 |
| Ubaque 2010 | Cr | mg/m$^3$ | HK | Coal & solid waste | Coal & solid waste | Stack | 3 | 0.002 | NA | MP 4 |
| Ubaque 2010 | Cr | mg/m$^3$ | HK | Coal & solid waste | Coal & solid waste | Stack | 3 | 0.004 | NA | MP 5 |

| | | | | | | | | | | |
|---|---|---|---|---|---|---|---|---|---|---|
| Ubaque 2010 | Cr | mg/m$^3$ | HK | Coal & solid waste | Coal & solid waste | Stack | 3 | 0.007 | NA | MP 6 |
| Ubaque 2010 | Sn | mg/m$^3$ | HK | Coal & solid waste | Coal & solid waste | Stack | 3 | 0.0128 | NA | MP 1 |
| Ubaque 2010 | Sn | mg/m$^3$ | HK | Coal & solid waste | Coal & solid waste | Stack | 3 | 0.0146 | NA | MP 2 |
| Ubaque 2010 | Sn | mg/m$^3$ | HK | Coal & solid waste | Coal & solid waste | Stack | 3 | 0.0117 | NA | MP 3 |
| Ubaque 2010 | Sn | mg/m$^3$ | HK | Coal & solid waste | Coal & solid waste | Stack | 3 | 0.0081 | NA | MP 4 |
| Ubaque 2010 | Sn | mg/m$^3$ | HK | Coal & solid waste | Coal & solid waste | Stack | 3 | 0.0214 | NA | MP 5 |
| Ubaque 2010 | Sn | mg/m$^3$ | HK | Coal & solid waste | Coal & solid waste | Stack | 3 | 0.0106 | NA | MP 6 |
| Ubaque 2010 | Mn | mg/m$^3$ | HK | Coal & solid waste | Coal & solid waste | Stack | 3 | 0.0091 | NA | MP 1 |
| Ubaque 2010 | Mn | mg/m$^3$ | HK | Coal & solid waste | Coal & solid waste | Stack | 3 | 0.0168 | NA | MP 2 |
| Ubaque 2010 | Mn | mg/m$^3$ | HK | Coal & solid waste | Coal & solid waste | Stack | 3 | 0.0167 | NA | MP 3 |
| Ubaque 2010 | Mn | mg/m$^3$ | HK | Coal & solid waste | Coal & solid waste | Stack | 3 | 0.0146 | NA | MP 4 |
| Ubaque 2010 | Mn | mg/m$^3$ | HK | Coal & solid waste | Coal & solid waste | Stack | 3 | 0.0107 | NA | MP 5 |
| Ubaque 2010 | Mn | mg/m$^3$ | HK | Coal & solid waste | Coal & solid waste | Stack | 3 | 0.0584 | NA | MP 6 |
| Ubaque 2010 | Ni | mg/m$^3$ | HK | Coal & solid waste | Coal & solid waste | Stack | 3 | 0.0021 | NA | MP 1 |

| | | | | | | | | | | |
|---|---|---|---|---|---|---|---|---|---|---|
| Ubaque 2010 | Ni | $mg/m^3$ | HK | Coal & solid waste | Coal & solid waste | Stack | 3 | 0.0116 | NA | MP 2 |
| Ubaque 2010 | Ni | $mg/m^3$ | HK | Coal & solid waste | Coal & solid waste | Stack | 3 | 0.0217 | NA | MP 3 |
| Ubaque 2010 | Ni | $mg/m^3$ | HK | Coal & solid waste | Coal & solid waste | Stack | 3 | 0.0103 | NA | MP 4 |
| Ubaque 2010 | Ni | $mg/m^3$ | HK | Coal & solid waste | Coal & solid waste | Stack | 3 | 0.0079 | NA | MP 5 |
| Ubaque 2010 | Ni | $mg/m^3$ | HK | Coal & solid waste | Coal & solid waste | Stack | 3 | 0.0015 | NA | MP 6 |
| Ubaque 2010 | Pb | $mg/m^3$ | HK | Coal & solid waste | Coal & solid waste | Stack | 3 | 0.005 | NA | MP 1 |
| Ubaque 2010 | Pb | $mg/m^3$ | HK | Coal & solid waste | Coal & solid waste | Stack | 3 | 0.007 | NA | MP 2 |
| Ubaque 2010 | Pb | $mg/m^3$ | HK | Coal & solid waste | Coal & solid waste | Stack | 3 | 0.005 | NA | MP 3 |
| Ubaque 2010 | Pb | $mg/m^3$ | HK | Coal & solid waste | Coal & solid waste | Stack | 3 | 0.002 | NA | MP 4 |
| Ubaque 2010 | Pb | $mg/m^3$ | HK | Coal & solid waste | Coal & solid waste | Stack | 3 | 0.005 | NA | MP 5 |
| Ubaque 2010 | Pb | $mg/m^3$ | HK | Coal & solid waste | Coal & solid waste | Stack | 3 | 0.003 | NA | MP 6 |
| Ubaque 2010 | Hg | $mg/m^3$ | HK | Coal & solid waste | Coal & solid waste | Stack | 3 | <0.0002 | NA | MP 1 |
| Ubaque 2010 | Hg | $mg/m^3$ | HK | Coal & solid waste | Coal & solid waste | Stack | 3 | <0.0002 | NA | MP 2 |
| Ubaque 2010 | Hg | $mg/m^3$ | HK | Coal & solid waste | Coal & solid waste | Stack | 3 | <0.0002 | NA | MP 3 |

| | | | | | | | | | | |
|---|---|---|---|---|---|---|---|---|---|---|
| Ubaque 2010 | Hg | mg/m$^3$ | HK | Coal & solid waste | Coal & solid waste | Stack | 3 | <0.0002 | NA | MP 4 |
| Ubaque 2010 | Hg | mg/m$^3$ | HK | Coal & solid waste | Coal & solid waste | Stack | 3 | <0.0002 | NA | MP 5 |
| Ubaque 2010 | Hg | mg/m$^3$ | HK | Coal & solid waste | Coal & solid waste | Stack | 3 | <0.0002 | NA | MP 6 |
| Vaidya 2015 | CO | ppm | NS | NS | NS | Site at brick kiln | 15 | 62.8 | 3.1 | summer |
| Vaidya 2015 | CO | ppm | NS | NS | NS | Site at brick kiln | 25 | 55.5 | 7.26 | winter |
| Vaidya 2015 | PM | mg/m$^3$ | NS | NS | NS | Site at brick kiln | 15 | 146.1 | 64.03 | summer |
| Vaidya 2015 | PM | mg/m$^3$ | NS | NS | NS | Site at brick kiln | 25 | 91.4 | 19.50 | winter |
| Zawilla 2014 | TSP | mg/m$^3$ | NS | NS | NS | Site at brick factory | 16 | 4.26 | 1.54 | Production area |
| Zawilla 2014 | TSP | mg/m$^3$ | NS | NS | NS | Site at brick factory | 16 | 9.97 | 1.87 | Mining area |

**Table S7. Summary of mean production-based emission factors (g/kg brick) stratified by kiln design and fuel type.**

| Pollutant | Kiln design | Fuel used | Number of studies | Number of samples | Mean ± SD | Min. | Max. |
|---|---|---|---|---|---|---|---|
| PM | Traditional | Coal | 2 | 2 | 0.46 ± 0.24 | 0.29 | 0.64 |
| | Improved | | 4 | 19 | 0.2 ± 0.15 | 0.09 | 0.31 |
| | NA | | 1 | 34 | 0.19 ± 0.27 | 0.19 | 0.19 |
| | Traditional | Coal & biomass | 1 | 15 | 0.89 ± 0.86 | 0.89 | 0.89 |
| | Improved | | 1 | 15 | 0.22 ± 0.2 | 0.22 | 0.22 |
| | Traditional | Biomass | 1 | 3 | 1.56 ± 1.4 | 1.56 | 1.56 |
| $PM_1$ | Traditional | Coal | 1 | 9 | 0.33 ± 0 | 0.33 | 0.33 |
| $PM_{2.5}$ | Traditional | Coal | 1 | 9 | 0.34 ± 0 | 0.34 | 0.34 |
| | Improved | | 12 | 36 | 0.17 ± 0.16 | 0.03 | 0.42 |
| | Traditional | Coal & biomass | 6 | 24 | 0.33 ± 0.23 | 0.08 | 1.06 |
| | Improved | | 2 | 4 | 0.14 ± 0 | 0.1 | 0.24 |
| | Traditional | Biomass | 2 | 4 | 0.53 ± 0.08 | 0.5 | 0.632 |
| | Improved | | 1 | 1 | 0.39 | 0.39 | 0.39 |
| $PM_{10}$ | Traditional | Coal | 1 | 9 | 0.34 ± 0.5 | 0.34 | 0.34 |
| | Traditional | Biomass | 1 | 1 | 0.74 | 0.74 | 0.74 |
| | Improved | | 1 | 1 | 0.45 | 0.45 | 0.45 |
| BC | Improved | Coal | 3 | 9 | 0.02 ± 0.01 | 0.01 | 0.04 |
| | Traditional | Coal & biomass | 3 | 15 | 0.03 ± 0.17 | 0.03 | 0.043 |
| | Improved | | 2 | 4 | 0.01 ± 0 | 0.01 | 0.02 |
| EC | Improved | Coal | 9 | 27 | 0.01 ± 0.008 | 0.001 | 0.03 |
| | Traditional | Coal & biomass | 3 | 9 | 0.16 ± 0.05 | 0.09 | 0.27 |
| | Traditional | Biomass | 2 | 4 | 0.16 ± 0.07 | 0.05 | 0.19 |
| | Improved | | 1 | 1 | 0.04 | 0.04 | 0.04 |

| | | | | | | | |
|---|---|---|---|---|---|---|---|
| OC | Traditional | Biomass | 1 | 1 | 0.35 | 0.35 | 0.35 |
| | Improved | | 1 | 1 | 0.29 | 0.29 | 0.29 |
| CO | Traditional | Coal | 3 | 11 | 7.18 ± 6.65 | 2.89 | 7.83 |
| | Improved | | 16 | 55 | 1.71 ± 0.87 | 0.31 | 4.5 |
| | Traditional | Coal & biomass | 6 | 35 | 2.26 ± 2.08 | 0.44 | 3.63 |
| | Improved | | 2 | 16 | 0.43 ± 0.37 | 0.35 | 1.62 |
| | Traditional | Biomass | 2 | 4 | 7.06 ± 0.2 | 5.01 | 13.2 |
| $CO_2$ | Traditional | Coal | 1 | 9 | 132 ± 79.7 | 132 | 132 |
| | Improved | | 7 | 28 | 81 ± 49.5 | 12.6 | 149 |
| | Traditional | Coal & biomass | 4 | 30 | 112 ± 73.8 | 14.9 | 179 |
| | Improved | | 3 | 19 | 112 ± 36.2 | 82 | 119 |
| | Traditional | Biomass | 1 | 3 | 526 ± 179 | 526 | 526 |
| NO | Traditional | Coal | 1 | 9 | 0.05 ± 0.03 | 0.05 | 0.05 |
| $NO_2$ | Traditional | Coal | 1 | 9 | 0 ± 0 | 0 | 0 |
| $NO_x$ | Traditional | Coal | 1 | 9 | 0.05 ± 0.03 | 0.05 | 0.05 |
| | NA | | 1 | 34 | 0.59 ± 0.8 | 0.59 | 0.59 |
| $SO_2$ | Traditional | Coal | 3 | 11 | 0.58 ± 0.25 | 0.24 | 2.68 |
| | Improved | | 7 | 28 | 0.66 ± 0.49 | 0.1 | 1.8 |
| | NA | | 1 | 34 | 5.48 ± 6.25 | 5.48 | 5.48 |
| | Traditional | Coal & biomass | 4 | 30 | 1.08 ± 1.09 | 0.52 | 1.9 |
| | Improved | | 3 | 19 | 0.21 ± 0.2 | 0.06 | 0.9 |
| | Traditional | Biomass | 1 | 3 | 0 ± 0 | 0 | 0 |
| VOCs | Improved | Coal | 2 | 8 | 82.7 ± 38.2 | 77.6 | 98 |
| | NA | | 1 | 34 | 0.27 ± 0.45 | 0.27 | 0.27 |
| | Traditional | Coal & biomass | 1 | 10 | 76.7 ± 47.2 | 76.7 | 76.7 |

**Table S8. Summary of mean fuel-based emission factors (g/kg fuel) stratified by kiln design and fuel type.**

| Pollutant | Kiln design | Fuel used | Number of studies | Number of samples | Mean ± SD | Min. | Max. |
|---|---|---|---|---|---|---|---|
| PM | Traditional | Coal | 1 | 1 | 42.2 | 42.22 | 42.22 |
| | NA | | 1 | 34 | 0.56 ± 1.13 | 0.56 | 0.56 |
| | Improved | Coal & biomass | 1 | 1 | 1.01 | 1.01 | 1.01 |
| $PM_1$ | Traditional | Coal & biomass | 1 | 1 | 1.76 | 1.759 | 1.759 |
| | Improved | | 1 | 1 | 1.82 | 1.823 | 1.823 |
| $PM_{2.5}$ | Traditional | Coal | 1 | 1 | 5.4 | 5.4 | 5.4 |
| | Improved | | 12 | 36 | 2.51 ± 1.54 | 0.5 | 5.88 |
| | Traditional | Coal & biomass | 8 | 26 | 5.3 ± 3.22 | 1.7 | 13.41 |
| | Improved | | 4 | 6 | 9.1 ± 1 | 3.1 | 19.06 |
| | Traditional | Biomass | 7 | 9 | 2.41 ± 0 | 1.24 | 4.62 |
| | Improved | | 2 | 2 | 1.8 ± 0.2 | 1.66 | 1.94 |
| BC | Traditional | Coal | 1 | 1 | 0.37 | 0.37 | 0.37 |
| | Improved | | 3 | 9 | 0.27 ± 0.09 | 0.06 | 0.31 |
| | Traditional | Coal & biomass | 4 | 16 | 0.42 ± 0.18 | 0.014 | 0.6 |
| | Improved | | 3 | 5 | 0.36 ± 0.2 | 0.112 | 0.466 |
| | Traditional | Biomass | 4 | 4 | 0.64 ± 0.32 | 0.28 | 1.03 |
| | Improved | | 2 | 2 | 0.41 ± 0.37 | 0.15 | 0.67 |
| EC | Improved | Coal | 9 | 27 | 0.17 ± 0 | 0.01 | 0.5 |
| | Traditional | Coal & biomass | 6 | 12 | 2.06 ± 0 | 0.014 | 3.7 |
| | Improved | | 3 | 3 | 0.007 ± 0 | 0.007 | 0.007 |
| | Traditional | Biomass | 1 | 3 | 1.1 ± 0 | 1.1 | 1.1 |
| OC | Traditional | Coal & biomass | 3 | 3 | 6.77 ± 1.89 | 4.68 | 8.37 |
| | Improved | | 3 | 3 | 1 ± 0.33 | 0.63 | 1.26 |
| | Traditional | Biomass | 4 | 4 | 0.45 ± 0.49 | 0.14 | 1.18 |
| | Improved | | 2 | 2 | 0.28 ± 0.35 | 0.03 | 0.52 |

| | | | | | | | |
|---|---|---|---|---|---|---|---|
| CO | Traditional | Coal | 2 | 2 | 96 ± 102 | 24 | 168 |
| | Improved | | 12 | 36 | 20.6 ± 2.77 | 5.6 | 67.4 |
| | Traditional | Coal & biomass | 5 | 20 | 24.8 ± 5.27 | 7.5 | 70.9 |
| | Improved | | 2 | 2 | 19.0 ± 12.6 | 10.1 | 27.97 |
| | Traditional | Biomass | 8 | 8 | 64.6 ± 30.4 | 25.7 | 105.3 |
| | Improved | | 2 | 2 | 54.9 ± 14.8 | 44.4 | 65.4 |
| $CO_2$ | Traditional | Coal | 1 | 1 | 1630 | 1630 | 1630 |
| | Improved | | 3 | 9 | 462 ± 147 | 237 | 1410 |
| | Traditional | Coal & biomass | 3 | 15 | 737 ± 104 | 242 | 2102 |
| | Improved | | 3 | 5 | 2280 ± 232 | 1981 | 2836 |
| | Traditional | Biomass | 7 | 7 | 1679 ± 96.2 | 1527 | 1787 |
| | Improved | | 2 | 2 | 1589 ± 8.49 | 1583 | 1595 |
| NO | Traditional | Coal & biomass | 1 | 1 | 0 | 0 | 0 |
| | Improved | | 1 | 1 | 1.28 | 1.28 | 1.28 |
| | Traditional | Biomass | 3 | 3 | 0.86 ± 0.17 | 0.76 | 1.05 |
| | Improved | | 2 | 2 | 1.02 ± 0 | 1.02 | 1.02 |
| $NO_2$ | Traditional | Coal & biomass | 1 | 1 | 0.3 | 0.297 | 0.297 |
| | Improved | | 1 | 1 | 0.08 | 0.08 | 0.08 |
| | Traditional | Biomass | 3 | 3 | 0.98 ± 0.05 | 0.93 | 1.01 |
| | Improved | | 2 | 2 | 1.7 ± 0 | 1.7 | 1.7 |
| $NO_x$ | Traditional | Coal | 1 | 1 | 1.42 | 1.42 | 1.42 |
| | NA | | 1 | 34 | 1.09 ± 0.9 | 1.09 | 1.09 |
| | Improved | Coal & biomass | 1 | 1 | 1.8 | 1.8 | 1.8 |
| $SO_2$ | Traditional | Coal | 2 | 2 | 114 ± 141 | 14.2 | 213 |
| | Improved | | 3 | 9 | 19.9 ± 13.7 | 1.14 | 33.7 |
| | NA | | 1 | 34 | 7.17 ± 7.49 | 7.17 | 7.17 |
| | Traditional | Coal & biomass | 3 | 15 | 24.5 ± 18.8 | 13 | 26.7 |
| | Improved | | 3 | 5 | 18.5 ± 22 | 7.97 | 24 |

| | | | | | | | |
|---|---|---|---|---|---|---|---|
| | Traditional | Biomass | 3 | 3 | 0.18 ± 0.08 | 0.13 | 0.27 |
| | Improved | | 2 | 2 | 1 ± 0 | 1 | 1 |
| VOCs | Improved | Coal | 2 | 8 | 1516 ± 623 | 1371 | 1953 |
| | NA | | 1 | 34 | 0.52 ± 0.92 | 0.52 | 0.52 |
| | Traditional | Coal & biomass | 1 | 10 | 1085 ± 947 | 1085 | 1085 |

**Table S9. Summary of mean energy-based emission factors (g/MJ) stratified by kiln design and fuel type.**

| Pollutant | Kiln design | Fuel used | Number of studies | Number of samples | Mean ± SD | Min. | Max. |
|---|---|---|---|---|---|---|---|
| PM | Traditional | Coal | 1 | 1 | 1.83 | 1.83 | 1.83 |
| | Improved | | 4 | 19 | 0.2 ± 0.14 | 0.1 | 0.23 |
| | Traditional | Coal & biomass | 1 | 15 | 0.66 ± 0.59 | 0.66 | 0.66 |
| | Improved | | 2 | 16 | 0.2 ± 0.19 | 0.038 | 0.21 |
| | Traditional | Biomass | 1 | 3 | 0.54 ± 0.49 | 0.54 | 0.54 |
| $PM_{2.5}$ | Improved | Coal | 2 | 8 | 0.24 ± 0.17 | 0.21 | 0.25 |
| | Traditional | Coal & biomass | 1 | 10 | 0.27 ± 0.21 | 0.27 | 0.27 |
| BC | Improved | Coal | 2 | 8 | 0.01 ± 0.004 | 0.011 | 0.014 |
| | Traditional | Coal & biomass | 1 | 10 | 0.02 ± 0.01 | 0.02 | 0.02 |
| CO | Traditional | Coal | 1 | 1 | 7.24 | 7.24 | 7.24 |
| | Improved | | 6 | 27 | 1.89 ± 1.14 | 0.24 | 4.39 |
| | Traditional | Coal & biomass | 2 | 25 | 1.91 ± 2.11 | 0.33 | 2.96 |
| | Improved | | 2 | 16 | 0.36 ± 0.31 | 0.32 | 1.04 |
| | Traditional | Biomass | 1 | 3 | 5.17 ± 0.21 | 5.17 | 5.17 |
| $CO_2$ | Improved | Coal | 6 | 27 | 82.2 ± 48.2 | 10.9 | 146 |
| | Traditional | Coal & biomass | 2 | 25 | 88.3 ± 62.3 | 10.7 | 140 |
| | Improved | | 2 | 16 | 113 ± 33.9 | 106 | 113 |
| | Traditional | Biomass | 1 | 3 | 181 ± 61.5 | 181 | 181 |
| $NO_x$ | Traditional | Coal | 1 | 1 | 0.06 | 0.06 | 0.06 |
| | Improved | Coal & biomass | 1 | 1 | 0.07 | 0.07 | 0.07 |
| $SO_2$ | Traditional | Coal | 1 | 1 | 9.2 | 9.2 | 9.2 |
| | Improved | | 6 | 27 | 0.63 ± 0.37 | 0.11 | 1.78 |

| | | | | | | | |
|---|---|---|---|---|---|---|---|
| | Traditional<br>Improved | Coal & biomass | 2<br>2 | 25<br>16 | 0.71 ± 0.57<br>0.08 ± 0.09 | 0.39<br>0.06 | 1.2<br>0.298 |
| | Traditional | Biomass | 1 | 3 | 0.07 ± 0.003 | 0.07 | 0.07 |
| VOCs | Improved | Coal | 2 | 8 | 67.8 ± 27.3 | 61.2 | 87.4 |
| | Traditional | Coal & biomass | 1 | 10 | 48.4 ± 41.6 | 48.4 | 48.4 |

Table S10. Summary of exposure biomarker concentrations.

| Exposure biomarker | | Exposure group | | | | | | | | | |
|---|---|---|---|---|---|---|---|---|---|---|---|
| ***1-OHP in urine (µmol/mol cr)*** | | ***Non-exposed*** | ***Low/moderate traffic*** | ***Heavy traffic*** | ***Waste landfill*** | ***Biomass combustion*** | ***Metallurgy*** | ***Brick kilns*** | | | |
| Martinez-Salinas 2010[46] | Geometric means | – | 0.08 ± 0.20 (n=39) | 0.20 ± 0.20 (n=17) | 0.30 ± 0.50 (n=32) | 3.25 ± 0.35 (n=105) | – | 0.35 ± 0.21 (n=65) | | | |
| Perez-Maldonado 2019[50] | Geometric means | – | 0.05 ± 0.04 (n=20) | 0.09 ± 0.05 (n=25) | 0.10 ± 0.08 (n=30) | 3.10 ± 1.25 (n=30) | – | 0.50 ± 0.15 (n=30) | | | |
| Flores-Ramirez 2018[33] | Medians | – | – | 0.06 (n=29) | 0.09 (n=19) | – | 0.03 (n=27) | 0.23 (n=40) | | | |
| Kamal 2014b[105] | Medians | 0.62 (n=34) | – | – | – | – | – | 1.53 (n=46) | | | |
| ***t,t-MA in urine (µg/g cr)*** | | | | | | | | | | | |
| Flores-Ramirez 2018[33] | Medians | – | – | 220.6 (n=29) | 427.4 (n=19) | – | 258.6 (n=27) | 429.7 (n=40) | | | |
| ***Fluoride in urine (mg/L)*** | | | | | | | | | | | |
| Flores-Ramirez 2018[33] | Medians | – | – | 2.3 (n=29) | 1.2 (n=19) | – | 1.7 (n=27) | 1.5 (n=40) | | | |
| Khanoranga 2019[43] | Means | 0.003 ± 0.002 (n=20) | – | – | – | – | – | 0.21 ± 0.19 (n=100) | | | |
| ***Heavy metals in blood*** | | | ***Controls*** | ***Nonworkers near kilns*** | ***Children at kiln sites*** | ***Brick workers*** | | | ***Workers in other industries*** | | |
| | | | | | | ***Brick makers*** | ***Brick carriers*** | ***Brick bakers*** | ***Furniture*** | ***Paint*** | ***Welding*** |
| ***Pb (µg/dl)*** | | | | | | | | | | | |
| Jahan 2016[102] | Means | | 1.28 ± 0.12 (n=57) | 2.90 ± 0.15 (n=30) | – | 5.10 ± 0.35 (n=45) | 5.19 ± 0.34 (n=30) | 5.63 ± 0.83 (n=35) | – | – | – |
| Flores-Ramirez 2018[33] | Medians | | – | 4.2 (n=40) | – | – | | | – | – | – |

| | | | | | | | | | | | |
|---|---|---|---|---|---|---|---|---|---|---|---|
| Berumen-Rodriguez 2021[100] | Medians | | – | – | – | 1.08<br>(n=42) | | | – | – | – |
| ***Cd (µg/dl)*** | | | | | | | | | | | |
| Jahan 2016[102] | Means | | 1.23 ± 0.12<br>(n=57) | 2.65 ± 0.15<br>(n=30) | – | 3.69 ± 0.18<br>(n=45) | 3.58 ± 0.32<br>(n=30) | 3.58 ± 0.25<br>(n=35) | – | – | – |
| David 2020[74] | Means | | 1.19 ± 0.01<br>(n=114) | – | | 3.09 ± 0.01<br>(n=118) | | | – | – | – |
| David 2021[101] | Means | | 1.40 ± 0.22<br>(n=98) | – | 2.16 ± 0.15<br>(n=134) | – | – | – | – | – | – |
| David 2022[9] | Means | | 2.37 ± 0.01<br>(n=200) | – | – | 5.14 ± 0.02<br>(n=346) | | | | | |
| ***Cr (µg/dl)*** | | | | | | | | | | | |
| Jahan 2016[102] | Means | | 1.83 ± 0.09<br>(n=57) | 2.01 ± 0.09<br>(n=30) | – | 2.55 ± 0.06<br>(n=45) | 2.46 ± 0.10<br>(n=30) | 2.73 ± 0.07<br>(n=35) | – | – | – |
| David 2020[74] | Means | | 2.81 ± 0.02<br>(n=114) | – | | 4.20 ± 0.02<br>(n=118) | | | – | – | – |
| David 2021[101] | Means | | 0.87 ± 0.05<br>(n=98) | – | 1.09 ± 0.05<br>(n=134) | – | – | – | – | – | – |
| David 2022[9] | Means | | 2.02 ± 0.01<br>(n=200) | – | – | 5.27 ± 0.02<br>(n=346) | | | – | – | – |
| ***Zn (µg/dl)*** | | | | | | | | | | | |
| David 2021[101] | Means | | 17.97 ± 1.99<br>(n=98) | – | 26.63 ± 2.21<br>(n=134) | – | – | – | – | – | – |
| David 2022[9] | Means | | 1.24 ± 0.01<br>(n=200) | – | – | 1.10 ± 0.01<br>(n=346) | | | – | – | – |
| ***Ni (µg/dl)*** | | | | | | | | | | | |
| David 2020[74] | Means | | 3.24 ± 0.02<br>(n=114) | – | | 5.59 ± 0.03<br>(n=118) | | | – | – | – |
| David 2021[101] | Means | | 0.54 ± 0.07<br>(n=98) | – | 0.96 ± 0.15<br>(n=134) | – | – | – | – | – | – |
| David 2022[9] | Means | | 3.88 ± 0.01<br>(n=200) | – | – | 6.40 ± 0.02<br>(n=346) | | | – | – | – |
| ***As (µg/dl)*** | | | | | | | | | | | |
| Ahmad 2020[97] | Means | | 0.97<br>(n=250) | – | – | 3.24<br>(n=65) | | | 2.53<br>(n=60) | 2.39<br>(n=60) | 2.32<br>(n=65) |
| Akram 2022[98] | Means | | 0.97 ± 0.1<br>(n=300) | | | 3.24 ± 0.34<br>(n=60) | | | 2.53 ± 0.15<br>(n=60) | 2.39 ± 0.12<br>(n=60) | 2.32 ± 0.14<br>(n=60) |

### Table S11. Characteristics of included studies with health data.

| Study | Country | Dates | Methods | Type of study | Population | Ages | Sex | Sample size | Health data collected |
|---|---|---|---|---|---|---|---|---|---|
| Ahmad 2020[97] | Pakistan | Not reported | The authors sought to study the genetic and expression variations of PSMD1 gene as a consequence of arsenic exposure and its potential implications in arsenic induced diseases. 250 blood samples of exposed industrial workers along with 250 controls were used. Tetra amplification refractory mutation system-PCR was used to determine the role of PSMD1 gene polymorphisms (rs1549339, rs13402242) in industrial workers and controls. Hair and nail pieces were also collected. Mean concentration of inorganic arsenic was calculated in blood, hair, and nail samples. No explicit information on sampling or eligibility criteria other than work groups. Number of brick kilns not reported. Authors evaluated outcomes in participants with >10 and <10 years of exposure. | Cross-sectional | 250 individuals with occupational exposure to arsenic (65 from brick kilns, 65 from welding, 60 from furniture and 60 from paint) along with age and gender matched 250 healthy individuals | Mean age (± SD) was 26.9 ± 19.7 years; for BKWs it was 30.4 ± 17.3 years. | Sex was recorded but not reported. | 500 | Genetic and expression variations of PSMD1 gene |
| Akram 2022[98] | Pakistan | Not reported | Blood samples were obtained from 300 occupational workers (60/industry) involved in welding, brick kiln, furniture, pesticide, and the paint industry with 5+ years of work experience, along with age and gender matched 300 unexposed control subjects. Total arsenic content was measured in blood samples of control and industrial workers using the wet acid digestion method. Eligibility criteria other than occupational work group were not specified. Number of brick kilns not reported. Authors evaluated outcomes in participants with >10, >25, <10, and <25 years of exposure. | Cross-sectional | 300 occupational workers (60/industry) involved in welding, brick kiln, furniture, pesticide, and the paint industry with 5+ years of work experience, along with age and gender matched 300 unexposed | Mean age (± SD) was 27.9 ± 21.1 years, for BKWs it was 29.3 ± 13.1 years. | Sex was collected but not reported. | 600 | Variation in HPRT, OGG1 gene expression, and DNA damage in blood cells by comet assay. |

| | | | | | | | | | |
|---|---|---|---|---|---|---|---|---|---|
| | | | | | control subjects | | | | |
| Ali 2013[69] | Pakistan | Apr 2011 | Questionnaires were used to gather health data. Children with tuberculosis, asthma, mental disability or malnutrition were excluded. Number of brick kilns not reported. | Cross-sectional | Children living in two villages near and far away from a brick kiln | < 5 years of age | Boys and girls | 188 (sex distribution not specified) | Acute respiratory infections in the last two weeks |
| Berumen-Rodriguez 2021[100] | Mexico | Nov 2019 (confirmed with corresponding author) | Spirometry tests (pre-and post-bronchodilator) were performed on participants who met the inclusion criteria, following 2005 ATS/ERS guidelines using the EasyOne® Plus Diagnostic portable spirometer. The normal values predicted were those established for the Mexican-American population in the NHANES III study. A Spanish-validated questionnaire was used to detect the risk of COPD (COPD-PS). Eligibility criteria were: working in a brick-kiln in the area; voluntary, signed, informed consent; and subjects with absence of chronic cough/sputum or dyspnea and without recent surgeries. Multiple brick kilns. The authors evaluated the relationship between COPD-PS questionnaire and number of years worked. | Cross-sectional | Adult brick kiln workers | Mean age (± SD) was 54.9 ± 16 years (confirmed with corresponding author) | Males | 41 (all males, confirmed with corresponding author) | Spirometry (post $FEV_1$, Post $FEV_1$%, Post PEF);; Risk of COPD by questionnaire; data on body mass index; prevalence of self-reported hypertension and diabetes |
| Berumen-Rodriguez 2023[99] | Mexico | Jan 2021 (confirmed with corresponding author) | A Spanish-validated questionnaire was used to detect the risk of COPD (COPD-PS). Research team collected urine and exhaled breath condensate for biomarkers and cytokine analysis. Participation was voluntary and all participants signed an informed consent, but eligibility criteria were not specified. Multiple brick kilns. | Cross-sectional | Adult brick kiln workers | Median age (range) was 57 (23–79) years | Males | 21 (all male, confirmed with corresponding author) | Risk of COPD by questionnaire; Cytokines in exhaled breath condensate |

| | | | | | | | | | |
|---|---|---|---|---|---|---|---|---|---|
| Biswas 2018[70] | Bangladesh | Apr –May 2018 | The Standardized Nordic questionnaire was used to collection information of musculoskeletal symptoms and pain. The American Thoracic Society Division of Lung Disease questionnaire was used to gather information on respiratory illness and symptoms. Workers with osteoarthritis, rheumatoid arthritis, and cervical spondylitis were excluded. Multiple brick kilns. The authors evaluated respiratory illness/musculoskeletal pain with the number of years worked (<5, 5-10, >10 years). | Cross-sectional | Adult brick kiln workers | Mean age (± SD) was 39 ± 13.2 years | 74.5% males and 25.5% females | 220 | Musculoskeletal pain/illness; Cough; Chronic cough; Phlegm; Chronic phlegm; Wheeze; Asthma; Bronchitis |
| Das 2014[71] | India | Mar 2012 – Apr 2013 | A modified Nordic musculoskeletal disorder questionnaire was used to collection information of musculoskeletal symptoms and pain. Hand-grip strength was measured with a dynamometer. Spirometry was conducted using an RMS HELIOS 401. Three recordings were attempted and the best of 3 was used. Peak expiratory flow was assessed using a Wright's peak flow meter. Heart rate was also collected. Eligibility criteria were age 18-58 years, experience in brick kiln activities or office activities for 1 year. Multiple brick kilns. The authors obtained information on the number of years worked but did not evaluate relationships between health outcomes and number of years worked. | Cross-sectional | Adult male brick kiln workers from 1 brick kiln in the Hooghly district, and male office workers (not involved in hand-intensive jobs). | Mean age (± SD) of male brick kiln workers was 33.5 ± 6.2 years; and of office workers was 34.2 ± 6.7 years | 100% males | 220 brick kiln workers and 130 office workers | Musculoskeletal pain/illness; Hand-grip strength; heart rate; $FEV_1$, FVC, $FEV_1$/FVC, Peak expiratory flow |
| Das 2019a[73] | India | Not reported | A modified Nordic musculoskeletal disorder questionnaire was used to collection information of musculoskeletal symptoms and pain. Hand-grip strength was measured with a dynamometer. Spirometry was conducted using an RMS HELIOS 401. Three recordings were attempted and the best of 3 was used. Peak expiratory flow was assessed using a | Cross-sectional | Male and female children who worked at 28 selected brick kilns or who were not involved in manual labor at the kilns | Mean age of child brick kiln workers and controls was 12.8 and 13.4 years, respectively | Males and females (sex distribution incompletely specified) | 112 brick kiln workers (66 female, 45 male) and 120 controls (sex distribution not specified) | Musculoskeletal pain/illness; Gastrointestinal problems; Cardiovascular problems; Respiratory problems (overall); Skin diseases; |

| | | | | | | | | | |
|---|---|---|---|---|---|---|---|---|---|
| | | | mini-Wright's peak flow meter. Heart rate was measured with the 10-beats method and blood pressure was measured using a sphygmomanometer and stethoscope. Eligibility criteria include children aged 9-16 years working in the kiln for at least 1 year. Controls were not involved in manual labor at the kiln, instead they did different types of household activities like serving tea, water and food. Multiple brick kilns. The authors obtained information on the number of years worked but did not evaluate relationships between health outcomes and number of years worked. | | | | | | Headache; Eye irritation; Hand-grip strength; heart rate; blood pressure (resting and just after work), $FEV_1$, FVC, $FEV_1$/FVC, Peak expiratory flow |
| Das 2019b[72] | India | Not reported | This study sought to evaluate sex differences in musculoskeletal disorders and physiological stress in BKWs. The study enrolled BKWs at 12 kilns. No other eligibility criteria were specified. A modified Nordic Questionnaire was completed by male and female brickfield workers. Physiology before/after work and lung function was also measured. Multiple brick kilns. The authors obtained information on the number of years worked but did not evaluate relationships between health outcomes and number of years worked. The study stratified by type of BKW (brick molder and carrier). | Cross-sectional | Males and female BKWs | Mean age (± SD) was 31.9 ± 8.8 years | 152 males (51%) and 148 females (49%) | 300 | Musculoskeletal disorders; physiological parameters (HR and blood pressure) before and after work; $FEV_1$, FVC, $FEV_1$/FVC, PEF. |
| David 2020[74] | Pakistan | Mar 2018 – Mar 2019 | The goal of the study was to compare reproductive parameters, heavy metal exposures, hematology, biochemistry and reproductive hormone concentrations. A blood sample was obtained from all participants. Eligibility criteria included adult females aged 18-55 years working at brick kilns and adult female aged 18-58 years who lived in the same district. Multiple brick kilns. Number of years | Cross-sectional | Adult female BKW and unexposed controls | Mean age (± SD) was 31.8 ± 17.4 years for the study population; 35.8 ± 19.6 for BKWs and 27.7 ± 13.9 for non-BKWs | 100% females | 232 (118 BKW and 114 controls) | Average age of menstrual onset; average number of dead children/abortion rate; cadmium, nickel and chromium levels; hematological and biochemical parameters |

| | | | | | | | | | |
|---|---|---|---|---|---|---|---|---|---|
| | | | worked collected but authors did not evaluate relationships between health outcomes and number of years worked. | | | | | | (SOD, guiacol peroxidase activity, ROS, thiobarbituric acid reactive species); lipid profile; hormone concentrations |
| David 2021[101] | Pakistan | Jun 2018 – Jun 2019 | The study sought to evaluate the environmental effects of heavy metals on biochemical profile and oxidative stress among children at brick kiln sites. Eligibility criteria include children ages 4-17 years working and living at brick kiln sites (convenience sample) and children who lived far away from the kilns but lived in the same district. Blood samples were collected and analyzed for complete blood count, heavy metal exposure, biochemistry and hormonal analysis. Multiple brick kilns. | Cross-sectional | Child BKWs and non-BKW controls | Mean age (± SD) was 11.2 ± 11.4 years; of BKWs was 9.9 ± 9.8 years; and, of non-BKWs was 13.0 ± 1.3 years | Sex distribution not reported | 232 children (134 BKWs and 98 non-BKWs) | Reactive oxidative species, Catalase activity, SOD, guiacol peroxidase activity, thiobarbituric acid reactive species growth hormone and cortisol, hematological parameters and DNA damage |
| David 2022[9] | Pakistan | Mar – Nov 2018 | Structured questionnaire to ask about self-reported illnesses and symptoms, and a blood sample to measure biochemical and hematological parameters, an antioxidant enzyme, and reproductive hormone parameters. Multiple brick kilns. Authors collected data on number of years living in kilns and number of years working in kilns, and analyzed the correlation between health outcomes and the number of years living in kilns. | Cross-sectional | Adult BKWs and adult non-BKWs who lived in the same district, but at least 40 km away from the kilns. Control participants' least exposed to environmental pollutants such as vehicle and industrial smoke were selected. | Mean age (± SD) of BKWs was 23.0 ± 4.3 years; of non-BKWs (controls) was 26.9 ± 10.9 years | 100% males | 546 participants: 346 brick kiln workers and 200 controls | Health history including asthma, allergies, diabetes, obesity, stomach issues, hyperandrogenism, kidney issues, tuberculosis, hepatitis, and use of medications; cadmium, nickel, chromium in BKWs; hemoglobin and other hematological parameters, SOD, ROS, FSH, LH, cortisol, and testosterone. |

| | | | | | | | | | |
|---|---|---|---|---|---|---|---|---|---|
| Erdim 2020[75] | Turkey | Sep 2016 – Mar 2017 | The goal of the study was to evaluate BKWs in the field using otorhinolaryngologic and head and neck surgery (ENT) assessments. Evaluations occurred before doing to the brick kiln in a subset of 29 participants, at the brick kiln in a subset of 63 participants, and after working at the brick kiln in 11 participants. The study involved a questionnaire and an ENT examination. Seventeen BKWs who completed the questionnaire (including mask use and number of years worked) did not participate in the examination. The authors report the average worker duration in the kiln but do not consider this in analysis. The study stratified by BKW type (those involved with the brick preparation before, during, and after they are baked in the kiln). | Cross-sectional | BKWs (unclear if children < 18 years were included). | Mean (± SD) age was 39.0 ± 9.7 years. | 99 (96%) male and 4 (4%) female | 103 | Otologic and structural nasal/rhinologic complication rates. |
| Goel 2015[76] | India | Not reported | Spirometry measured via computerized Spiro-exel Medicaid system, Chandigarh. Best of three readings considered for analysis. Chest x-ray (PA-view, 35x35cm) read by experienced radiologist. Eligibility criteria are non-smoker (never smoked), smokers who are not-brick kiln workers (daily tobacco use for more than 10 years); and smokers who are brick kiln workers (daily tobacco use for more than years and brick kiln worker for more than 10 years). Number of brick kilns not reported. The authors limited their enrollment to BKWs who worked more than 10 years. | Cross-sectional | Adult males who worked at a brick kiln (did not report on how they were selected), and non-smokers were randomly selected from the general population | Mean age (± SD) of non-brick kiln workers and non-smoker was 36 ± 4.8 years; of non-brick kiln workers who are smokers was 35 ± 14.5 years; and brick kiln workers who are smokers was 40 ± 7.9 years. | 100% males | 25 non-smokers, 50 non-brick kiln workers smokers, and 50 brick kiln workers who are smokers. | Chest x-ray findings; $FEV_1$, FVC, $FEV_1$/FVC, Peak expiratory flow, $FEF_{25-75\%}$ |
| Gonzalez 2021[77] | Colombia | Nov 2017 – Jul 2018 (confirmed with | This study sought to characterize respiratory symptoms in BKWs in Colombia. A validated questionnaires asking about respiratory symptom was used. Multiple brick kilns. The authors | Cross-sectional | Adult brick kiln workers | Mean age (range) was 38 (18-73) years | 81.1% males | 82 | Wheezing and chest tightness, shortness of breath whether at rest, during |

| | | | | | | | | | |
|---|---|---|---|---|---|---|---|---|---|
| | | corresponding author) | stratified by BKW type (baker, unloader, burner, or other). | | | | | | exercise or at night. Cough and phlegm (however, prevalence of phlegm not reported), whether daytime and/or night-time. Breathing, whether there are problems with breathing and how often they occur. Asthma: history and use of controller medications; nasal allergies, and insect sting allergy. |
| Gupta 2019[78] | India | Not reported | Symptom and illness history questionnaire collected using the American Thoracic Society Division of Lung Disease questionnaire. Eligibility criteria included age greater than 18 years and worked in kilns for at least 6 months. Multiple brick kilns (number of kilns not reported). Authors collected information of number of years worked (<3, 3-5, >5 years) but did not conduct an analysis linking health outcomes to the number of years worked. | Cross-sectional | Adult brick kiln workers in a rural area of the Jammu district | Mean age and SD are not reported (authors reported the number of people in 5 age categories) | 407 males and 285 females | 692 | Respiratory symptoms include chronic cough, phlegm, dyspnea, and chronic bronchitis. |
| Hamid 2023[96] | Pakistan | Aug 2020, Jan 2021 | Questionnaire about health issues. No eligibility criteria specified. Multiple brick kilns (number of kilns not reported). Authors did not collect information on the number of years worked. | Cross-sectional | Adult and child brick kiln workers in Sheikhupura and Pattoki, Pakistan | Mean age not provided. 76% were above 30 years; 12% were aged 21-30 years; 13% were aged 11-20 years; 42 children were under the age of 14 years. | 200 males, 180 females, 60 children | 440 | Dry cough; chest tightness; sore throat; common cold; itching/redness; rashes above elbows/knees/face; scaly patches/dandruff; chronic back pain; elbow/wrist pain/neck/should |

| | | | | | | | | | |
|---|---|---|---|---|---|---|---|---|---|
| | | | | | | | | | er pain; eye inflammation/irritation; weary eyes. |
| Jahan 2016[102] | Pakistan | Not reported | The authors sought to study antioxidant enzymes status and reproductive health of adult male workers exposed to brick kiln pollutants. Authors did not collect data on the number of years worked. The study stratified by BKW type (brick maker, carriers, and bakers). | Cross-sectional | Adult BKWs and non-BKWs (two types, individuals living close to BKWs and other who were considered unexposed) | Mean (± SD) age was 27.3 ± 12.1 years; for BKWs was 27.9 ± 14.7 years; for non-BKWs living nearby was 27 ± 8.5 years; and for controls 25.9 ± 5 years. | 100% males | 197 (110 BKWs, 30 non-BKWs living in same vicinity and 57 unexposed adult males) | Hematological parameters; blood lead, cadmium and chromium levels; catalase activity, peroxidase activity, glutathione reductase activity, reduced glutathione, lipid peroxidation assay (malodialadehyde. thiobarbituric acid reactive substance; leutinizing hormone, testosterone. |
| Joshi 2008[103] | Nepal | Jun 2004 – Sep 2005 | Structured questionnaires and physical examination to evaluate for pallor, wax in ears, nasal abnormalities, abdominal distention, enlargement of cervical nodes, tonsillitis (enlarged tonsils). Eligibility criteria was all students under fifth grade who were present in the schools during the days of physical health examinations. Multiple brick kilns. | Cross-sectional | Children at a school close to brick kilns and children at a school far away from brick kiln | Mean age (± SD) of children at a school nearby brick kilns was 8.3 ± 2.3 years; of children at a school far from brick kilns was 7.8 ± 2.3 years (these values are from a subsample of 141 children who underwent | Not specified | 200 children at school near brick kilns, 120 at school far away from brick kilns | Pallor; Wax in ears; Nasal abnormalities; abdominal distention, enlargement of cervical nodes, tonsillitis (enlarged tonsils). |

| | | | | | | | | | |
|---|---|---|---|---|---|---|---|---|---|
| | | | | | | school health examination) | | | |
| Kamal 2014b[105] | Pakistan | Not reported | The authors sought to investigate the clinico-chemical parameters and level of exposure of BKWs to polycyclic aromatic hydrocarbons (PAHs). They enrolled adult BKWs and non-exposed controls aged ≥ 18 years and obtained urine and blood samples to measure urinary biomarkers of PAH exposure (i.e., 1-hydroxypyrene (1-OHPyr), α-naphthol and β-naphthol) and blood superoxide dismutase (SOD) and other hematologic parameters. Other than work groups, controls lived in cantonment areas away from brick kilns. Multiple brick kilns (at least 5 participants per kiln). Authors collected information of number of years worked but did not conduct an analysis linking health outcomes to the number of years worked. | Cross-sectional | Adult BKWs and unexposed controls | Mean age (± SD) was 41.1 ± 11.9 years (42.2 ± 13 in BKWs and 39.6 ± 10.3 in non-BKWs) | 100% males | 80 (46 BKWs and 34 controls) | Hematological parameters, C reactive protein, SOD, urinary biomarkers of PAH exposure (1-hydroxypyrene (1-OHPyr), α-naphthol and β-naphthol) |
| Kaushik 2012[79] | India | Not reported | Pulmonary function tests using the Sibelmed Datospir 120B precision portable spirometer. Participants had to do at least 3 acceptable maneuvers. Reproducibility and acceptability criteria used to select the best values following 2005 ATS/ERS guidelines. Oxidative stress biomarkers (serum malondialdehye, ferric reducing ability of plasma, serum glutathione S-transferase, an glutathione content in red blood cells) was collected. Genomic DNA was extracted from blood samples. Eligibility criteria was male brick workers, aged 20–40 years, who were occupationally exposed to silica in brick kilns and males not | Cross-sectional | Adult brick kiln workers and age-, sex-, and socioeconomic status-matched controls | Mean age (± SD) of BKWs 29.2 ± 4.8 years; of controls 28.3 ± 4.1 years | 100% males | 31 brick kiln workers and 32 controls | Pulmonary function tests including $FEV_1$, FVC, $FEV_1$/FVC, and peak expiratory flow were collected. Concentrations of oxidative stress biomarkers and DNA fragmentation or smearing was reported. |

| | | | | | | | | | |
|---|---|---|---|---|---|---|---|---|---|
| | | | exposed to brick works,selected from the staff working as sweepers, sanitary workers, daily wage laborers and security guards. Brick kiln workers and controls were matched on age, sex and socioeconomic status. Exclusion criteria were kiln workers with less than 1 year experience, on medication that affects respiratory function or a history of respiratory illness, upper respiratory tract infections in preceding three weeks or with any systemic illness (e.g., diabetes, tuberculosis, hypertension among others that were not specified). Number of brick kilns not reported. The authors evaluated for differences in lung function and oxidative stress parameters between BKWs with <5 and >5 years of experience. | | | | | | |
| Kazi 2019[80] | India | Dec 2016 – Oct 2017 | A multi-stage sampling was done with at least 10 workers per kiln from an unspecified number of kilns (reported number of kilns in the study area was 65). Data collection methods include standardized questionnaires and physical examination. Eligibility criteria included being available, having worked in brick kilns for 6 or more months, ≥ 18 years, not a casual laborer, work not limited to transportation of bricks for the purpose of distribution once final product was made. Multiple brick kilns. Authors collected information of number of years worked and evaluated for differences in musculoskeletal symptoms by number of years worked (5 or less vs. 6-10 years); and differences in respiratory symptoms by number of years worked (10 or less vs more than 10 years). The study stratified by BKW type (paatla, mhaapa, bigaari, and bhatkar). | Cross-sectional | Adult brick kiln workers from 65 kilns | Mean age (± SD) was 35.3 ± 11.0 years | 44.3% female, 55.7% males | 420 workers (186 females and 234 males) | Musculoskeletal complaints, respiratory complaints, skin complaints, heat stress, fever, injuries, eye complaints, GI complaints, burns, other complaints. |

| | | | | | | | | | |
|---|---|---|---|---|---|---|---|---|---|
| Khan 2019[95] | Pakistan | May 2016 – Apr 2017 | Structured questionnaires and physical examinations were performed on participants living in an exposed area to brick kilns and living in an unexposed area to brick kilns. Eligibility criteria were not specified. Multiple brick kilns (number of kilns not specified). | Cross-sectional | Adult and child residents of areas exposed and unexposed to brick kilns | Mean age (and SD) not provided. Age is presented in categories: 5-14 years (10% during non-crushing and 9% during crushing), 15-60 years (77% during non-crushingand 78% during crushing), >60 years (13% during non-crushing and crushing) in affected area; and 5-14 years (12% during non-crushing and crushing), 15-60 years (70% during non-crushing and crushing), and >60 years (18% during non-crushing and crushing) | 58% males and 42% females | 100 participants from the exposed area (58 males and 42 females), 100 participants from the unexposed areas (58 males and 42 females) | Acute respiratory infections; Asthma; Cough; COPD; Hypertension; diabetes mellitus |
| Khisroon 2018[81] | Pakistan | Apr 2014 - Mar 2015 | This study sought to evaluate DNA damage in individuals exposed to brick kiln pollution. The authors enrolled 100 BKWs from 20 kilns and a group of unexposed controls. Controls were healthy individuals having not history of occupational exposure to brick kilns or any other genotoxic chemicals. All participants were males. No other eligibility criteria were specified. Blood | Cross-sectional | Adult BKWs from 20 kilns and adult non-BKWs | Mean age (± SD) was 28.5 ± 6.2 years; for BKWs it was 28.4 ± 6.6 years; for non-BKWs it was 28.6 ± 5.4 years | 100% males | 150 (100 BKWs and 50 non-BKWs) | Total comet score |

| | | | | | | | | | |
|---|---|---|---|---|---|---|---|---|---|
| | | | samples were obtained from all participants. An alkaline comet assay and electrophoresis and neutralization, and staining with acridine orange were used to evaluate DNA damage. Multiple brick kilns. Authors collected number of years worked and evaluated differences in total comet score by number of years worked (<15 and ≥15 years). | | | | | | |
| Love 1999[106] | United Kingdom | Not reported | Workers at 18 brick and tile factories from 23 job groups. Modified MRC questionnaire was used for respiratory symptoms. Chest X-ray was obtained following a standard method. Respiratory dust and silica was collected through personal samplers and at specific locations to evaluate concentrations in areas where people passed through but did not carry out specific tasks. No eligibility criteria were specified. Multiple brick kilns. Authors collected number of years worked and calculated cumulative exposures to quartz and dust. They evaluated for an association between the presence of small opacities (≥0/1) and cumulative exposures to quartz and dust; and for associations between chronic bronchitis/COPD and cumulative exposure to dust. The authors record mean dust and quartz concentrations by BKW type and then use the mean exposure categories to stratify for health outcomes. | Cross-sectional | Adult employees at the brick and tile factories | Mean age 40 years (SD not reported) | 96.2% males, 3.8% females | 1925 employees (1852 men and 73 women) | Profusions of small opacities; Chronic bronchitis; Breathlessnes;Wheeze |
| Nasir 2021[7] | Pakistan | Sep – Oct 2018 | Children aged 5 - 12 years in areas within 3 km radius of brick kilns compared to those that are outside of this defined radius (controls). Measured height and cognitive ability using the Raven test score. This study used the 2007 WHO international reference to calculate Z-scores. Their analysis used propensity score matching (using 4 different matching | Cross-sectional | Children living within 3 km of brick kilns vs. those living further away | Mean age (± SD) was 9.0 ± 2.2 years for children living within 3 km of the brick kiln and 9.3 ± 1.9 years for | Not specified | 383 chlldren, 191 from kiln areas, 192 from control areas | Height (Z-scores) and cognitive ability. |

| | | | | | | | | | |
|---|---|---|---|---|---|---|---|---|---|
| | | | techniques) to calculate average treatment effect on the treated (ATT) is a probit regression. No other eligibility criteria were specified. Multiple brick kilns (number not specified). | | | control children | | | |
| Rahman 2013[82] | Pakistan | Oct – Nov 2007 | Spirometry was conducted in the standing position with the Microloop/MicroDirect office spirometer following the 1994 American Thoracic Society guidelines. Eligibility criteria include 25-65 years, at least 5 years working in brick kilns (if from the brick kiln worker group), no history of chronic disease (tuberculosis, asthma, mental disability) and no history of smoking. Controls (from the surrounding communities) were matched to brick kiln workers by age, sex and education, who were not working in pottery or other types of kilns (e.g., iron). Number of brick kilns not reported. Authors collected number of years worked. | Cross-sectional | Adult brick kiln workers and non-kiln workers from the surrounding communities | Mean age 35 years (SD not reported) | Sex distribution was not reported | 407 brick kiln workers and 407 non kiln workers | Cough; Dyspnea; COPD (although use of bronchodilators in spirometry was not reported); Raw lung function values were not reported but they reported average percent predicted for BKWs only |
| Raza 2014[107] | Pakistan | Fall 2009 – Spring 2010 | Lung function was assessed with a portable digital spirometer (Spirolab III, SN A23- 053, MIR, Italy). No eligibility crtieria were specified. Multiple brick kilns (number of brick kilns not specified). Authors collected information on the number of years worked and evaluated for an association between $FEV_1$ or FVC and number of years worked. They stratified outcomes by BKW type (thapai, modulation, loading, keri, burning, and unloading). | Cross-sectional | Adult and child brick kiln workers | Mean (and SD) age not reported. Age is reported as the number of participants in four age categories: 11-25 years (36%), 26-40 years (31%), 41-55 years (27%), 56-70 years (6%). | Not reported | 156 | $FEV_1$, FVC and $FEV_1$/FVC were reported as percent predicted values (reference population not specified). |
| Raza 2018[83] | Pakistan | Not reported | This study sought to evaluate Redox balance and DNA fragmentation in arsenic-exposed occupational workers from different industries. Blood, hair, and nail samples were collected from | Cross-sectional | Arsenic exposed workers (including BKWs) and | Mean age (± SD) was 27.3 ± 17.1 years; for BKWs it was | Not reported | 450 (50 BKWs) | DNA fragmentation. Antioxidant enzymes (catalase activity, |

| | | | | | | | | | |
|---|---|---|---|---|---|---|---|---|---|
| | | | welding, brick kiln, furniture, pesticide, and paint industries (n = 50/industry) of Pakistan along with age- and sex-matched 200 controls (eligibility criteria of controls was not specified). DNA damage was calculated using DNA fragmentation assay. Antioxidant enzymes (catalase activity, superoxide dismutase, glutathione peroxidase) were measured using ELISA. Number of kilns not reported. The authors collected information on number of years worked (categorized as less than 10 or more than 10 years) but did not appear to evaluate the association between CAT/GPx/SOD and number of years worked in brick kilns. | | unexposed controls. | 30,4 ± 15.1 years; and unexposed controls it was 27 ± 21.9 years. | | | superoxide dismutase, glutathione peroxidase) |
| Raza 2021[108] | Pakistan | Jan – Apr 2018 | Questionnaires were administered for respiratory symptoms. Lung function was assessed with a portable digital spirometer (SP10 Spiroton, MDX Instruments) following 2005 ATS/ERS guidelines. Eligibility criteria was age 18-60 years, at least 1 year of brick kiln work experience, apparently health exposed to kiln pollution for at least 8 hours daily over 1+ years, agreed to participate voluntarily and signed consent, no abnormalities in vertebral column, thoracic cage or neuromuscular disease, no previous diagnosis of ischemic heart disease, lung cancer, tuberculosis, cor pulmonale, pulmonary effusion, pneumonia or history of abdominal or thoracic surgery. Multiple brick kilns. Authors collected information on number of years worked but did not analyze relationship between health outcomes and number of years worked. They stratified by BKW type (modulation, loading, burning, and unloading). | Cross-sectional | Adult brick kiln workers in 3 kilns from the modulation area (mainly exposed to dust but sometimes exposed to pollutants from kiln chimney) and kiln area (exposed to kiln chimney pollutants, kiln dust and high temperature) | Mean (and SD) age was not reported. | Not reported | 45 workers in the kiln area and 15 in the modulation area | $FEV_1$; FVC; $FEV_1$/FVC |

| | | | | | | | | | |
|---|---|---|---|---|---|---|---|---|---|
| Raza 2022[84] | Pakistan | 2017 – 2019 | Questionnaires were administered for respiratory symptoms. Lung function was assessed with a portable digital spirometer (SP10 Spiroton, MDX Instruments) following 2005 ATS/ERS guidelines. Eligibility criteria was age 18-60 years, at least 1 year of brick kiln work experience, apparently health exposed to kiln pollution for at least 8 hours daily over 1+ years, agreed to participate voluntarily and signed consent, no abnormalities in vertebral column, thoracic cage or neuromuscular disease, no previous diagnosis of ischemic heart disease, lung cancer, tuberculosis, cor pulmonale, pulmonary effusion, pneumonia or history of abdominal or thoracic surgery. Multiple brick kilns. Authors did not collect information on number of years worked. | Cross-sectional | Adult brick kiln workers from 15 kilns | Mean age not provided. Age was reported as the number of participants across 7 age categories. | 83% males, 17% males | 506 (419 males and 87 females) | Cough; phlegm; wheeze; Dyspnea (grades I and II); self-reported asthma; physician-diagnosed asthma; coughing up blood; chest pain; chest tightness; dry nose; dry throat; throat pain; $FEV_1$; FVC; percent predicted values of $FEV_1$ and FVC; $FEV_1$/FVC (reference population not reported). |
| Roshania 2022[85] | India | Jun 2018, Jan 2019 | Using a stratified cluster design, authors conducted two waves of primary data collection among 2564 randomly selected circular migrant children under three years of age temporarily residing across 1156 brick kilns in Bihar, India, and conducted multilevel modeling to estimate the association of the number of migration episodes and age at first migration with stunting and wasting. Inclusion criteria were self-identification as a circular migrant household, defined as living away from their home block (sub-district) for employment purposes for a total of at least 60 days in the previous year, with at least one return home during that year; and presence of at least one child under three years of age at the kiln. Using a random number table, three eligible families per kiln were selected, one with a child in the 0 to 11-month age group and | Cross-sectional, cluster sampling | Children aged < 36 months | 14.6% of children were aged 0-5 months; 17,7% were aged 6-11 months; 44.1% were aged 12-23 months; 23.6% were aged 24-35 months | 50.7% boys, 49.3% girls | 2564 children living in 1156 brick kilns | Height and weight (Z-scores) |

| | | | | | | | | | |
|---|---|---|---|---|---|---|---|---|---|
| | | | two with a child in the 12 to 35-month age group. Multiple brick kilns. | | | | | | |
| Saldaña-Villanueva 2023[86] | Mexico | Not reported | Assessment of spirometry using the EasyOne spirometer (ndd) following 2005 ATS/ERS guidelines. The predicted forced vital capacity (%FVC) and forced expiratory volume in the first second (%$FEV_1$) were estimated for each subject based on their sex, age, and height, using the formulas provided by Perez-Padilla. Blood and urine samples for serum and urine biomarkers. Three types of workers (mercury miners, brick kiln workers and quarry workers) were included in the study. No other eligibility criteria were specified. Number of brick kilns was not reported. The authors report on number of years worked, but they did not evaluate the association between health outcomes and number of years worked. | Cross-sectional study | Adults: 34 miners dedicated to mercury extraction: 40 brick kiln workers; and 36 quarry workers | Median age was 49 years (range 20 – 86) | 100% males | 110 workers | $FEV_1$, FVC, PEF, $FEV_1$/FVC, % predicted $FEV_1$, % predicted FVC; Serum Creatinine, HbA1C, estimated GFR. |
| Sanjel 2016[109] | Nepal | Mar 2015 – Apr 2016 | Environmental exposures to total dust, $PM_{10}$ and $PM_{2.5}$ was measured with the Dusttrack monitor. Questionnaires to gather information on injuries and health problems (breathlessness, persistent cough, eye problems, skin problems, stomach problems/diarrhea, fever, headache, extreme fatigue, feeling bad all over) were used. Eligibility criteria were not specified. Multiple brick kilns. Authors collected information on the number of years worked. | Cross-sectional | Adult and child brick kiln workers and grocery workers | Mean (± SD) age was 31.7 ± 13.0 years in brick kiln workers and 33.3 ± 9.0 in grocery workers | 71% males, 29% females | 400 brick kiln workers (102 females and 298 males) and 400 grocery workers (130 females and 270 males) | Breathlessness; Persistent cough; Eye problems; Skin problems; Stomach problems/diarrhea; Fever; Headache; Extreme fatigue; Feeling bad all over |
| Sanjel 2017[94] | Nepal | Feb 2015 – April 2016 | Same study as Sanjel 2016, but this study reports on chronic cough (4-6 times per day occurring on most days of the week), chronic phlegm (twice a day for most days of the week and at least 3 months of the year for at least 2 consecutive years); chronic | Cross-sectional | Adult and child brick kiln workers and grocery workers | Same as above | Same as above | Same as above | Chronic cough; Chronic phlegm; Chronic bronchitis; Frequent wheeze; Wheeze with |

| | | | | | | | | | |
|---|---|---|---|---|---|---|---|---|---|
| | | | bronchitis (chronic cough and phlegm on most days of the week and at least 3 months of the year for at least 2 consecutive years); and wheezing (chest sounds wheezy or whistling most days or nights in past 2 months). Multiple brick kilns. Authors collected information on the number of years worked and reported work duration as a significant predictor of adverse health outcomes. The study stratified by BKW type (green brick molding and stacking/carrying, red brick loading/carrying, coal preparation, and firing). | | | | | | shortness of breath |
| Shaikh 2012[5] | Pakistan | Apr – May 2011 | Respiratory symptom and illness history taken using the American Thoracic Society Division of Lung Disease questionnaire. Convenience sample to select brick kilns and workers. Eligibility criteria are male workers above 18 years of age who have worked at the kilns for at least 5 years. Multiple brick kilns. Authors reported work duration of 5-9 years and greater than or equal to 10 years. The study stratified by BKW type (carrier, baker, and molder). | Cross-sectional | Adult brick kiln workers | Mean (± SD) age was 31.0 ± 9.1 years | 100% males | 340 | Chronic cough; Chronic phlegm; Chronic bronchitis; Dyspnea; Asthma |
| Sherris 2021[87] | Bangladesh | Jan 2005 – Dec 2014 | Time series analysis of clinical records of physician-diagnosed pneumonia/upper respiratory infections and particulate matter (evaluated for source contributions). No other eligibility criteria were specified. Multiple brick kilns (number not specified). | Time series analysis | Children in Kamalapur, Dhaka | % for ages 0, 1, 2, 3, 4, 5 were 33%, 35%, 19%, 9%, and 4% for pneumonia; and 21%, 30%, 22%, 16%, and 11% for upper respiratory infections | Pneumonia cases had 56% boys; upper respiratory cases had 51% boys | 28,089 clinic visits | Pneumonia; upper respiratory infections |

| | | | | | | | | | |
|---|---|---|---|---|---|---|---|---|---|
| Sheta 2015[88] | Egypt | Not reported | Spirometry was conducted. Chest X-rays were obtained. A questionnaire recommended by the American Thoracic Society was used to collect data on chronic respiratory diseases. Eligibility criteria for brick kiln workers with exposures to dust and smoke in kiln and working as molders or bakers in the kiln industry for at least 5 years; no history of exposure to dust or smoke due to other causes or previous history of respiratory disease (bronchial asthma, chronic bronchitis, pulmonary tuberculosis and pneumonia). Eligibility criteria for the control group was no history of exposure to dust or smoke in job and matched to brick kiln workers by sociodemographic characteristics. Multiple brick kilns. Authors reported work duration of 5-10 years and greater than or equal to 10 years and analyzed the odds of chronic respiratory disease based on work duration. The study stratified by BKW type (bakers and molders). | Cross-sectional | Adult brick kiln workers and reference group not exposed to smoke or dust | Not reported | 100% males | Brick kiln workers (173) and non-kiln workers (170) | Chronic cough; Chronic bronchitis; Chronic phlegm; Dyspnea; Wheeze; Asthma; $FEV_1$; FVC; $FEV_1$/FVC; $FEF_{25-75}$; Percent predicted values for $FEV_1$ and FVC (reference population not reported) |
| Shrestha 2021[89] | India | Nov 2019 – May 2020 | Questionnaires for health outcomes. Participants were selected based on a probability proportional to size based on occupational category (molder, loader, firemen). Multiple brick kilns. Authors reported work duration of 0-10, 11- 20, and 21-30 years and included disease prevalence for each category. | Cross-sectional | Adult brick kiln workers aged 25-65 years | % of participants aged 24-35, 36-50, 51-65 years were 29.1%, 54.9% and 16% | Not reported | 450 | Self-reported history of tuberculosis, injury, skin disease, respiratory disease, eye disease, and musculoskeletal disorder |
| Sinaga 2022[90] | Indonesia | Feb – Dec 2020 | Assessment of height in non-twin children aged 0-24 months living within 2 km of brick kilns whose families have been living there for more than 5 years in 2 villages, and children aged 0-24 months in two villages that did not have brick kilns. If two babies were found in the family, only one was selected. | Cross-sectional | Children living in four villages of the Pagar Merbau Subdistrict, two villages have brick kilns and two | Ages 0-24 months, mean age not reported | Not reported | 192 | Height (Z-scores)/stunting |

| | | | | | | | | | |
|---|---|---|---|---|---|---|---|---|---|
| | | | | | villages did not | | | | |
| Srivastava 2002[91] | India | Not reported | Questionnaires were used for self-reported symptoms. Lung function was done using a portable spirometer (HI-298; Chest, Japan) in standing position following 1979 American Thoracic Society recommendations. A subsample of 46 participants had a chest x-ray. Eligibility criteria were not specified. Multiple brick kilns. The study stratified by BKW type (baharai, furnace, nikasi, pathai, and others). | Cross-sectional | Adult brick kiln workers and non-kiln workers | Mean (± SD) age was 31.3 ± 9.9 years in brick kiln workers and 33.4 ± 8.7 years in controls | 95% males; 5% females | 257 brick kiln workers (244 males and 13 females) and 131 controls (116 males and 15 females) | Chronic bronchitis; Silicosis; Eye irritation; Musculoskeletal pain; Malnutrition; Diarrhea; Respiratory infections; Chest X-ray findings in a subsample; Respiratory morbidity; Percent predicted values of vital capacity, FVC, $FEV_1$/FVC and MMEF stratified by smoking status (reference population not reported) |
| Subhanullah 2022[92] | Pakistan | Dec 2018 | Questionnaires on self-reported health outcomes. No eligibility criteria were specified. Number of brick kilns not reported. | Cross-sectional | Adult and child brick kiln workers | Not reported | Not reported | 50 | Eye irritation, Respiratory disorders, headache, and skin disorders |
| Tandon 2017[114] | India | 2010 – 2012 | Non-smoking male brick kiln workers and an age-matched reference group of males. Spirometry was conducted using a portable digital spirometer (Medspiror, Recorders and Medicare Systems Pvt Ltd, Chandigarh, India). Eligibility criteria were age 18-35 years, males, apparently healthy, have completed primary school. Excluded if illiterate; had a history of wheezing, smoking, tobacco chewing, alcohol abuse, cardiac and/or respiratory illnesses; systemic illnesses | Cross-sectional study | Adult brick kiln workers and non-kiln workers | Mean (± SD) age was 31.6 ± 4.9 in non-smoking brick kiln workers and 30.6 ± 4.4 in the reference group. | 100% non-smoking males | 110 brick kiln workers and 90 controls | Frequent coughing; Shortness of breath; Irritation of the respiratory tract; Chronic phlegm; Chest tightness; $FEV_1$; FVC; $FEV_1$/FVC; $FEF_{25-75\%}$; Peak expiratory flow; Percent predicted values for $FEV_1$, |

| | | | | | | | | |
|---|---|---|---|---|---|---|---|---|
| | | | (hypertension, diabetes, thyroid disorders), visible musculoskeletal deformities or injury of chest wall; use of medications that could affect the outcome of the study (asthma medicines, anti-depressants). Multiple brick kilns. The authors analyzed respiratory outcomes based on a work duration of either <8 or >/= 8 years. | | | | | | FVC, $FEV_1$/FVC, $FEF_{25-75\%}$ and peak expiratory flow (reference population not reported). |
| Thomas 2015[93] | India | Aug 2011 – Aug 2022 | This study sought to evaluate the prevalence of chest symptoms among brick kiln workers and care seeking behavior. They enrolled 4002 participants from 55 brick kilns who were 18 years of age and older who worked for at least 3 months. A questionnaire asked about chest symptoms (as a patient who has productive cough for more than 2 weeks or more accompanied, with or without chest pain, intermittent fever and/or a history of hemoptysis). The authors also asked about alcohol use using the Alcohol Use Disorders Identification Test (AUDIT) questionnaire and quantified smoking history. Multiple brick kilns. The authors reported chest symptom prevalence based on brick kiln chamber work duration of </= 4, 5-6, or >6 months. | Cross-sectional study | Adult and child BKWs | Mean age not reported. Instead, age was presented in categories: 15-24 years (25.7%), 25-34 years (29.1%), 35-44 years (21.5%), 45-54 years (15.5%), and ≥ 55 years (8.1%). | 52% male | 4002 | Chest symptoms. AUDIT score. |
| Vaidya 2015[110] | India | 2008 - 2010 | Questionnaires and physical exams to evaluate for backache, headache, vision problems, burning sensation in hands and feet, cough, breathlessness, tremors in fingers and inability to sleep. Assessment of carbon monoxide and silica concentrations at fixed sites. Eligibility criteria were all women aged 18-40 years working at study sites who were present on the day of the study and were willing to participate. Multiple brick kilns. The authors reported work | Cross-sectional | Adult BKWs and reference participants (construction workers) | Mean age not provided. Age was reported as the number of participants across 5 age categories. | 100% females | 40 BKWs and 63 controls | Cough; Breathlessness; Backache, Headache, Vision problems, Burning sensation in hands and feet; Tremors in fingers; Inability to sleep. |

| | | | | | | | | | |
|---|---|---|---|---|---|---|---|---|---|
| | | | duration of <1, 1-5, or >5 years, but do not use this data in their analysis. | | | | | | |
| Zawilla 2014[111] | Egypt | Not reported | This study sought to evaluate liver function in silica-exposed workers in Egypt. Exposed workers were recruited from a clay brick factory. Single brick kiln. The control subjects were security personnel and administrative workers in a small insurance company, matched to BKWs by age, body mass index, and smoking status. Controls did not reside in the same industrial area as the exposed workers and were never occupationally exposed to silica dust or hepatotoxins. Exclusion criteria for both the exposed and control groups were: any history of alcohol consumption, BMI> 30 kg/$m^2$, use of drugs with possible hepatotoxic effect, diabetes, uncontrolled hypertension, current or previous viral hepatitis, schistosomiasis, or an autoimmune disease. The authors reported duration of employment but do not use this data in their analysis. | Cross-sectional | BKWs and unexposed controls | Mean (± SD) age was 47.4 ± 7.1 years; for BKWs it was 47.4 ± 7.3 years; for non-BKWs it was 47.5 ± 6.7 years. | Not reported | 87 BKWs and 45 controls | Albumin, ALT, AST, GGT, Bilirubin, MMP-9, IgG, IgE. Silicosis. |

**Table S12. Quality control of studies reporting health data (Newcastle-Ottawa Scale for cross-sectional studies).**

| | Selection | | | | Comparability | Outcome | Total stars |
|---|---|---|---|---|---|---|---|
| | Representativeness of sample | Selection of reference group | Ascertainment of exposure (or disease) | Non-respondents | | | |
| Ahmad 2020[97] | 0 | 0 | 1 | 0 | 2 | Assessment: 2<br>Statistics: 1 | 6 |
| Akram 2022[98] | 0 | 0 | 1 | 0 | 2 | Assessment: 2<br>Statistics: 1 | 6 |
| Ali 2013[69] | 0 | 1 | 1 | 0 | 0 | Assessment: 1<br>Statistics: 0 | 3 |
| Berumen-Rodriguez 2021[100] | 0 | 0 | 1 | 0 | 0 | Assessment: 2<br>Statistics: 0 | 3 |
| Berumen-Rodriguez 2023[99] | 0 | 0 | 1 | 0 | 0 | Assessment: 2<br>Statistics: 0 | 3 |
| Biswas 2018[70] | 1 | 0 | 1 | 0 | 0 | Assessment: 2<br>Statistics: 1 | 5 |
| Das 2014[71] | 1 | 1 | 1 | 0 | 0 | Assessment: 2<br>Statistics: 0 | 5 |
| Das 2019a[73] | 0 | 1 | 1 | 0 | 0 | Assessment: 2<br>Statistics: 0 | 4 |
| Das 2019b[72] | 1 | 0 | 1 | 0 | 0 | Assessment: 2<br>Statistics: 0 | 4 |
| David 2020[74] | 1 | 1 | 1 | 0 | 0 | Assessment: 2<br>Statistics: 0 | 5 |
| David 2021[101] | 1 | 1 | 1 | 0 | 0 | Assessment: 2<br>Statistics: 0 | 5 |
| David 2022[9] | 1 | 1 | 1 | 0 | 0 | Assessment: 2<br>Statistics: 0 | 5 |
| Erdim 2020[75] | 0 | 0 | 1 | 0 | 0 | Assessment: 2<br>Statistics: 0 | 3 |
| Goel 2015[76] | 0 | 1 | 1 | 0 | 0 | Assessment: 2<br>Statistics: 0 | 4 |
| Gonzalez 2021[77] | 1 | 0 | 1 | 0 | 0 | Assessment: 2<br>Statistics: 0 | 4 |
| Gupta 2019[78] | 1 | 0 | 1 | 0 | 0 | Assessment: 2<br>Statistics: 0 | 4 |
| Hamid 2023[96] | 1 | 0 | 1 | 1 | 0 | Assessment: 1<br>Statistics: 0 | 4 |
| Jahan 2016[102] | 0 | 0 | 1 | 0 | 1 | Assessment: 2<br>Statistics: 1 | 5 |

| | | | | | | | |
|---|---|---|---|---|---|---|---|
| Joshi 2008[103] | 0 | 1 | 2 | 0 | 1 | Assessment: 2<br>Statistics: 0 | 6 |
| Kamal 2014b[105] | 1 | 1 | 1 | 1 | 0 | Assessment: 2<br>Statistics: 1 | 7 |
| Kaushik 2012[79] | 0 | 0 | 1 | 0 | 2 | Assessment: 2<br>Statistics: 0 | 5 |
| Kazi 2019[80] | 1 | 0 | 1 | 0 | 0 | Assessment: 0<br>Statistics: 0 | 2 |
| Khan 2019[95] | 0 | 0 | 2 | 0 | 0 | Assessment: 1<br>Statistics: 0 | 3 |
| Khisroon 2018[81] | 0 | 1 | 1 | 0 | 0 | Assessment: 2<br>Statistics: 0 | 4 |
| Love 1999[106] | 1 | 0 | 2 | 1 | 2 | Assessment: 2<br>Statistics: 1 | 9 |
| Nasir 2021[7] | 1 | 1 | 2 | 0 | 2 | Assessment: 2<br>Statistics: 1 | 9 |
| Rahman 2013[82] | 1 | 1 | 1 | 0 | 2 | Assessment: 1<br>Statistics: 0 | 6 |
| Raza 2014[107] | 0 | 0 | 2 | 0 | 0 | Assessment: 2<br>Statistics: 0 | 4 |
| Raza 2018[83] | 0 | 1 | 1 | 0 | 2 | Assessment: 2<br>Statistics: 0 | 6 |
| Raza 2021[108] | 0 | 0 | 1 | 1 | 0 | Assessment: 2<br>Statistics: 0 | 4 |
| Raza 2022[84] | 1 | 1 | 2 | 0 | 0 | Assessment: 2<br>Statistics: 0 | 6 |
| Roshania 2022[85] | 1 | 1 | 1 | 0 | 2 | Assessment: 2<br>Statistics: 1 | 8 |
| Saldaña-Villanueva 2023[86] | 0 | 0 | 1 | 0 | 0 | Assessment: 2<br>Statistics: 0 | 3 |
| Sanjel 2016[109] | 1 | 0 | 2 | 0 | 0 | Assessment: 1<br>Statistics: 0 | 4 |
| Sanjel 2017[94] | 1 | 0 | 2 | 0 | 0 | Assessment: 1<br>Statistics: 0 | 4 |
| Shaikh 2012[5] | 0 | 0 | 1 | 0 | 2 | Assessment: 2<br>Statistics: 0 | 5 |
| Sherris 2021[87] | 1 | 0 | 2 | 0 | 2 | Assessment: 2<br>Statistics: 1 | 8 |
| Sheta 2015[88] | 1 | 1 | 2 | 0 | 1 | Assessment: 2<br>Statistics: 0 | 7 |
| Shrestha 2021[89] | 0 | 0 | 1 | 0 | 0 | Assessment: 1<br>Statistics: 0 | 2 |

| | | | | | | | |
|---|---|---|---|---|---|---|---|
| Sinaga 2022[90] | 0 | 0 | 1 | 0 | 0 | Assessment: 1<br>Statistics: 0 | 2 |
| Srivastava 2002[91] | 0 | 0 | 1 | 0 | 1 | Assessment: 2<br>Statistics: 0 | 4 |
| Subhanullah 2022[92] | 1 | 0 | 2 | 0 | 0 | c) Assessment: 1<br>Statistics: 0 | 4 |
| Tandon 2017[114] | 1 | 1 | 2 | 0 | 2 | Assessment: 2<br>Statistics: 0 | 8 |
| Thomas 2015[93] | 0 | 0 | 1 | 0 | 0 | Assessment: 1<br>Statistics: 0 | 2 |
| Vaidya 2015[110] | 1 | 0 | 2 | 0 | 0 | Assessment: 1<br>Statistics: 0 | 4 |
| Zawilla 2014[111] | 1 | 1 | 1 | 1 | 2 | Assessment: 2<br>Statistics: 0 | 8 |

**Table S13. Summary of respiratory health data reported in the studies.**

| | | |
|---|---|---|
| **Population** | **n (%)** | **References** |
| Adults only | 23 (68%) | 5,9,70–72,76–80,82,84,86,88,89,91,99,100,106,108,110,111,114 |
| BKWs only | 17 (68%) | 5,70,72,77,78,80,84,86,89,92,93,96,99,100,106–108 |
| BKWs and reference group | 12 (35%) | 9,71,73,76,79,82,88,91,94,109,110,114 |
| Men only | 11 (32%) | 5,9,71,76,79,86,88,99,100,111,114 |
| Adults and children | 7 (21%) | 92–96,107,109 |
| Children only | 4 (12%) | 69,73,87,103 |
| Participants living near or far from kilns | 3 (9%) | 69,95,103 |
| Women only | 1 (3%) | 110 |
| **Health outcomes** | | |
| Lung function | 14 (41%) | 71–73,76,79,82,84,86,88,91,100,107,108,114 |
| Cough or chronic cough | 14 (41%) | 5,70,77,78,82,84,88,94–96,108–110,114 |
| Breathlessness, dyspnea or shortness of breath | 11 (32%) | 5,77,78,82,84,88,106,108–110,114 |
| Phlegm or chronic phlegm | 9 (26%) | 5,70,77,78,84,88,94,108,114 |
| Wheezing | 8 (24%) | 5,70,77,84,88,94,106,108 |
| Asthma | 8 (24%) | 5,9,70,84,88,94,95,108 |
| Chronic bronchitis | 7 (21%) | 5,70,78,88,91,94,106 |
| COPD | 4 (12%) | 82,95,99,100 |
| **Examined associations** | | |
| Health outcomes by number of years worked at the kilns | 10 (29%) | 5,70,79,80,88,89,100,106,107,114 |
| Health outcomes by type of BKW occupation | 10 (29%) | 5,70,77,80,84,88,89,94,107,108 |
| **Location** | | |
| Multiple brick kilns | 27 (79%) | 5,9,70–73,77,78,80,84,87–89,91,93–96,99,100,103,106–110,114 |

**Table S14. Summary of selected biomarkers and hematological parameters between brick kiln workers (BKW) and controls (or reference group).**

| | | | Mean (SD) | |
|---|---|---|---|---|
| **Biomarkers, Authors** | | **Units** | **BKW** | **Control** |
| **Superoxide dismutase** | | | | |
| | David 2020[74] | U/mg protein | 6.74 (0.98) | 7.31 (1.71) |
| | David 2021[101] | U/mg protein | 0.24 (16.67) | 1.03 (47.42) |
| | David 2022[9] | U/mg protein | 14.4 (9.11) | 16.9 (10.18) |
| | Kamal 2014b[105] | U/mg Hb | 2.30 (0.83) | 1.11 (0.29) |
| | Raza 2018[83] | ng/mL | 1.89 (5.94) | 6.43 (7.04) |
| **Peroxidase** | | | | |
| | David 2020[74] | U/min | 19.17 (12.06) | 22.54 (7.79) |
| | David 2021[101] | U/min | 1.01 (6.02) | 6.24 (9.4) |
| | David 2022[9] | nmol | 19.2 (26.79) | 21.7 (31.4) |
| **Catalase** | | | | |
| | David 2021[101] | U/mg | 0.20 (2.89) | 0.50 (5.44) |
| | Raza 2018[83] | ng/mL | 74.85 (5.72) | 81.38 (16.72) |
| **Reactive oxygen species** | | | | |
| | David 2020[74] | nmol | 3.41 (2.39) | 2.24 (4.27) |
| | David 2021[101] | n/mol) | 1.87 (3.24) | 0.87 (0.06) |
| | David 2022[9] | μmol/min | 1.50 (1.30) | 1.17 (0.85) |
| **Cortisol** | | | | |
| | David 2020[74] | ng/mL | 0.73 (0.54) | 0.53 (0.53) |
| | David 2021[101] | ng/mL | 1.81 (0.58) | 0.83 (1.39) |
| | David 2022[9] | ng/mL | 3.04 (0.96) | 1.83 (0.58) |
| | Overall | | 2.31 (1.23) | 1.23 (1.02) |
| **Hemoglobin** | | | | |
| | David 2020[74] | g/dL | 7.40 (2.06) | 10.92 (2.14) |
| | David 2021[101] | g/dL | 12.79 (1.6) | 12.05 (1.78) |
| | David 2022[9] | g/dL | 12.5 (7.63) | 14.1 (3.68) |
| | Kamal 2014b[105] | g/dL | 13.8 (1.6) | 15.2 (0.7) |
| | Overall | | 11.72 (6.09) | 12.92 (3.19) |

**Figure S1. CO emission factors versus modified combustion efficiency (MCE) in (a) g/kg brick, (b) g/kg fuel, and (c) g/MJ.** We plot CO emission factors versus MCE, categorized by brick kiln design and fuel used. Symbols in red and blue represent kiln types categorized as traditional and improved, respectively. Symbols in grey represent kilns whose type was not specified. Brick kilns that used coal only are shown as squares, those that used coal and biomass are shown as triangles, and those that used biomass only are shown as circles. Means for traditional and improved kiln designs are show in the red and blue diamonds, respectively.

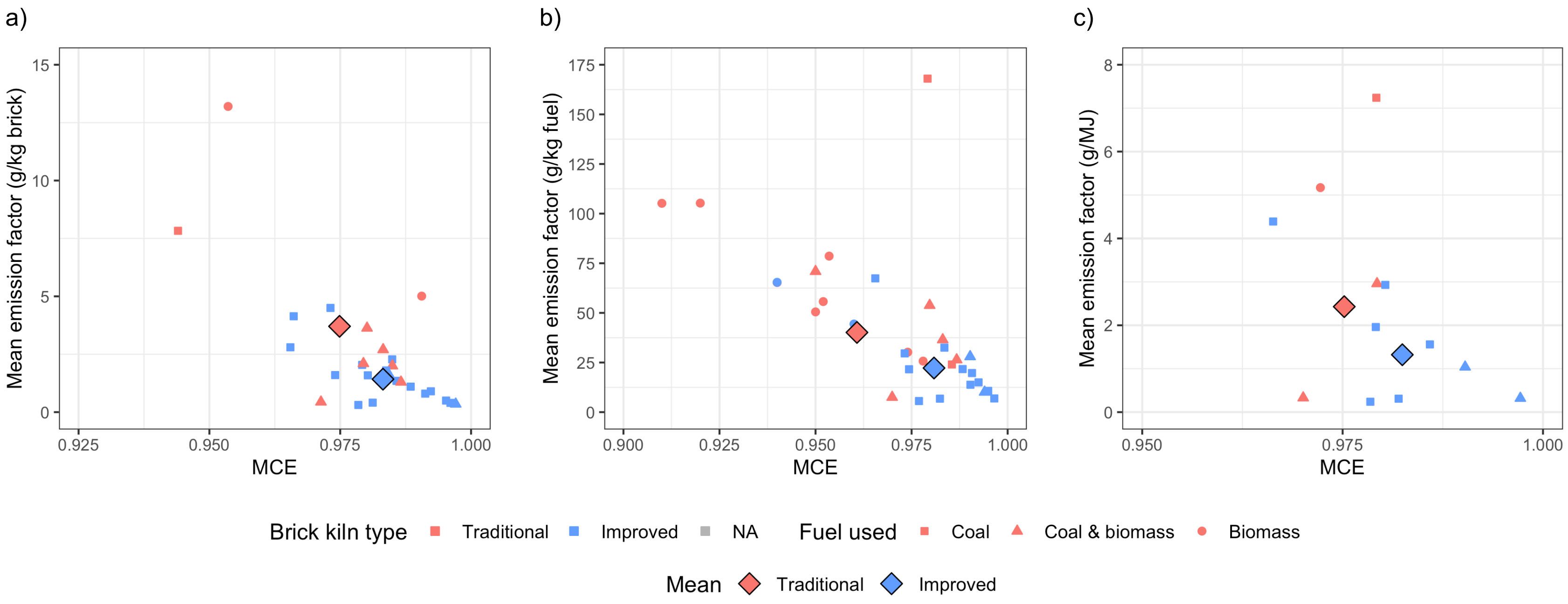
